# Common centromeric variation associates with molecular and complex traits in humans

**DOI:** 10.64898/2026.10.09.26365154

**Authors:** Yikai Dong, David Cutler, Joshua Weinstock

**Affiliations:** Department of Human Genetics, Emory University School of Medicine, Atlanta, GA, USA

## Abstract

Human centromeres harbor extensive structural variation that has remained largely inaccessible to human genetics because their repetitive *α*-satellite arrays are absent from standard reference and imputation panels. Here we catalog 188,589 centromeric structural variants (SVs) from haplotype-resolved long-read assemblies of 196 Human Pangenome Reference Consortium individuals aligned to the telomere-to-telomere CHM13 reference. Resolving recurrent SVs into monophyletic components along centromeric haplotype phylogenies makes them imputable from pericentromeric SNPs (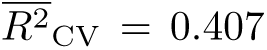 vs. 0.065 unresolved). In 124 individuals with haplotype-resolved nanopore methylation, we identify 418 same-chromosome SV–CpG associations that form local hypomethylation-enriched domains in the centromeric region, most strongly for deletions. We also show that a truncated 113-bp monomer on chromosome 8 is associated with the position of the CENP-A-associated centromere dip region (CDR) in 151 individuals, independent of array length. Imputed into 630 individuals with expression and splicing data, centromeric SVs are associated with 141 genes, 14 of them new relative to the SNP-based cis-QTL analysis, including the H3K9 methyltrans-ferase *SUV39H1*. In 315,204 All of Us participants, we identify 170 associations between 61 SV predictors and 27 clinical phenotypes, including substance-related disorders and breast cancer. Our study brings a previously inaccessible class of variation into population-scale genetics and links centromeric SVs to centromere organization, gene expression, splicing and complex traits.

## 1 Introduction

Centromeres are essential chromosomal structures that direct accurate chromosome segregation during cell division through the recruitment of the kinetochore [McKinley and Cheeseman, 2016], yet they embody a well-known evolutionary paradox: their strictly conserved function is encoded by some of the most rapidly evolving sequences in the genome [Henikoff et al., 2001]. Despite their central role in cell division, centromeres have remained among the least characterized regions of the human genome. Human centromeres consist predominantly of *α*-satellite monomers (171 bp) organized into higher-order repeat (HOR) arrays that span hundreds of kilobases to several megabases, preventing the unique alignment of short sequencing reads (50 to 300 bp). Due to this technical challenge, centromeres have been excluded from genome-wide association studies (GWAS), expression quantitative trait locus (eQTL) analyses, and clinical variant interpretation [Altemose et al., 2022, Miga and Alexandrov, 2021, Aganezov et al., 2022]. Common centromeric variation across humans therefore remains poorly understood, and its contribution to complex trait variation is entirely unknown.

The completion of the telomere-to-telomere (T2T) CHM13 assembly provided the first gapless representation of human centromeric sequences across all autosomes and the X chromosome [Nurk et al., 2022]. Recently, the Human Pangenome Reference Consortium (HPRC) [Liao et al., 2023] and the Human Genome Structural Variation Consortium (HGSVC) [Ebert et al., 2021] have produced haplotype-resolved long-read assemblies from diverse human populations, which enable the cataloging of centromeric structural variants across the genome.

Many reports show that centromere dysfunction contributes to disease risk. Kinetochore dysfunction is an important source of chromosome missegregation [Cimini et al., 2001, Zielinska et al., 2015], including Robertsonian translocations and Down syndrome [de Lima et al., 2025]. Moreover, such errors drive the chromosomal instability that is a hallmark of cancer, in which pericentromeric satellite arrays are themselves expanded and rearranged [Barra and Fachinetti, 2018, Chen et al., 2025, Bersani et al., 2015]. However, testing whether common inherited centromeric structural variation is also associated with complex traits requires genotypes for that variation in large phenotyped cohorts.

Structural variants (SVs; insertions, deletions, duplications, and other rearrangements of 50 or more base pairs) are a major source of human genetic diversity and influence gene expression and complex traits [Bai et al., 2026]. Expression-altering SVs tend to have larger effects than single-nucleotide variants [Chiang et al., 2017, Scott et al., 2021]. Centromeric SVs, however, can be directly genotyped only in the few individuals with long-read assemblies, for example, the *∼*232 individuals in HPRC Release 2 [Lucas et al., 2026]. Extending these genotypes to the hundreds of thousands of samples in existing biobank and population cohorts, which have only SNP-array or short-read whole-genome sequencing (WGS) data, would increase power for discovery of links between the centromere and complex traits; however, existing imputation panels exclude centromeric variation [Taliun et al., 2021, McCarthy et al., 2016].

To address these limitations, we develop a framework for imputing centromeric SV genotypes from pericentromeric SNPs using an elastic-net strategy analogous to PrediXcan/TWAS [Gamazon et al., 2015, Gusev et al., 2016]. Recombination is strongly suppressed within centromeric regions [Langley et al., 2019], creating extended haplotype blocks that link centromeric SVs to flanking SNP genotypes via LD. Thus, we train penalized regression models on the subset of individuals with pericentromeric SNPs (from short-read panels) as predictors and centromeric SVs (called from long-read assemblies) as the response variable, and apply the trained model to predict centromeric SV dosages in other cohorts with available SNP genotype data.

Here, we apply this framework across 23 human chromosomes (chr1–22 and X), training imputation models on 188 HPRC genomes and imputing centromeric SV dosages into cohorts with expression, splicing, and clinical data (Figure 1). With directly genotyped SVs, we test associations with genome-wide CpG methylation and with the position of the CENP-A-associated hypomethylated dip within the active satellite array. We also uncover associations with cis and distal expression and splicing, including a gene implicated in heterochromatin formation. Finally, at biobank scale in All of Us, we assess the association between centromeric variation and complex traits, finding numerous links that are unexplained by SNPs alone.

**Figure 1:**
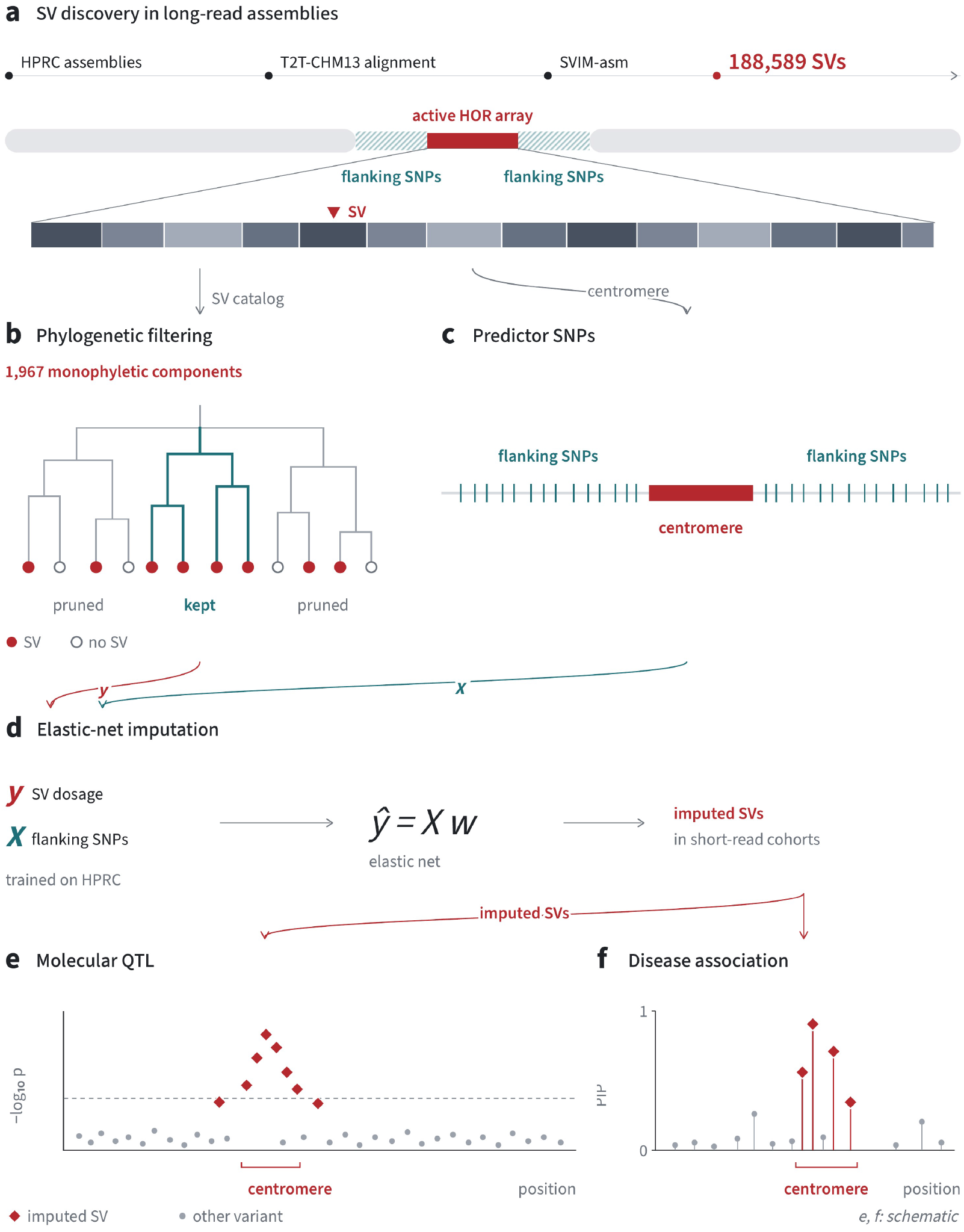
Centromeric structural-variant discovery, imputation and association with gene expression and disease. **a** Variant calling. Haplotype-resolved HPRC long-read assemblies were aligned to the telomere-to-telomere CHM13 reference, and structural variants were called within the active *α*-satellite higher-order-repeat (HOR) core with SVIM-asm, resulting in 188,589 centromeric SVs across chromosomes 1–22 and X. **b** The 1,967 monophyletic components resolved from complex multi-allelic SVs by per-chromosome neighbor-joining of centromeric haplotypes. **c** Pericentromeric predictor SNP sets, defined by three strategies (fixed flanking interval; boundary-anchored count; midpoint-anchored count). SNPs were extracted from the pericentromeric interval flanking each SV under each strategy, and the model with the highest cross-validated *R*^2^ among eight candidates was retained for that SV. **d** Per-SV elastic-net imputation into 630 unrelated 1000 Genomes samples. Models were fitted on the 188 HPRC individuals with matched SNPs under 4-fold cross-validation, and imputed SV dosage was used for the samples with matched MAGE expression and splicing. **e** Imputed SVs were tested as cis- and trans-QTLs for gene expression (eQTL) and RNA splicing (sQTL). **f** Imputed SV dosage was tested for disease association in All of Us participants and fine-mapped against the highly-correlated and tagged SNPs of each SV under in-sample LD.

## 2 Results

### 2.1 Centromeric SVs are abundant across 196 human genomes

Human centromeres include megabase-scale *α*-satellite higher-order repeat (HOR) arrays (0.34 Mb on chromosome 21 to 4.97 Mb on chromosome 18 in CHM13). Because 150-bp reads align ambiguously within these repetitive arrays, short-read SV catalogs and genotyping resources have typically excluded the centromere [Sudmant et al., 2015, Audano et al., 2019, Zook et al., 2020, Collins et al., 2020, Du et al., 2022, Ebler et al., 2022, Bai et al., 2026], whereas long-read assemblies have resolved complete centromeres [Logsdon et al., 2024, 2025]. To characterize structural variation in the centromere, we analyzed whole-genome haplotype-resolved assemblies (QV *≈* 55) from 196 individuals of the Human Pangenome Reference Consortium (HPRC) Release 2 (Figure 2a). We aligned each haplotype to the CHM13 v2.0 T2T reference [Nurk et al., 2022] with minimap2 [Li, 2018]. We then called structural variants per haplotype with SVIM-asm [Heller and Vingron, 2021] in haploid mode, keeping calls inside the active *α*-satellite HOR arrays annotated by CenSat v2.1 [Altemose et al., 2022] and summing the two haplotypes into diploid genotypes (Methods). We assessed the quality and consistency of the calls across SVIM-asm settings, their yield relative to PAV [Ebert et al., 2021] and dipcall [Li et al., 2018a] within active HOR arrays, their reproducibility between two assemblies of the same individual (HG002), and the assembly contiguity required to span an HOR array (Supplementary Notes 1–2).

**Figure 2:**
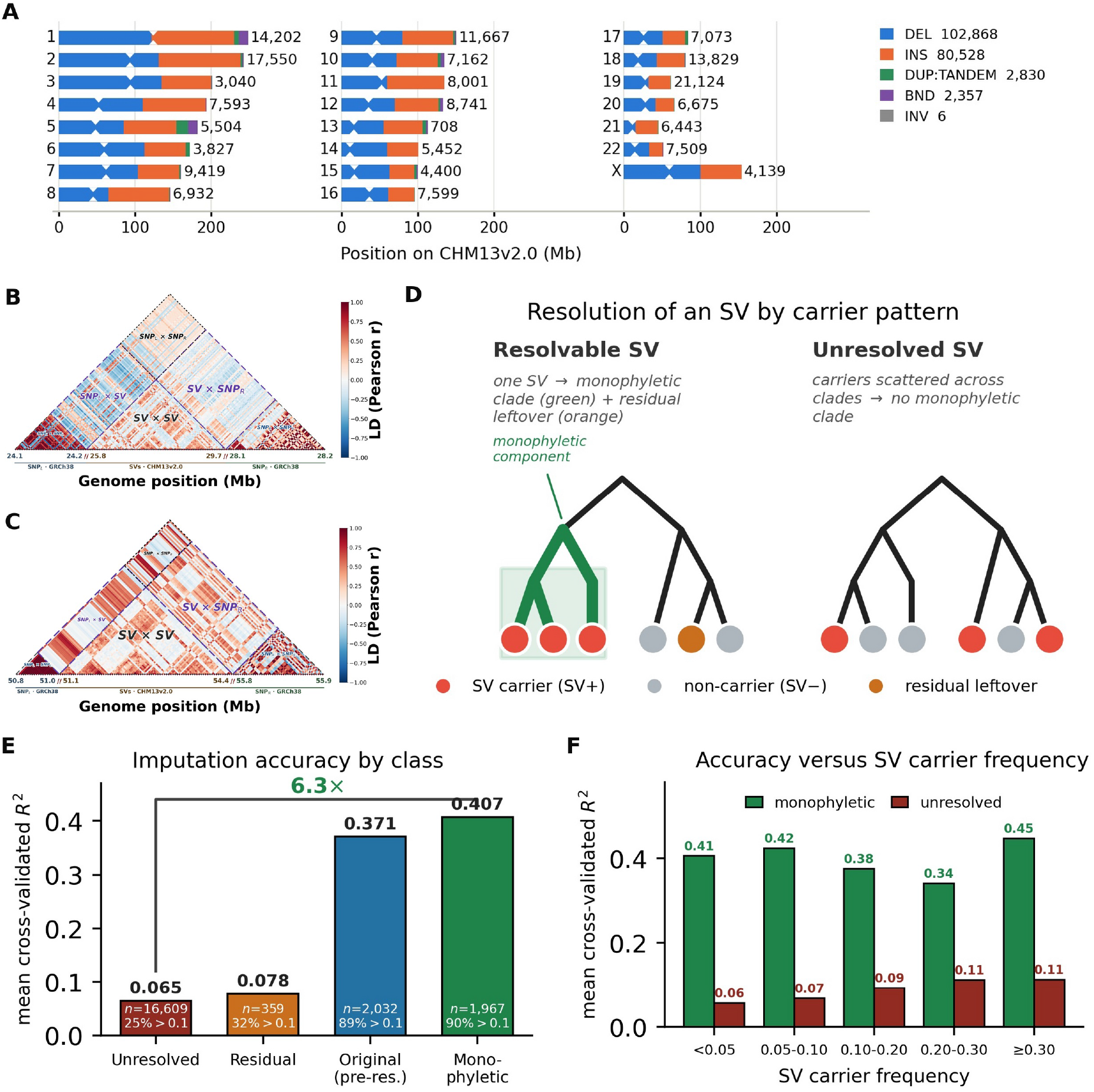
Centromeric structural variants in the active satellite array are linked to pericentromeric SNPs, and phylogenetic resolution improves SV imputation. **a** Each bar shows one chromosome at true length on a shared CHM13 v2.0 axis, filled by its SV-type composition and labeled with its total SV count. Notches mark the centromere, drawn wider than the arrays themselves (0.34–4.97 Mb on chromosomes of 45–248 Mb) so that they remain visible. **b, c** Pairwise LD (Pearson *r*) among pericentromeric SNPs and centromeric SVs across the chromosome-19 (b) and chromosome-11 (c) centromeres in the 188 individuals with assembled SVs and 1000 Genomes genotypes. Blocks were thinned; the SVs shown are the highest-frequency variants and the SNPs those most correlated. **d** Schematic of a *resolvable* SV, carriers forming a monophyletic clade (green), separated by resolution as an imputable component, off-clade carriers as a *residual*. An *unresolved* SV has carriers scattered across clades. **e** Mean cross-validated *R*^2^ by class (eight-model ensemble, 4-fold CV). Monophyletic components (*n* = 1,967) reach 0.407 compared with 0.371 for the same SVs before resolution (*n* = 2,032); unresolved SVs (*n* = 16,609) 0.065; residual components (*n* = 359) 0.078. **f** Advantage of monophyletic components over their own *higher-frequency* pre-resolution parents and over unresolved SVs in every carrier-frequency bin. All pericentromeric SV–SNP LD and monophyletic SV trees can be viewed at https://yikaidong-git.github.io/CenSV.

Across the autosomes and chromosome X, we called 188,589 centromeric SVs. Deletions (DEL; 102,868) and insertions (INS; 80,528) account for most (*∼*97%) calls, with tandem duplications (DUP:TANDEM; 2,830), breakends (BND; 2,357), and inversions (INV; *n* = 6) making up the remainder. The number of SVs varied considerably across chromosomes, ranging from 708 on chromosome 13 to 21,124 on chromosome 19. Of all calls, 96.5% involved only *α*-satellite-rich sequence, and the lengths of 84% of *α*-satellite-rich insertions and deletions lie within 10 bp of a multiple of the 171-bp monomer (Supplementary Note 13).

To determine which structural variants can be resolved within repetitive centromeric arrays, we introduced 1,017 SVs of known sequence into simulated CHM13 centromeres and applied our variant calling process (Supplementary Note 13). The simulation allowed us to quantify recall and precision across SV types, frequencies, and chromosomal origin. Recall required exact reconstruction of the altered sequence, allowing equivalent breakpoint placements and split calls. Recall was highest for insertions, deletions and duplications up to 1 kb (88%) and lowest for the largest SVs (18%). Large SVs were often represented as multiple smaller, incorrect calls; overall, 29% of reported calls contributed to an exact reconstruction, with substantial variation among chromosomes. Restricting calls to minor allele frequency *≥*5%, raised precision from 30% to 53%. These simulations show that our variant calling process is well-powered for small, simple SVs in haplotypes with limited sequence divergence from CHM13. Given the somewhat high false discovery rates, especially in chromosomes with greater sequence identity, we emphasize that our reported SVs should be considered candidate SV calls.

This catalog reveals extensive structural variation within regions that have been largely absent from genetic association studies. Direct genotypes for these variants come from the relatively small number of individuals with complete long-read assemblies. Suppressed recombination around centromeres creates long SNP haplotypes that can serve as proxies for otherwise inaccessible centromeric SVs. We therefore developed models to impute SV genotypes from pericentromeric SNPs.

### 2.2 Most centromeric SVs resolved to a single lineage are imputable from flanking SNPs

Recurrent mutation poses a particular challenge for imputation because the same apparent SV can occur on multiple flanking-SNP backgrounds. We therefore resolved complex SVs by centromeric haplotype lineage before prediction. We modeled the relationship between each SV and its surrounding SNPs using elastic-net regression. This provides a simple model for a region with little recombination: correlated SNPs collectively mark the SV across an extended interval, and regularization reduces them to a sparse set of informative predictors. The resulting models are interpretable, computationally efficient, and can be applied to cohorts with standard SNP genotype data without requiring a phased reference panel. For each SV, we selected the best of eight candidate elastic-net models and measured its accuracy using cross-validated *R*^2^ (CV *R*^2^; Eq. 5; Methods).

The 16,609 unresolved SVs reached a mean CV *R*^2^ of 0.065, with 25.0% reaching the CV *R*^2^ *≥* 0.1 threshold used for downstream association testing. Haplogroup resolution separated complex, multi-allelic SVs into monophyletic components, each explained by a single branch of the centromere haplotype phylogeny (Methods; Supplementary Fig. S1, Supplementary Table S4). Our resolution approach identified 135 of 2,103 resolved monophyletic SVs that had arisen independently on multiple haplotype backgrounds. We treated each origin as a separate monophyletic component and trained an independent prediction model for 1,967 of the 2,247 components, each with at least ten carriers. Overall, 1,967 monophyletic components reached a mean CV *R*^2^ of 0.407, with 90.4% reaching CV *R*^2^ *≥* 0.1 (Figure 2; Table 1).

**Table 1:** Imputation accuracy by haplogroup-resolution class. (eight-model ensemble, cross-validated *R*^2^). Resolving complex SVs into monophyletic haplogroup components (*n* = 1,967) gives a mean imputation accuracy of 0.407, compared with 0.371 for the 2,032 parent SVs. Unresolved SVs have much lower accuracy (0.065), reflecting an absence of single-branch structure.

| SV class | *n* | Mean CV *R*<sup>2</sup> | % *R*<sup>2</sup> ≥ 0.1 | % *R*<sup>2</sup> ≥ 0.3 |
|---|---|---|---|---|
| Unresolved | 16,609 | 0.065 | 25.0 | 3.5 |
| Residual | 359 | 0.078 | 32.3 | 2.8 |
| Original (pre-resolution) | 2,032 | 0.371 | 89.4 | 60.2 |
| Monophyletic | 1,967 | 0.407 | 90.4 | 64.4 |

This contrast indicates that recurrent mutation makes centromeric SVs much harder to impute. Once an SV was assigned to a single haplotype lineage, a small number of SNPs was often sufficient to predict its genotype accurately.

Imputation accuracy nevertheless varied substantially among monophyletic SVs. Beyond mutation recurrence, the strongest determinants were locus mapping quality, homozygous-alternate carrier count, active HOR span, and SV length. Locus mapping quality was measured by the mean MAPQ and the fraction of secondary, ambiguously mapping contig alignments overlapping the SV. Allele frequency and genome-composition features (GC content, satellite complexity, ancestry allele frequency) contributed more modestly (Methods; Supplementary Note 3).

### 2.3 Centromeric structural variants are associated with CpG hypomethylation within 5 Mb

We first examined molecular effects within the centromere itself, using haplotype-resolved CpG methylation to test whether centromeric SVs are associated with the chromatin state of the surrounding satellite array. We combined haplotype-resolved CpG methylation with directly genotyped centromeric SVs in 124 individuals from the 1000 Genomes ONT cohort [Gustafson et al., 2024]. Across 68 SVs on 19 autosomes, we identified 418 same-chromosome SV–CpG associations at global pair-level Benjamini–Hochberg *q <* 0.01, involving 42 SVs and 376 CpGs, which cluster into 16 genomic sites at 50 kb resolution (Figure 3; Supplementary Figs. S12 and S14).

**Figure 3:**
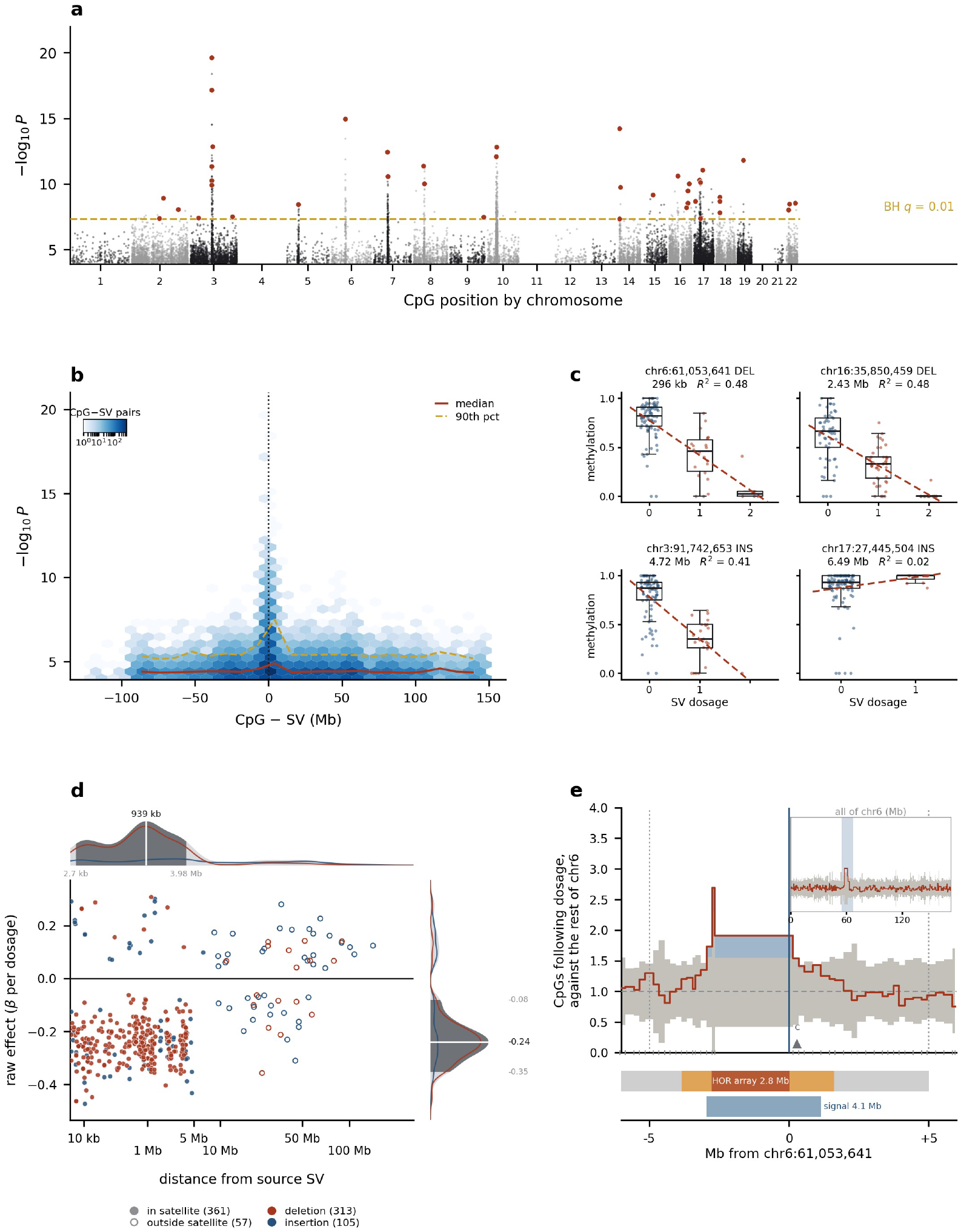
Centromeric SV dosage marks CpG hypomethylation in a local domain. **a** 82,365,801 cis tests between 68 directly genotyped SVs and same-chromosome CpGs in 124 individuals. 14,615 associations with *P ≤* 10*^−^*^4^ are shown, of which 418 reach *q <* 0.01. Red points mark the lead CpG for each of the 42 SVs contributing at least one significant association. Chromosomes 4, 11, and 20 had no retained SV. **b** Association strength and CpG-to-SV distance for the same 14,615 associations. Hexagonal bins show the number of CpG– SV pairs. Association strength rises sharply within 5 Mb of the SV. **c** Methylation against SV dosage at four targets, ordered by distance. **d** Raw effect against distance for the 418 discoveries. Most effects are concentrated at distances *<* 5 Mb and are negative. **e** Share of CpGs whose methylation is associated with chr6:61,053,641 deletion dosage, in 31 carriers and 93 non-carriers, compared with the same share beyond 20 Mb on chromosome 6. Gray marks the envelope of 60 permuted dosage vectors; shading marks the 3.8 Mb SHED, where the share lies above it, blue where carriers are less methylated. Bars beneath show the annotated centromere, its HOR array and the signal extent. The inset shows all of chromosome 6, and the triangle marks the CpG in (c). Supplementary Fig. S10 applies the same construction to all eight variants.

These associated CpGs were concentrated near the centromeric SVs. Of the 418 associated pairs, 360 lay within 5 Mb of one another, an 18.8-fold excess over permutation (Methods), whereas associations separated by more than 25 Mb were depleted. The local effects also differed by SV type. Among pairs within 5 Mb, increasing deletion dosage was associated with lower methylation in 97.3% of cases, including 18 of 20 deletion variants. Insertions showed a weaker directional bias despite having effects of similar raw magnitude. Centromeric deletions were thus associated with local hypomethylation. Along the chromosome, we identified contiguous regions near centromeric SVs that were enriched for CpGs whose methylation varied with SV dosage, compared with distal regions of the same chromosome. Because these local associations predominantly linked higher deletion dosage to lower methylation, we term these regions structural-variant-associated hypomethylation-enriched domains (SHEDs); they spanned 0.6 to 4.4 Mb at five of eight variants (Figure 3e; Supplementary Fig. S10). All 360 associations within 5 Mb lie in the CenSat-annotated centromeric region, across the *α*-satellite array and the adjacent satellite and transition sequence, and one domain reaches the pericentromeric flank (Supplementary Fig. S12e,f). Local CpG methylation distinguished deletion carriers from non-carriers in held-out individuals at nine of ten deletion sites (Supplementary Fig. S9), and CpGs sharing a source variant remain more co-methylated than matched background CpGs after adjusting for its dosage (Supplementary Fig. S13). Within SHEDs, increasing deletion dosage was generally associated with lower CpG methylation, with an effect magnitude that was approximately uniform within about 5 Mb.

### 2.4 A deletion on chromosome 8 is associated with the position of the likely kinetochore assembly site

This spatially bounded relationship motivated analysis of the centromere dip region (CDR), the hypomethylated interval overlapping CENP-A chromatin [Logsdon et al., 2021, Altemose et al., 2022]. We tested whether centromeric sequence variation was associated with CDR position. Most active arrays contain a single CDR. Merging dips within 150 kb gave a mean of 1.07 annotated CDRs per active array (93.1% with one; Supplementary Notes 7, 11).

We tested 11,129 centromeric SV loci against eight phenotypes describing the position, abundance, geometry, and methylation of the CDR (Methods). After correcting for all locus– phenotype tests, a single region on chromosome 8 was robustly associated with CDR position. The strongest association involved a 113-bp deletion at chr8:44,300,221 (*t* = *−*6.17, *P* = 1.0 *×* 10*^−^*^7^; allele frequency 0.55; *n* = 151; each copy associated with a haplotype-averaged CDR position approximately 300 kb nearer the array center). All study-wide-significant chromosome 8 associations fell within a 213.6-kb interval at chr8:44.25–44.46 Mb (Figure 4a,b; Extended Data Table 1). Multiple SV genotypes within this interval were associated with both measures of CDR position, indicating that they reflect a single chromosome 8 association signal (Supplementary Note 7).

**Figure 4:**
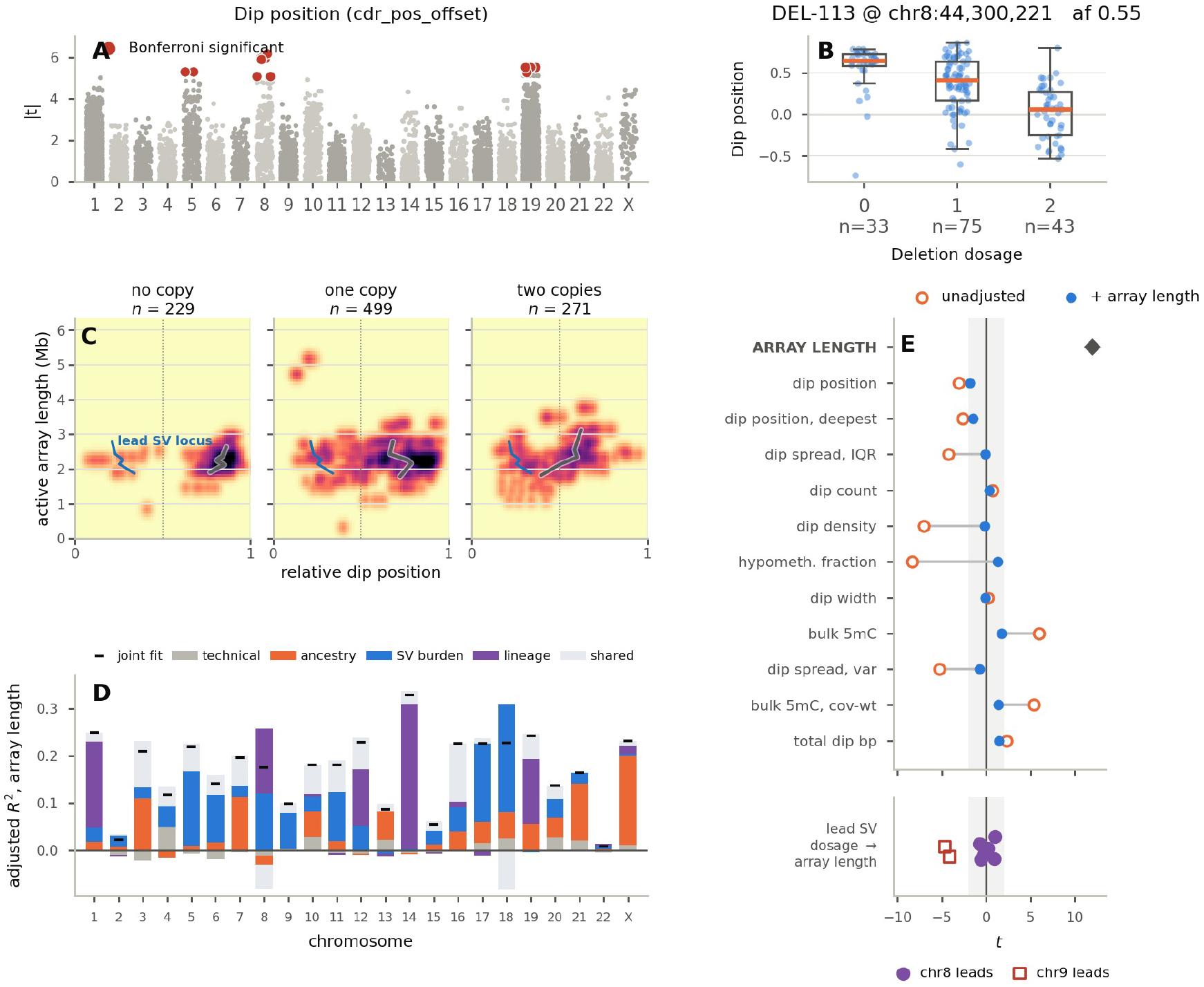
Centromeric SVs are associated with the position of the centromere dip region, the likely kinetochore assembly site. **a** Association of each of the 11,129 centromeric SV loci with dip position across all 23 chromosomes; *t* measures the strength of evidence for association. Leads arise on chromosomes 5, 8, 9 and 19 (Results). **b** The lead 113-bp deletion at chr8:44,300,221 is associated with dip position (*t* = 6.17, *n* = 151); each copy is associated with a haplotype-averaged dip position approximately 300 kb nearer the array center. Boxes show the median and interquartile range, whiskers 1.5 IQR. **c** Dip position along the chromosome-8 array against array length, split by dosage of the lead deletion. Relative position runs from 0 at the array start to 1 at its end. The gray line is the median dip position within each length bin. The blue line marks the lead deletion’s locus. Dip position shifts with deletion dosage, and array length is similar across dosage groups (Extended Data Fig. 5). For array-dip heatmaps of all 24 chromosomes, see Extended Data Fig. 4. **d** Variance in array length, the covariate adjusted in (e), explained by genomic features on each chromosome. Bars partition the joint *R*^2^ into the unique contribution of each predictor block and their shared contribution. **e** SV-burden association *t*-statistics for each phenotype before (open) and after (filled) adjustment for array length. The chromosome-8 association remains significant under this adjustment, and array expansion and ancestry background are candidate explanations for the leads on chromosomes 5, 9 and 19. The association between array length and dip position occurs across chromosomes, and dip position itself shows substantial haplotype-level variation, as shown in Supplementary Fig. S17; the homolog comparison and the scope of tag-SNP attribution are in Extended Data Fig. 6.

To characterize the locus at the sequence level, we decomposed the chromosome 8 active array of the CHM13 reference (chr8:44,243,546–46,325,080, 2.08 Mb) into *α*-satellite monomers, which HORmon assigns to seven major sequence classes (M1 to M7) by similarity to per-class consensus sequences (Methods). HORmon assigned the lead deletion as a 113 bp monomer of class M5 lying immediately adjacent to a full-length M5, and StringDecomposer assigned the same class to this sequence under a different partition of the region. The fragment matches the M5 consensus prefix more closely than any of 927 non-M5 monomers of this array (*P* = 0.001). Three nearby copies of this arrangement occur within 26.6 kb, two of which contain a CENP-B box variant with adenine at the first position, which occurs in four of the 5,263 boxes of this array. Three of the four monomers below 150 bp on this array fall in class M5 (Extended Data Fig. 7, Supplementary Note 7).

CDR position behaves as a stable property of a centromeric haplotype; dips were tightly clustered within each haplotype-chromosome but differed substantially among individuals. On chromosomes 6, 10, 12, 19 and X the dips resolve into two preferred positions, indicating that the kinetochore assembly site has more than one stable location along an array (Supplementary Table S10). The median within-haplotype standard deviation in relative dip position was 0.061, compared with 0.373 among individuals, a 6.1-fold difference (6.0-fold on chromosome 8). The chromosome 8 SVs therefore identify arrays on which functional centromeric chromatin occupies reproducibly different positions (Figure 4c).

We also found significant SV-CDR associations on chromosomes 5, 9 and 19. Because both SV discovery and several CDR phenotypes depend on HOR array size and position, we next tested whether variation in array coordinates explains these associations. Total SV burden strongly tracked array length (*t* = 11.95), and array length accounted for associations with phenotypes such as the hypomethylated fraction, dip density, and dip spread. For the chromosome 5, 9 and 19 leads, array expansion and ancestry background are candidate explanations; the chromosome 8 association remains significant after adjustment for array length (Extended Data Fig. 1, Supplementary Note 7). Centromeric structural variation on chromosome 8 is therefore associated with the position of the likely kinetochore assembly site.

### 2.5 Imputed centromeric SVs extend association testing to 630 individuals with expression data

We next extended the analysis beyond centromeric chromatin to gene expression and splicing. We imputed SVs with CV *R*^2^ *≥* 0.1 into 630 unrelated 1000 Genomes individuals with matched multi-ancestry gene expression (MAGE) data [Taylor et al., 2024] (Methods). After filtering to common SVs (frequency in [0.05, 0.99]), we tested imputed SV dosage for association with splicing and with gene expression in *cis* and *trans* (Methods).

### 2.6 Centromeric SVs associate with expression or splicing of 141 genes, including 13 distal genes on the same chromosome

We analyzed genes on the same chromosome separately from genes on other chromosomes and distinguished proximal (*±*10 Mb) from more distal same-chromosome genes.

Centromeric SVs were associated with 86 eGenes and 74 sGenes in cis (each SV’s own chromosome; Methods) at *q <* 0.05 (141 unique genes; Figure 5a). Fourteen of these (3 eGenes and 11 sGenes) were undetected by the MAGE consortium’s own SNP-based cis-QTL analysis at *q <* 0.05. Two further definitions of centromeric variation, short-read k-mer haplogroups (ascairn [Shiraishi et al., 2026]; 728 individuals) and *α*-satellite higher-order repeat variants called in the same assemblies (StV [Altemose et al., 2022]; the same 630 individuals), identified cis-eGene and cis-sGene sets of which two-thirds or more fell within ours (Supplementary Note 5).

**Figure 5:**
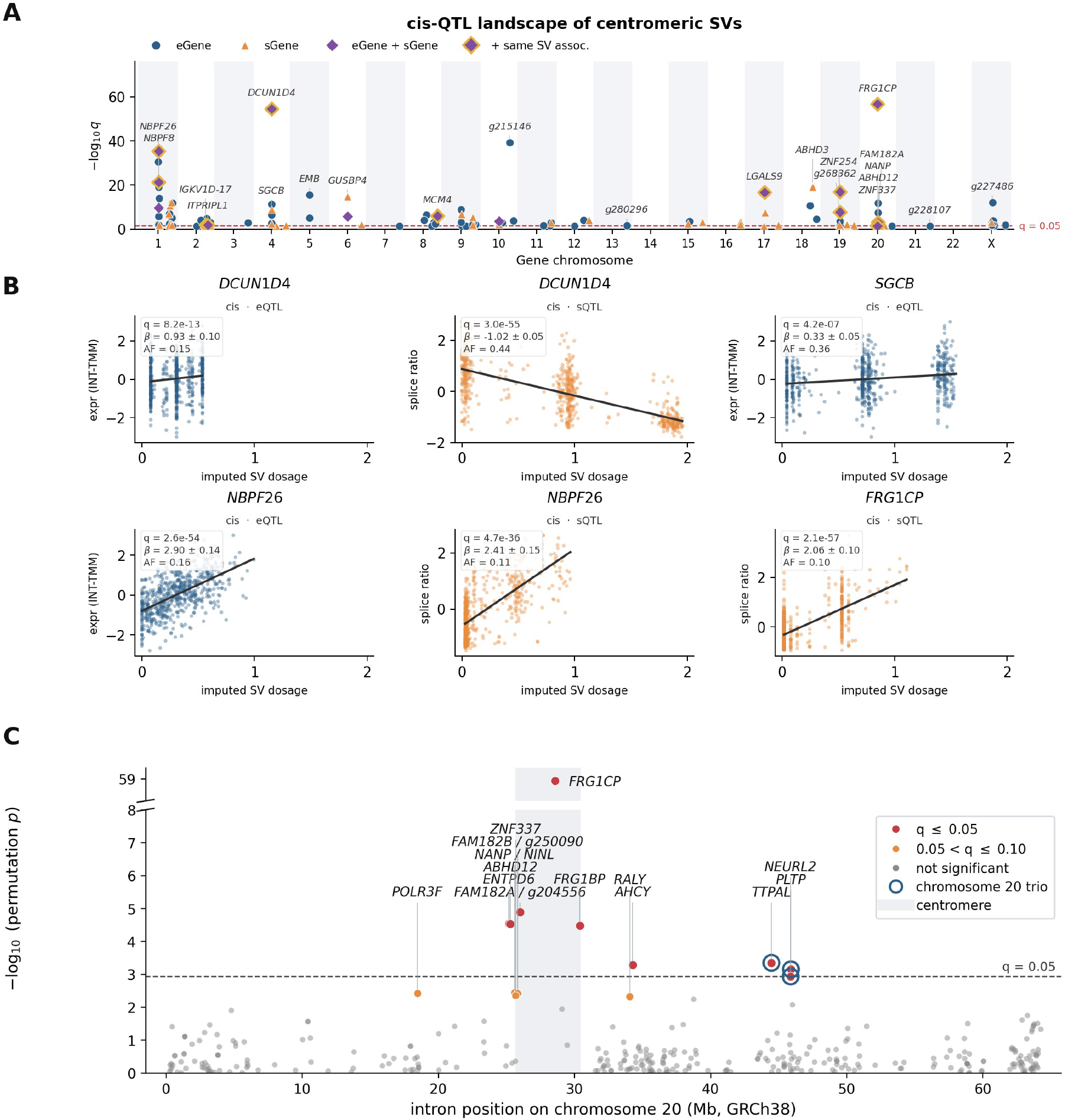
Imputed centromeric SV dosage is associated with expression or splicing of 141 genes. **a** *Cis* associations at *q <* 0.05: 86 eGenes (circles), 74 sGenes (triangles), and genes significant for both (diamonds), positioned by genomic location within the cis region (10 Mb from the centromere and the remainder of that chromosome). Gold outlines mark the 14 of 19 dual-modality genes whose expression and splicing associations share the same significant SV. Genes without an HGNC symbol are labeled *g* followed by the last six digits of their Ensembl accession (prefix: ENSG00000). **b** Imputed SV dosage against the molecular phenotype for six representative associations, each shown with a linear fit, the permutation *q* value for that modality, the effect size *β* (s.e.) per allele of imputed SV dosage from the cis-QTL model, and allele frequency. Imputed dosages show genotype-like segregation. **c** Splicing associations across chromosome 20. *TTPAL*, *PLTP*, and *NEURL2* lie 14–16 Mb from the centromere (Supplementary Note 6). AF, allele frequency; eGene, gene with a significant expression association; sGene, gene with a significant splicing association.

We observed that a 2,057-bp insertion within the active chromosome X *α*-satellite array was associated with expression of *SUV39H1* (cis-eQTL *q* = 1.5 *×* 10*^−^*^2^; CV *R*^2^ = 0.17). Notably, *SUV39H1* encodes an H3K9 methyltransferase that helps establish and maintain centromeric heterochromatin [Rea et al., 2000, Peters et al., 2001, Ma et al., 2024, Carty et al., 2026, Sidhwani et al., 2025]. This association indicates a connection between structural variation within an active centromeric array and the expression of a key regulator of centromeric heterochromatin.

Several associations extended far beyond the pericentromeric region. Thirteen genes at 10 loci lay at least 10 Mb from both the centromere and the nearest tag SNP, at distances of 12.2–132.4 Mb, and all of these associations persisted after conditioning on the local tag SNPs (Supplementary Note 6). On chromosome 20, distinct centromeric SVs were associated with splicing of *TTPAL*, *PLTP* and *NEURL2*, 14–16 Mb from the centromere and 13–15 Mb from the nearest tag SNP (cis-sQTL *q* = 3.1 *×* 10*^−^*^3^, 2.7 *×* 10*^−^*^2^ and 3.9 *×* 10*^−^*^2^, respectively; Figure 5c). In contrast, zero *trans* associations survived multiple-testing and paralog-mismapping controls (Supplementary Fig. S6; Supplementary Note 6). Thus, the associations detected here were confined to the same chromosome, with some extending far beyond the *±*10 Mb region.

In summary, common structural variation in the centromere associates with distal gene expression and splicing on the same chromosomes, and these associations persisted after conditioning on the tag SNPs used to impute the SVs (Supplementary Note 6).

### 2.7 Imputed centromeric SVs associate with 27 clinical phenotypes in All of Us

Imputation also makes centromeric variation accessible to phenome-wide analysis at biobank scale.

We imputed centromeric SV dosages into All of Us [All of Us Research Program Genomics Investigators, 2024] using WGS-derived tag SNP genotypes and tested them across both case-control and quantitative phenotypes. We scored 10,799 of the 11,129 SV predictors and tested them against 2,417 phecodeX [Shuey et al., 2023] definitions, for a total of 26.1 million SV– trait tests. Sample sizes ranged from 123,760 to 315,204 participants across traits (Methods). Association statistics were well calibrated across the phenome-wide analysis (*λ*_GC_ = 1.035), and all 7.5 million tests of a prespecified set of 3,097 negative-control SV predictors had *q ≥* 0.10 (Figure 6b; Methods).

**Figure 6:**
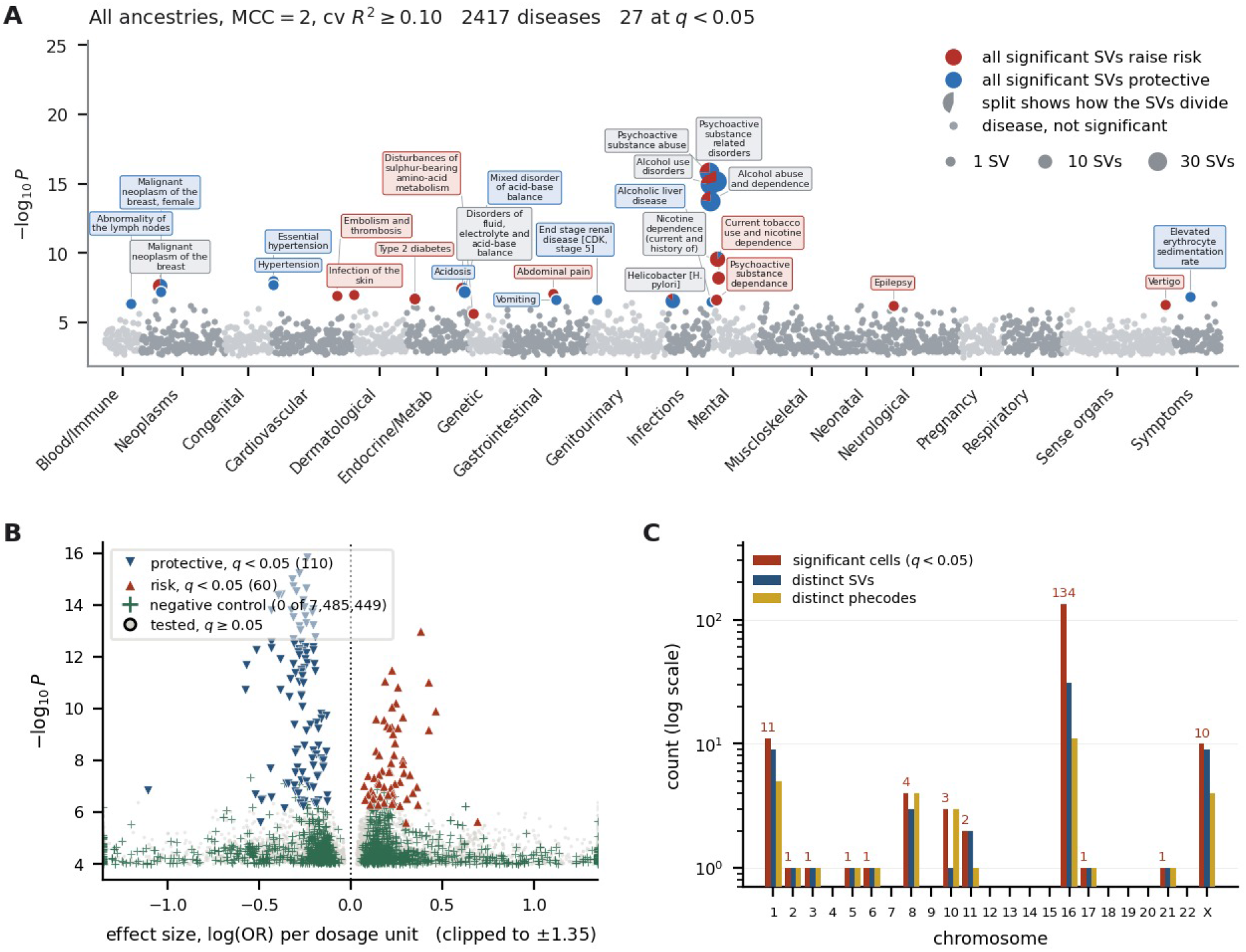
Phenome-wide associations of imputed centromeric SV dosage in All of Us. **a** Phenome-wide disease association analysis, 2,417 phecodeX definitions against 10,799 scored predictors (CV *R*^2^ *≥* 0.10), all ancestries (*λ*_GC_ = 1.035). Each point represents one phecode at its strongest predictor. The 27 clinical phenotypes at *q <* 0.05 are named, and drawn as circles whose area grows with the number of distinct SVs associated with that phenotype and which are divided red against blue in proportion to how those SVs split on direction of effect. **b** Effect size against significance for every SV–trait pair retained at *P ≤* 10*^−^*^4^; the 170 at *q <* 0.05 are 110 inverse and 60 positive associations. Green crosses are the 1,481 retained SV–trait pairs of the pre-selected negative controls; zero of 7,485,449 control tests reach *q <* 0.05. Axis clipped to *±*1.35. **c** Per chromosome, the number of significant SV–phecode associations, of distinct SVs in those associations, and of distinct phecodes.

We identified 170 SV–clinical phenotype associations (*q <* 0.05) involving 61 SV predictors and 27 clinical phenotypes, with another 85 associations at *q <* 0.10 (Figure 6a; Supplementary Note 8). The strongest association connected a chromosome-16 deletion to psychoactive substance-related disorders (OR = 0.79, *P* = 1.5 *×* 10*^−^*^16^; 26,943 cases). The same deletion was associated with two additional substance-use clinical phenotypes. Across all significant associations, odds ratios ranged from 0.33 to 2.00, with a median of 0.81.

The associations spanned 12 chromosomes. Chromosome 16 contributed 134 of the 170 significant SV–clinical phenotype associations, representing 31 correlated SV predictors and 11 clinical phenotypes (OR 0.56–1.59). Chromosome X contributed another 10 associations (OR 0.61–2.00), and the remaining 26 were distributed across ten autosomes (OR 0.33–1.44; Figure 6c). Outside the chromosome-16 cluster, signals included seven chromosome-1 SV predictors associated with breast cancer (OR 0.64–1.25; 9,124 cases) and associations with hypertension-related phenotypes (OR = 0.86; 103,924 cases; Figure 6a,c). Thus, the phenome-wide results nominate centromeric SVs and their linked variants across several domains of complex-trait biology.

The 3,097 negative-control SV predictors were well imputed and were selected for a null result in the summary-level joint fine-mapping (Supplementary Note 10; Methods). The smallest *P* value among their 7.5 million tests, 4.5 *×* 10*^−^*^8^, was consistent with the minimum expected by chance. The control set therefore supports the calibration of the overall analysis.

The chromosome-16 signal was concentrated in the AMR ancestry group. Of the 170 significant associations, 138 reached nominal significance in AMR, compared with 12 in EUR, two in AFR, and none in EAS, SAS, or MID (Extended Data Fig. 2a). This concentration arose although the EUR sample was approximately three times larger for these phenotypes. Chromosome 16 also has the highest mean *F_ST_* among imputable centromeric SVs (Supplementary Note 12). The chromosome-16 cluster may therefore reflect an ancestry-specific genetic association or other factors, such as ancestry-correlated differences in recruitment, healthcare use, or diagnostic coding.

Because imputed SVs are correlated with the SNPs used to predict them, disease associations cannot automatically be attributed to the SV itself. We therefore jointly fine-mapped each SV with nearby SNPs. A chromosome-1 deletion associated with disorders of fluid and electrolyte balance (*q* = 0.008) led its credible set at posterior probability 0.999 under all three fine-mapping methods and both variant-panel constructions. Across the 476 fits, an SV was included in a credible set in 69 and led that set in 34, 26 of them under at least two methods; a SNP had the larger statistic in 451 (median margin 1.90 in *|z|*; Extended Data Fig. 2b), consistent with most SVs sharing their association with linked SNPs.

Taken together, these results show that imputed centromeric SV dosages are associated with complex traits and diseases in large-scale populations.

## 3 Discussion

Human centromeres have long been excluded from population-scale genomics because *α*-satellite arrays are technically challenging to analyze. This has greatly inhibited study of genetic variation in the centromere, which plays an essential role in cell division. Here, we show that centromeric SVs are associated with the methylation of the surrounding satellite array, with the position of the likely kinetochore assembly site, with the expression and splicing of genes along the same chromosome, and with complex traits. The analyses of gene expression and complex traits were enabled by imputation. Because suppressed recombination preserves associations between satellite arrays and flanking SNPs, statistical models trained on the 188 long-read HPRC assemblies with matched SNP genotypes aligned to CHM13 can impute centromeric SV dosage from pericentromeric SNPs in genotyped cohorts with adequate pericentromeric SNP coverage. Imputation thereby brings these otherwise difficult-to-measure SVs into existing population cohorts with short-read sequencing or genotype arrays, extending previous work imputing short tandem repeats, variable-number tandem repeats, and structural variants from SNP haplotypes [Saini et al., 2018, Gymrek et al., 2016, Fotsing et al., 2019, Bakhtiari et al., 2021, Mukamel et al., 2021, Bai et al., 2026, Noyvert et al., 2025]. Complementary k-mer approaches infer centromere haplogroups or structural features directly from short reads [Shiraishi et al., 2026, Hain et al., 2026]. Our observations permit four conclusions.

First, centromeric deletions are strongly linked to hypomethylation domains (SHEDs) in proximal satellite sequence. Experimental depletion of DNA methylation alters CENP-A localization and chromosome stability [Salinas-Luypaert et al., 2025]. The methylation associations were concentrated near the tested SV, and among deletion–CpG pairs within 5 Mb, increasing deletion dosage was associated with lower methylation in nearly all cases. We note that three mechanisms would be consistent with this observation: 1. deletions may alter local methylation; 2. the deletion and the hypomethylation may both reflect linked variants or a broader property of the underlying haplotype; 3. methylation state may shape which deletions arise or persist.

Second, centromeric SVs are also associated with the position of the likely kinetochore assembly site. On chromosome 8, a 113-bp deletion and linked variants were associated with the position of the centromere dip region (CDR), the hypomethylated interval that overlaps CENP-A chromatin [Altemose et al., 2022]. The positional association was separable from array length, dip extent, and overall methylation, which suggests that the location of the functional centromere can vary independently of the amount of hypomethylated sequence. Recent population-scale studies using orthogonal approaches observed a similar result, connecting chromosome-8 centromere structure to CDR position [Sun et al., 2026, Gao et al., 2026]. Although the CDR-associated 113-bp deletion on chromosome 8 was well imputed, we found little evidence that it was associated with complex traits, consistent with common variation in CDR position on chromosome 8 being tolerated at the level of complex traits. However, the CDR is located preferentially toward one end of the active array on some chromosomes, which is compatible with a model of selection favoring biased kinetochore assembly site positioning.

Third, centromeric SVs are associated with gene expression and RNA splicing of genes on the same chromosome. Some involved nearby genes, whereas others involved genes far from the centromere and persisted after conditioning on the local tag SNPs, consistent with structural variants influencing the regulation of multiple genes [Scott et al., 2021]. An informative example linked a chromosome-X centromeric insertion to expression of *SUV39H1*, which encodes an H3K9 methyltransferase involved in establishing and maintaining pericentromeric heterochromatin [Rea et al., 2000, Peters et al., 2001]. Its recruitment to pericentromeric DNA and role in maintaining the boundary of CENP-A chromatin connect this association to centromere organization [Ma et al., 2024, Carty et al., 2026, Sidhwani et al., 2025]. Combined with the methylation associations, this result raises the possibility that centromeric SVs influence centromeric chromatin both through methylation of the surrounding satellite array and through expression of a gene whose product helps maintain centromeric heterochromatin. More distant associations, including the chromosome-20 splicing signals, suggest candidate mechanisms involving intermediate regulatory genes and long-range chromatin organization.

Fourth, beyond molecular phenotypes, centromeric SVs are associated with complex traits. In All of Us, associations with clinical phenotypes as diverse as substance-related disorders and breast cancer point to centromeric SVs as candidate contributors to complex-trait variation. These observations led us to ask: how is it that the centromere, which contains no protein-coding genes, might influence complex traits? We suspect that this contribution is driven primarily by proximal mechanisms, in which centromeric variation alters the methylation and heterochromatin landscape in pericentromeric regions, thus altering the regulation of pericentromeric protein coding genes in *cis*, which may in turn affect downstream genes in *trans*.

Four limitations of this work define directions for future study. First, high sequence identity within HOR arrays limits both the recall and precision of SV calling in centromeres. In simulations, recall was strong for insertions, deletions and duplications up to 1 kb and declined for larger changes and greater sequence divergence from CHM13. In contrast, large SVs were often represented as several smaller, incorrect calls. Precision varied substantially among chromosomes, indicating considerable heterogeneity in variant calling difficulty across chromosomes. Our SV catalog should therefore be interpreted as candidate SV calls whose sequence-level reliability depends on event type, size and chromosome context. An incorrectly defined SV can nevertheless mark a centromeric haplotype associated with a molecular or clinical phenotype, because errors in SV definition are highly unlikely to confound association testing with phenotypes. Second, monophyletic SVs are imputed far more accurately than non-monophyletic SVs, and the latter will require dedicated approaches to imputation and analysis. Third, experimental approaches will be needed to partition causality between the SVs and the SNPs in linkage disequilibrium with them. Finally, we imputed SVs with a regularized linear model, and classical HMM-based imputation algorithms and reference panels tuned for centromeric SVs may reach higher accuracy. Limitations in imputation are largely a function of panel size and will ease as long-read pangenome resources grow. The variation captured by complete-centromere assemblies is vast [Logsdon et al., 2024, Gao et al., 2026], and expanding haplotype-resolved cohorts [Liao et al., 2023] will deepen the resolution, extend imputation to the recurrent variants now beyond reach, and eventually power trait mapping.

Overall, we anticipate that our approach, which enables the characterization of SVs in short-read and genotyping-array-based cohorts, will facilitate the standard inclusion of centromeric SVs in future human genetic association studies.

## 4 Methods

### 4.1 Assembly alignment and structural-variant calling

We obtained haplotype-resolved assemblies for 196 HPRC Release 2 individuals [Lucas et al., 2026] and aligned each haplotype to the CHM13 v2.0 reference (chromosomes 1–22 and X from Nurk et al. [2022], chromosome Y from Rhie et al. [2023]) with minimap2 [Li, 2018] in assembly- to-reference mode (preset: `asm5`, output: `–cs`, chaining bandwidth for repetitive centromeric sequence: 2 kb, `-r2k`). We called structural variants genome-wide with SVIM-asm v1.0.3 [Heller and Vingron, 2021] in haploid mode (minimum mapping quality *≥* 20, minimum SV length 10 bp, maximum SV size 500,000 bp, and query and reference gap tolerance 200 bp); events exceeding the maximum SV size are reported as breakends. We then retained calls whose breakpoints fell within the active *α*-satellite HOR arrays defined by CHM13 v2.0 CenSat v2.1 annotations [Altemose et al., 2022], merged the per-haplotype callsets into a multi-sample cohort VCF, and summed the two haplotypes of each individual into diploid genotypes: 0 (homozygous reference), 1 (heterozygous), or 2 (homozygous alternate). We analyzed 23 chromosomes (chr1–22 and X) through an identical pipeline. Seven chromosomes (chr1, 5, 13, 14, 19, 21, and 22) contain HOR arrays with cross-chromosome *α*-satellite homology [Alexandrov et al., 2001, Shepelev et al., 2015] that can confound haplogroup resolution. We used HiFi-based HPRC assemblies (Supplementary Note 2), and we selected SVIM-asm over the assembly SV caller PAV [Ebert et al., 2021] after comparing both inside active HOR arrays of HG002 (Supplementary Note 1).

### 4.2 Simulation benchmark

We introduced SVs of known sequence into the active HOR arrays of CHM13 chromosomes and applied the same alignment and SV-calling pipeline; Supplementary Note 13 defines recall, precision and specificity.

### 4.3 Haplogroup resolution of centromeric SVs

We developed a per-chromosome phylogenetic approach to assign centromeric SVs to haplotype lineages.

Based on the phased, haplotype-resolved assembly calls, we built a binary presence/absence matrix of SVs across the 2*N* haplotypes of the *N* individuals. We computed pairwise cosine distances between haplotypes,

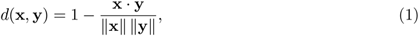

where **x**, **y** are binary SV vectors for two haplotypes, and built a neighbor-joining (NJ) tree from the distance matrix with scikit-bio [Saitou and Nei, 1987]. At each iteration the NJ algorithm joins the pair of nodes (*i, j*) that minimizes

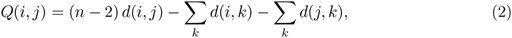

where *n* is the current number of nodes, proceeding until the tree is fully resolved. We midpoint-rooted the tree and used each internal branch to divide the haplotypes into two groups. For every SV with presence/absence vector **s** *∈ {*0, 1*}^H^* across the *H* haplotypes and each branch-partition indicator **b** *∈ {*0, 1*}^H^*, we computed the coefficient of determination

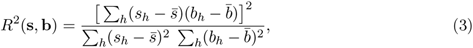

the squared Pearson correlation between the SV pattern and the bipartition. We classified an SV as *monophyletic* if its best-matching branch reached *R*^2^ *≥* 0.3 and shared at least ten carriers with that branch. We genotyped the component as the dosage of those shared carriers and imputed it independently. Because an SV may have multiple origins, we masked the matched carriers and repeated the search on the remaining off-branch carriers. We accepted another monophyletic component whenever a branch reached *R*^2^ *≥* 0.3 with at least five carriers and recorded any remaining unassignable carriers as a *residual* component. We left SVs unresolved if their first-pass component shared fewer than ten carriers with any branch or if no branch reached *R*^2^ *≥* 0.3. For each resolved SV, we retained the multi-allelic record before this procedure as its *original (pre-resolution)* record.

### 4.4 Pericentromeric SV–SNP linkage disequilibrium

We measured linkage disequilibrium (LD) between centromeric SVs and flanking SNPs. For each chromosome we assembled a combined panel of haplogroup-resolved centromeric SV genotypes and pericentromeric SNP genotypes in the 188 individuals with both, and computed the pairwise Pearson correlation *r* between every pair of panel variants. The active *α*-satellite HOR interval follows the CHM13 v2.0 CenSat annotation [Altemose et al., 2022]; we selected SNPs from the 1000 Genomes high-coverage phased VCFs [Byrska-Bishop et al., 2022] within 100 kb outward of each HOR edge and restricted the analysis to biallelic sites with minor allele frequency *≥* 0.05.

### 4.5 Pericentromeric SNP extraction

We extracted pericentromeric predictor SNPs from the 1000 Genomes Project high-coverage phased VCFs [Byrska-Bishop et al., 2022] for both the training and imputation-target samples. For training, we used the 188 HPRC individuals that overlap the 1000 Genomes panel and therefore have both directly assembled centromeric SVs (the response) and short-read SNP genotypes (the predictors). We built and integrated eight candidate models spanning three SNP-selection strategies:

- **Strategy 1 (fixed genomic interval):** SNPs within a fixed distance of 100, 200, or 500 kb outward from each centromere boundary. We defined centromere boundaries using the UCSC hg38 `cytoBand` [Furey and Haussler, 2003] annotation [Kent et al., 2002], merging each chromosome’s p- and q-arm `acen` bands. We manually adjusted the intervals for chr1, chr2, chr6 and chrX (by 0.7–3.0 Mb; Supplementary Table S13).
- **Strategy 2 (boundary-anchored SNP count):** the nearest 500, 1,000, or 2,000 SNPs on each side of the centromere boundary, searched up to 20 Mb outward.
- **Strategy 3 (midpoint-anchored SNP count):** the nearest 500 or 1,000 SNPs on each side of the GRCh38 centromere middle position.

Two ensembles integrated these candidates by retaining, per SV, the single model with the highest cross-validated *R*^2^: the best of six candidate models (Strategies 1 and 2, at three settings each, all anchored at the centromere boundary) and the best of eight (adding the two Strategy 3 models, anchored at the midpoint); we used the best-of-eight ensemble in the main analysis.

#### Heterochromatin-aware flanking

Several centromeres are adjacent to large, SNP-sparse pericentromeric heterochromatin blocks (e.g., 1q12, 7qh, 9q12, 11p). For the fixed-interval strategy (Strategy 1) we therefore adopted chromosome-specific boundaries: we extended a centromere edge outward to include an adjacent heterochromatin block when the gap between the block and the boundary was *≤* 1 Mb, before applying the flanking interval. We used unadjusted boundaries for the remaining chromosomes: 16 of 19 have no heterochromatin block within 1 Mb, and for the other three (chr2, chr16, chr21) we retained the unadjusted boundary.

We retained biallelic SNPs with minor allele frequency *≥* 0.05 and *<* 5% missing genotypes.

### 4.6 SV genotype prediction with an elastic-net model

For each SV we trained a separate elastic-net model predicting its genotype from the extracted pericentromeric SNP genotypes. The elastic net [Zou and Hastie, 2005] minimizes

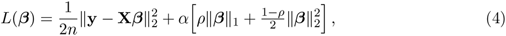

where **y** *∈ {*0, 1, 2*}^n^* is the SV genotype vector, **X** is the standardized SNP genotype matrix, *α* is the regularization strength, and *ρ* (the `l1_ratio`) balances the L1 (lasso) and L2 (ridge) penalties. We standardized SNP features to zero mean and unit variance (`StandardScaler`, scikit-learn [Pedregosa et al., 2011]); we used SV genotypes in raw integer form. The unpenalized intercept *β*_0_ used for prediction (Eq. 6) is fit separately by scikit-learn and is omitted from Eq. 4.

We selected hyperparameters by cross-validation (scikit-learn `ElasticNetCV`, `max_iter` = 2000) over *ρ ∈ {*0.3, 0.5, 1.0*}* and 50 automatically spaced *α* values per *ρ*, using *K* = 4 folds (`KFold`, shuffled, `random_state` = 42); we chose the combination minimizing mean cross-validated squared error (MSE). We filtered SVs to carrier frequency in [0.05, 0.99] before training, and predictor SNPs to carrier frequency *≥* 0.05.

The main quality metric was the cross-validated *R*^2^,

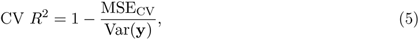

computed from held-out fold predictions against observed genotypes [Marchini and Howie, 2010, Das et al., 2016]. We trained models on the 188 HPRC individuals with both long-read-assembled centromeric SV genotypes and short-read-genotyped pericentromeric SNPs. We benchmarked the per-SV elastic net against a Beagle v5 reference-panel copying model on an assembly-cophased reference panel, with the identical shared SNP set, effective population size *n_e_* = 2000, and leave-one-out affine recalibration of Beagle’s posterior dosage (Supplementary Note 4).

### 4.7 Determinants of SV imputability

For each imputed SV we assembled per-SV features in four groups: intrinsic SV properties (length, carrier frequency, homozygous-alternate count, SV type), locus/cis-region properties (number and distance of available predictor SNPs), chromosome-level properties (active HOR span, number of HOR intervals, pericentromeric flank length, acrocentric status), and locus mapping-quality properties (mean MAPQ, fraction of secondary contig alignments, contig count, and edit distance at the SV locus). We related these to the best-of-eight ensemble cross-validated *R*^2^ and to the binary imputability thresholds (*R*^2^ *≥* 0.1 and *≥* 0.3). We fit three complementary models: (i) a univariate association analysis (Spearman rank correlation and, for binary outcomes, ROC-AUC); (ii) a linear mixed-effects model with fixed effects for the numeric features and a per-chromosome random intercept; and (iii) a gradient-boosted regression/classification model (HistGradientBoostingRegressor/Classifier) with feature importances estimated by permutation importance on a held-out test split. We reported model-output diagnostics (e.g., the number of nonzero coefficients) separately and excluded them from the predictor set. We also compared reproducibility of imputable and unimputable SVs in the Q100 HG002 v1.1 assembly (Supplementary Note 3).

### 4.8 SV dosage imputation into 1000 Genomes samples

We selected imputation target samples from the 1000 Genomes high-coverage cohort by excluding the HPRC training individuals, all members of their families, and non-founder (offspring) individuals, then intersecting with the samples with matched MAGE expression data, leaving 630 unrelated samples (AFR 159, EUR 135, EAS 127, SAS 118, AMR 91). For each chromosome, we extracted SNP genotypes at the exact training positions from the 1000 Genomes phased VCFs [Byrska-Bishop et al., 2022]. Matching required identical chromosome, position, reference, and alternate alleles; we assigned absent or allele-mismatched target SNPs the homozygous-reference genotype (0) before standardization. We scaled target SNP genotypes with the training-fitted StandardScaler and predicted SV dosages as

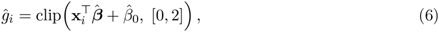

We retained SVs with CV *R*^2^ *≥* 0.1 and further required SV carrier frequency in [0.05, 0.99] among the 630 samples before QTL association testing.

### 4.9 Cis- and trans-QTL association testing

We obtained expression and splicing phenotypes from MAGE v1.0 [Taylor et al., 2024], lymphoblastoid cell-line RNA-seq of 1000 Genomes individuals (inverse-normal-transformed, TMM-normalized expression for 20,154 genes; LeafCutter [Li et al., 2018b] intron-excision ratios for splicing). Association testing used tensorQTL [Taylor-Weiner et al., 2019]. For each SV–gene pair we fit the linear model

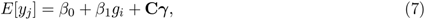

where *y_j_* is the normalized phenotype, *g_i_* the imputed SV dosage, and **C** the covariates (sex, 5 genotype PCs, and 60 PEER factors [Stegle et al., 2012] for expression or 15 for splicing). We treated genes on the SV’s own chromosome as cis (covering both the conventional *±*10 Mb region around the centromere and the remainder of that chromosome) and genes on all other chromosomes as trans. For cis genes, per-gene significance used 10,000 permutations with a Beta-distribution approximation [Ongen et al., 2016], and cross-gene correction used *q* values [Storey and Tibshirani, 2003] (*q <* 0.05). In trans, we tested each centromeric SV against genes and splicing phenotypes on all other chromosomes. We retained pairs at nominal *P <* 10*^−^*^5^ and declared trans genes significant at a genome-wide threshold of *P <* 5 *×* 10*^−^*^8^. We included SVs with CV *R*^2^ *≥* 0.1 and carrier frequency in [0.05, 0.99]. We further assessed trans associations using Benjamini–Hochberg and Bonferroni correction over the full number of SV–phenotype pairs tested, cross-mappability between each target gene and the pericentromere of the SV’s own chromosome [Saha and Battle, 2018a], conditioning on the tag SNPs of the SV’s imputation model, and a search of the complete CHM13 assembly (`minimap2 -x asm20 -N 50`) for paralogs of the target locus that GRCh38 omits; the same paralog search was applied to distal cis genes against the pericentromere of their own chromosome. We assessed calibration using genomic-control *λ* for the per-gene Beta-approximated permutation *P* values (Supplementary Note 6).

To identify genes detected only by the SV analysis, we compared significant SV-associated genes with the MAGE consortium’s SNP-based cis-QTL results. We classified genes significant in our analysis and absent from the MAGE set of genes with a significant SNP-based cis-QTL (*q <* 0.05) as novel.

We also evaluated two alternative genotyping methods with the same QTL mapping (Supplementary Note 5). StV [Altemose et al., 2022] defines variants from *α*-satellite higher-order repeat monomer organization using HumAS-HMMER HOR monomer annotations. ascairn [Shiraishi et al., 2026] infers centromere haplogroups from short-read k-mer content using their published panel of assembled active-HOR haplotypes. We imputed StV dosages using the same procedure and tested both approaches with the main association-testing framework.

### 4.10 Methylation-QTL association testing

#### Methylation and genotype data

Haplotype-resolved whole-genome CpG methylation (5mC bedMethyl) from the 1000 Genomes ONT Sequencing Consortium data release (`s3://1000g-ont/PROCESSED_DATA/ALIGNED_TO_HG38/MODKIT/`) [Gustafson et al., 2024] provided the phenotype. We retained CpGs with *≥* 10*×* coverage and a methylation call in at least 90% of analyzed samples. We genotyped the SVIM-asm catalog sites (see Assembly alignment and structural-variant calling) using CHM13-aligned Nanopore long reads with Sniffles2 v2.7.1 [Smolka et al., 2024]. Of the 176 individuals with methylation data, we included 124 with corresponding long-read genotype data. We imputed missing SV genotypes using the cohort mean dosage for the corresponding SV. We selected catalog SVs for read genotyping if they had *q <* 0.01 in the imputed-dosage association analysis or a SuSiE [Wang et al., 2020] inclusion probability *≥* 0.8. We included genotyped SVs in the association analysis if they had a depth-qualified call rate *≥* 0.8, an observed allele frequency *≥* 0.01, at least three observed carriers, and a minor allele count *≥* 8. This retained 68 SVs on 19 autosomes, including 37 deletions and 31 insertions.

#### Genome-wide association testing and multiple-testing correction

We fit each SV– CpG pair by ordinary least squares:

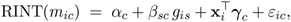

where *m_ic_* is the methylation fraction of individual *i* at CpG *c*, *g_is_ ∈* [0, 2] the alternate-allele dosage of SV *s*, RINT(*·*) the rank-inverse-normal transform applied across individuals within each CpG, and **x***_i_* a covariate vector of sex, five genotype principal components and ten methylation principal components. We tested a pair only when its complete-case minor-allele count was at least 8. Standard errors are heteroskedasticity-consistent (HC3).

We tested associations using RINT-transformed methylation values and estimated effect sizes from the same model fitted to untransformed methylation fractions. Accordingly, effect sizes reported in the Results represent the change in methylation fraction per alternate allele.

We tested each retained SV against all retained CpGs genome-wide, comprising 82,365,801 same-chromosome and 1,570,858,672 off-chromosome tests. We applied Benjamini–Hochberg false-discovery-rate correction across all SV–CpG pairs, using *q <* 0.01 for significance. We calibrated the global null distribution using 10,000 replicates of a Mammen score-multiplier wild bootstrap to obtain a maximum-statistic reference distribution. We retained all associations with *P ≤* 10*^−^*^4^ for downstream analyses.

#### Locality and genomic context

We compared distances between associated CpGs and their source SVs with a null distribution generated from 20,000 permutations. Permutations preserved the number of discoveries per SV and sampled CpGs from each SV’s tested-CpG distribution. For source SVs with at least one associated CpG within 5 Mb, we classified effect direction as predominantly negative if more than half of the raw *β* estimates within this distance were negative. We evaluated effect direction and magnitude separately for deletions and insertions using a two-sided binomial test of the proportion of predominantly negative SVs against 0.5 and the median *|β|* within 5 Mb, respectively. We annotated target CpGs with CenSat and classified them as satellite-resident or non-satellite, and by CenSat region into the *α*-satellite array containing the SV (HOR, diverged HOR and monomeric intervals merged within 500 kb), other centromeric sequence, the pericentromeric flanks (intervals labeled p_arm or q_arm) and sequence beyond the centromere (Supplementary Fig. S12e,f). We counted a CpG as dosage-responsive if its absolute effect in the observed data ranked among the top three effects from the real fit and 60 permuted dosage fits. SHEDs are bins of 2,200 CpGs within 5 Mb of an SV whose proportion of dosage-responsive CpGs, relative to the proportion beyond 20 Mb, exceeds the whole-track permutation envelope (Supplementary Fig. S10).

#### Conditional analysis of nearby SNP variation

To assess whether methylation-QTL associations were independent of nearby common SNP variation, we performed conditional analyses using two CHM13-referenced SNP panels: CHM13-aligned long-read variant calls from the 1000 Genomes ONT Sequencing Consortium data release [Gustafson et al., 2024] and the T2T-CHM13 1000 Genomes joint callset. In each panel, we fine-mapped SNPs within 1, 5, 10, and 20 Mb of the source SV using SuSiE [Wang et al., 2020], then tested the SV association conditionally in a cross-fitted analysis with fine-mapping and evaluation in disjoint sample folds. We considered associations robust to local SNP variation if SuSiE supported the SV and a direction-concordant conditional association remained for at least two distances in both panels. Separately, we conditioned associations on the SV’s elastic-net imputation score and its non-zero SNP predictors.

#### Robustness

We applied three sensitivity refits to the discovery set: adjustment for per-sample CpG read depth, leave-one-carrier-out analysis, and restriction to individuals with observed depth-qualified SV genotypes. The leave-one-carrier-out analysis required each association to retain its sign and at least half of its original *|β|* in every refit, while the genotype-complete analysis excluded all mean-imputed dosages. We report retention, sign agreement and significance for all three in Supplementary Fig. S14a–c. Genotype composition and Hardy–Weinberg fit for every discovery variant are given in Supplementary Table S12.

#### Measurement availability

To assess carrier-dependent CpG measurement in the assembly-based HPRC cohort, we matched each discovery CpG to twenty controls on source variant, order of distance, CenSat class, mean methylation and number of valid observations. We then regressed measurement presence on carrier dosage in both sets under a shared correction (Supplementary Fig. S11, Supplementary Note 9). We estimated effect sizes in the discovery cohort alone (Supplementary Note 9).

### 4.11 Centromere dip region calling and dip-position QTL mapping

#### CDR calling and annotation

We obtained haplotype-resolved 5mC from modkit bed-Methyl pileups for 384 HPRC Release 2 haplotypes. We restricted modification calls to 5mC alone (Supplementary Note 7). We then computed mean methylation in 5-kb bins in absolute reference coordinates, using `bedtools intersect -f 1` [Quinlan and Hall, 2010] to retain bins within the *active α*-satellite HOR array, defined per sample as the union of the active-HOR intervals of the global centromere annotation [Gao et al., 2026]. We called dips with CDR-Finder [Mastrorosa et al., 2024] at its recommended parameters (thresholds 0.34/0.30, merge distance 1 bp, edge distance 500 kb, baseline 0.4). We merged dips within 150 kb into annotated CDRs as in Gao et al. [2026]. We placed per-haplotype methylation in reference coordinates by walking the SVIM-asm CIGAR strings.

#### Definition of CDR phenotypes

Writing the active array as [`start`, `end`] with *covered* length *L* (the summed length of the merged annotated rows) and covered midpoint *m*, each dip’s relative position is (dip covered offset *− m*)*/*(*L/*2), running from *−*1 at the array start through 0 at its center to +1 at its end. We computed ten phenotypes per (sample, assembly haplotype, chromosome) over the active array only, then averaged them over an individual’s two haplotypes and rank-inverse-normal transformed them. They span four axes: position, scatter, abundance and level (Extended Data Table 2). Six of the ten are normalized by array length.

#### SV association testing

We tested all 11,129 cataloged loci across 23 chromosomes using SV genotypes derived directly from the assemblies. We derived genotypes per chromosome through deterministic haplogroup resolution on a bcftools merge [Danecek et al., 2021] cohort VCF, with cosine-neighbor-joining restricted to the 188 individuals overlapping the methylation cohort before tree construction. We grouped loci with identical genotypes in all 188 samples into 7,276 genotype groups, each with one test statistic, and report results per genotype group.

We tested each SV against each phenotype with 10 ancestry principal components and a base-caller indicator as covariates, under a Freedman–Lane [Freedman and Lane, 1983] permutation null restricted to the same chromosome. Sample size varies by chromosome with methylation overlap (e.g. *n* = 151 for chromosome 8). The genome-wide test used 10^5^ permutations; we re-ran each chromosome-by-phenotype pair with a locus at that floor at 10^7^ permutations, and a second round increased the permutations for every remaining pair with a locus within *∼*100-fold of the loosest threshold.

We excluded the two sensitivity phenotypes (Extended Data Table 2) from the Bonferroni correction, leaving eight main phenotypes and *α* = 0.05*/*(11,129 *×* 8) = 5.62 *×* 10*^−^*^7^.

#### Array-length and ancestry sensitivity analyses

*Array stability.* We tested each lead for associations of dosage with both array endpoints and array length, and excluded leads with dosage-dependent array length for every length-normalized phenotype. We also added the end-points as covariates. *Ancestry.* A within-superpopulation random-effects meta-analysis required minor allele frequency *≥* 0.05, at least twelve carriers, at least three informative superpopulations, consistent sign and leave-one-population-out stability. Chromosome X is reported separately and any chromosome-X candidate is sex-adjusted.

We also tested whether total SV burden was associated with CDR phenotypes. We rank-inverse-normal transformed burden per (individual, chromosome) and each phenotype within chromosome and fit *y_ic_ ∼* burden_cis_*_,ic_* +burden_trans_*_,i_*_(_*_−c_*_)_+quality+10 GRM PCs+(1 *|* individual) over 3,123 individual-by-chromosome rows, separating centromere-local from systemic or technical effects, with assembly-quality terms included. We confirmed significance using 1,000 individual-label shuffles and re-fitted every burden association with array length as a covariate; we report the adjusted estimates.

#### Conditional analysis and fine-mapping of the chromosome-8 locus

To ask whether the chromosome-8 lead is attributable to the deletion or to SNPs in linkage disequilibrium with it, we applied the same three-part evaluation used for the methylation-QTL leads (see Methylation-QTL association testing), against the identical association model (rank-inverse-normal dip position, intercept, ten ancestry principal components and a basecaller indicator). We used assembly-derived deletion genotypes. Each of the ten pericentromeric SNP panels used for conditioning (namely fixed intervals of 0.1, 0.2, 0.5, 1 and 2 Mb, the nearest 500, 1,000 and 2,000 SNPs to each centromere boundary, and the nearest 500 and 1,000 to the midpoint) served in turn as the competitor set. Within each panel we (i) fitted SuSiE jointly over the SV and up to 300 selected SNPs and read the SV posterior inclusion probability and credible-set assignment, (ii) conditioned on up to 20 SNPs pruned greedily from the strongest at *|r| <* 0.8, and (iii) ran a five-fold cross-fitted ridge residual test with a heteroskedasticity-robust score standard error. In (iii), covariate coefficients and SNP selection are estimated *within each training fold* and applied to held-out samples. We also report the maximum absolute correlation between the deletion and any single panel SNP, together with the number of individuals with discordant genotypes at the two.

For the chromosome-8 interval we calibrated the statistic directly. Taking the top lead as the conditioning variant, we computed the smallest conditional *P* over a declared candidate set, then repeated that computation on 2,000 phenotype permutations. The candidate set was the pre-specified leads, meaning the variants that independently reached study-wide significance in the main analysis before any conditioning; an open search over all distinct genotype vectors in chr8:44.2–44.5 Mb is a sensitivity analysis (Supplementary Note 7).

#### Sequence characterization of the chromosome-8 locus

We decomposed the chromosome-8 active *α*-satellite array of CHM13 v2.0 into monomers with HORmon v1.0.0 [Kunyavskaya et al., 2022], which identified nine monomer classes, grouped into seven major classes (M1 to M7), and a per-monomer partition of the array. As a sensitivity check we repeated the decomposition with StringDecomposer v1.1.2 [Dvorkina et al., 2020] against the same monomer set. We classified monomers shorter than 150 bp as shortened. We constructed repeat units by cutting the monomer class string at each occurrence of the unit’s boundary class. To compare the chromosome-8 count with other centromeres, we applied the same decomposition and the same threshold to all 26 active arrays of the reference, then tested the class distribution of the pooled shortened monomers against class abundance by permutation, with 200,000 draws. We tested class assignment of a shortened monomer against a prefix null of the 927 non-M5 monomers among 1,000 drawn from the same array (Supplementary Note 7).

For each of the 392 HPRC Release 2 haplotype assemblies we mapped two reference coordinates flanking each copy into assembly coordinates by the same SVIM-asm CIGAR walk used for coordinate lifting, then counted assembly bases between them. We assigned spans within 5 bp of a predicted length to that length and treated the remainder as unresolved.

### 4.12 Biobank-scale association test and fine-mapping in All of Us

#### Scoring

We applied elastic-net weights to short-read whole-genome genotypes in the All of Us Research Program [All of Us Research Program Genomics Investigators, 2024] (Controlled Tier, curated data release v8) with `plink2 –score` [Chang et al., 2015] (`variance-standardize`), giving a per-participant dosage *ĝ* = **X*β̂***. Of 11,129 predictors, we scored 10,799 that were scoreable in the All of Us variant panel. We retained only weight SNPs present in that panel above the MAF threshold and quantified per-SV predictor coverage before any association testing. Imputation accuracy is the cross-validated *R*^2^ of the model that generated the dosage, estimated in the training cohort by four-fold cross-validation (see SV genotype prediction with an elastic-net model).

#### Predictor sets

We assigned the 10,799 scored predictors to four pre-specified sets that defined the multiple-testing families: discovery predictors (3,416); negative controls (3,097); newly scoreable predictors with no discovery prior (4,207); and chromosome-X predictors (79). Negative-control predictors had posterior inclusion probabilities below 0.01 for all 464 traits in the summary-level fine-mapping (Supplementary Note 10) and otherwise follow the identical procedure.

#### Phenotypes and testing

We defined disease phenotypes using phecodeX. The main definition requires a minimum code count of two (MCC = 2), giving 2,530 phenotypes, of which 2,417 met the testing threshold of at least 50 cases and at least 50 controls. We analyzed the permissive single-code definition (MCC = 1) with the same pipeline as a case-definition sensitivity analysis (Supplementary Fig. S2). Reported case counts include only participants in the regression, after cohort intersection and sex restriction.

For phenotype *j* with case indicator **y***_j_∈ {*0, 1*}^n^*, the covariate matrix is

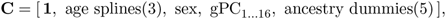

and the null

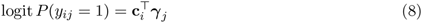

was fit once per phenotype by iteratively reweighted least squares, giving fitted probabilities *µ̂_ij_* and weights **W***_j_* = diag(*µ̂_ij_*(1 *− µ̂_ij_*)). Each SV dosage **g***_s_* was residualized on the covariate basis once per phenotype by the Frisch–Waugh–Lovell identity,

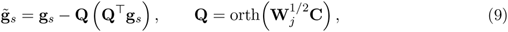

so that a single null fit serves every SV. The Rao score test for *H*_0_ : *β_sj_* = 0 in logit 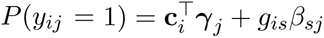 is then

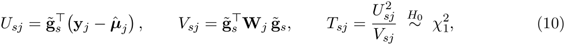

with *β̂_sj_* = *U_sj_/V_sj_*. This gave 26,101,183 valid SV–phecode tests. Genomic inflation was computed as 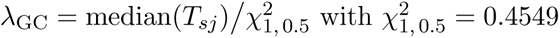.

We re-fit every SV–trait pair reaching *q <* 0.05 or *P <* 10*^−^*^4^ under the score test using exact Firth logistic regression [Firth, 1993, Heinze and Schemper, 2002], which maximizes the penalized log-likelihood

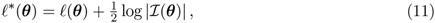

where *I* is the Fisher information. Significance came from the penalized likelihood-ratio test. When the exact fit did not converge, we substituted an approximate Firth recalibration; all associations reported as significant rest on the exact fit. We then applied Benjamini–Hochberg FDR [Benjamini and Hochberg, 1995] *within* each predictor set: for the *m_t_* tests of set *t* with ordered *P*_(1)_ *≤ · · · ≤ P*_(*m*_*t*_)_, we declared associations significant up to *k* = max*{ i* : *P*_(_*_i_*_)_ *≤ iα/m_t_ }* at *α* = 0.05. We corrected the negative-control set across its own 7,485,449 tests.

#### Score-test calibration

We validated the score test against the exact Firth fit within every carrier-count bin and every predictor set.

#### Per-ancestry analysis

We re-tested every SV–trait pair independently within each of the six ancestry groups, with exact per-ancestry Firth regression and within-group Benjamini–Hochberg correction. To test whether a signal restricted to one ancestry group reflects residual structure inside that group, we recomputed principal components within the AMR group, added them to the null model, and re-fit the association. We also re-fit SV–trait pairs after restricting to unrelated participants.

#### Quantitative traits

We tested the same predictors against a panel of quantitative laboratory and anthropometric traits, giving 755,930 SV–trait pairs. Each SV–trait pair received a cross-ancestry classification based on independent per-ancestry fits and an inverse-variance-weighted meta-analysis: consistent (including heterogeneous and power-limited subclasses) where per-ancestry estimates agree in direction and magnitude, possibly stratified where they do not, discordant where they oppose, and underpowered otherwise.

#### Chromosome X

The scoring pipeline applies a uniform 0*/*1*/*2 map, which for these data is the X-inactivation model. We re-scored chromosome-X SV–trait pairs under three ploidy models (the map in use, halved male dosage, and per-sex standardization) and computed the correlation between dosage and sex under each model.

#### Fine-mapping

We restricted fine-mapping to unrelated participants (*n* = 291,668, after removing 23,536 related or otherwise flagged participants, 7.5%), to keep relatedness from inflating the in-sample LD matrix; the EUR (*n* = 168,174) and AMR (*n* = 54,387) ancestry groups are nested within it.

We fit each candidate association under two panel constructions. The *single-SV* construction contains the target SV plus its most strongly correlated tag SNPs (median *∼*120); the *joint* construction additionally includes every same-chromosome SV, so an SV competes against other SVs as well as against SNPs. An SV included in a joint panel is termed a candidate if the disease screen nominated it and a competitor otherwise. We computed identifiability diagnostics on the full panel and after excluding the SV’s own elastic-net predictors. We fit each panel with three methods: an individual-level linear-probability model and summary-level fits on linear-probability and on logistic statistics. For each fit, we computed the LD matrix in sample as the correlation of the covariate-residualized dosage matrix. In total, we completed 476 fits.

#### Credible sets and significance labels

We extracted credible sets uniformly across methods with susie_get_cs at coverage 0.95 and purity threshold min_abs_corr = 0.5, with at most ten components. We labeled fits without a set meeting coverage and purity unresolved.

For each SV, we report credible-set inclusion and the highest posterior within that set separately. The main quantitative discriminator is the margin in *|z|* between the best SV and the best SNP in the same fit, together with the multiple *R*^2^ of the SV on its own panel. Negative-control predictors entered fine-mapping panels on the same terms as other SVs.

## Data availability

All datasets used in this study are listed in Supplementary Table S14 with their versions and accessions or URLs.

All of Us Controlled Tier data (curated data release v8) are available to authorized researchers through the Researcher Workbench (https://www.researchallofus.org/data-tools/workbench/); individual-level data remain within the Workbench. The UK Biobank genotyping-array marker set is available to approved researchers through the UK Biobank access application.

The haplogroup-resolved centromeric SVs, their imputed dosages and the molecular and disease associations reported for each can be browsed at CenSV (https://yikaidong-git.github.io/CenSV).

## Code availability

All custom code and pipelines used in this study are publicly available at GitHub:

- `cenSVhap`, the centromere haplogroup-resolution method introduced here, which builds a per-chromosome neighbor-joining tree over haplotype SV profiles and assigns each variant to the tree branch that best explains its carriers (https://github.com/YikaiDong-git/cenSVhap);
- `cenSVimp`, the centromeric-SV imputation framework, covering pericentromeric SNP panel construction, elastic-net training and cross-validation, and imputation into SNP-genotyped cohorts (https://github.com/YikaiDong-git/cenSVimp);
- `cenSV-analysis`, the workflows built on published tools for assembly alignment and structural-variant calling, expression, splicing and methylation QTL mapping, centromere dip-region calling, population-genetic summaries, the joint fine-mapping analysis, the All of Us Researcher Workbench drivers, the SVIM-asm simulation benchmark (Supplementary Note 13) and the figure-generation scripts (https://github.com/YikaiDong-git/cenSV-analysis);
- CenSV, the interactive browser for the variants and associations reported here (https://yikaidong-git.github.io/CenSV).

The published tools these workflows call, with the version of each, are listed in Supplementary Table S14.

## Data Availability

Haplotype-resolved assemblies for 196 HPRC Release 2 individuals, together with per-haplotype cenSat annotations, modkit 5mC bedMethyl pileups, Minigraph-Cactus chains, and the PAV call set, are available through the AWS Open Data program at s3://human-pangenomics/submissions/DC27718F-5F38-43B0-9A78-270F395F13E8--INT_ASM_PRODUCTION/ (https://humanpangenome.org/hprc-data-release-2/). HPRC sample and population metadata are available at https://github.com/human-pangenomics.
The T2T-CHM13 v2.0 reference, CenSat v2.1 satellite annotation, and Q100 HG002 v1.1 assembly are available at s3://human-pangenomics/T2T/ (https://github.com/marbl/CHM13). The global centromere annotation is available on Figshare at https://doi.org/10.6084/m9.figshare.29921990.
The 1000 Genomes 30-fold NYGC high-coverage phased genotypes, pedigree file, and GRCh38 alignments are available at http://ftp.1000genomes.ebi.ac.uk/vol1/ftp/data_collections/1000G_2504_high_coverage/ (https://www.internationalgenome.org/data-portal).
The 1000 Genomes ONT Sequencing Consortium 5mC bedMethyl pileups, CHM13-aligned read alignments and variant calls, and R9.4.1 assemblies are available through the AWS Open Data program at s3://1000g-ont/ (https://s3.amazonaws.com/1000g-ont/index.html). The T2T-CHM13 1000 Genomes joint callset used as the second conditional SNP panel is the SHAPEIT5-phased release at s3://human-pangenomics/T2T/CHM13/assemblies/variants/1000_Genomes_Project/chm13v2.0/Phased_SHAPEIT5_v1.1/.
MAGE v1.0 expression and splicing phenotypes are available on Zenodo at https://zenodo.org/records/10535719 and at https://github.com/mccoy-lab/MAGE.

## Acknowledgments

We thank the Human Pangenome Reference Consortium for the haplotype-resolved assemblies, the Telomere-to-Telomere Consortium for the CHM13 reference, and the MAGE, 1000 Genomes Project, 1000 Genomes ONT Sequencing Consortium, UK Biobank and FinnGen resources for making their data publicly available. The All of Us Research Program is supported by the National Institutes of Health, and we gratefully acknowledge All of Us participants for their contributions, without whom this research would not have been possible. Computational resources were provided by the Emory University Human Genetics Compute Cluster (HGCC). We thank Glennis Logsdon for insightful discussions regarding variant calling.

## Contributions

J.W. conceived the overall project. J.W. and Y.D. jointly developed the research questions and hypotheses. Y.D., J.W., and D.C. developed the methodology. Y.D. developed the software and performed the investigations and visualization. Y.D. and J.W. performed formal analyses. J.W. and D.C. provided resources and jointly supervised the study. Y.D. wrote the original draft. Y.D. and J.W. reviewed and edited the manuscript. All authors read and approved the final manuscript.

## Extended Data

**Extended Data Table 1:** Chromosome-8 dip-position cluster under median-of-dips and deepest-dip position. Three of the four SVs reach study-wide Bonferroni significance (*α* = 5.6 10*^−^*^7^ over 11,129 loci eight main phenotypes) at 10^7^ permutations under both definitions, the empirical floor *P* = 10*^−^*^7^. Allele frequency accompanies the best-supported haplogroup resolution of each SV (Methods). *n* = 151 individuals.

|  |  |  | Median of dips |  | Deepest dip |  |
|---|---|---|---|---|---|---|
| SV | Position (chr8) | AF | *t* | *P* | *t* | *P* |
| DEL, 113 bp | 44,300,221 | 0.55 | −6.17 | 1.0 × 10<sup>−7</sup> | −6.19 | 1.0 × 10<sup>−7</sup> |
| INS, 59 bp | 44,302,147 | 0.66 | −5.97 | 1.0 × 10<sup>−7</sup> | −6.06 | 1.0 × 10<sup>−7</sup> |
| DEL, 347 bp | 44,460,941 | 0.21 | +5.09 | 2.0 × 10<sup>−7</sup> | +5.37 | 1.0 × 10<sup>−7</sup> |
| INS, 1,868 bp | 44,247,324 | 0.12 | −4.99 | 9.0 × 10<sup>−7</sup> | −5.05 | 4.0 × 10<sup>−7</sup> |

**Extended Data Table 2:** The centromere-dip phenotype family. (Methods). Ten phenotypes describing four aspects of CDR organization (position, dispersion, abundance, and methylation level), computed per haplotype-chromosome over the active *α*-satellite array, averaged over both haplotypes, and rank-inverse-normal transformed. Relative position *x* is the dip midpoint from the array center scaled by half the array length: 1 at the array start, 0 its center, +1 its end. Extended Data Fig. 4 plots (*x* + 1)*/*2. Two *sensitivity variants* describe the same measurements under a different rule and serve as sensitivity analyses; the Bonferroni correction covers the eight main phenotypes. Six of the ten are normalized by array length, itself tested directly.

| Name in text | Code name | Axis | Definition | Length-norm. |
|---|---|---|---|---|
| Dip position | cdr_pos_offset | location | median relative position of the dips | yes |
| Deepest-dip position | cdr_pos_offset_deepest | location | relative position of the deepest dip | yes |
| Dip spread (IQR) | cdr_pos_iqr | scatter | interquartile range of relative position | yes |
| Dip spread (variance) | cdr_pos_var<sup>†</sup> | scatter | variance of relative position | yes |
| Dip count | n_cdrs | abundance | number of dips called in the array | no |
| Dip density | cdrs_per_mb | abundance | dips per Mb of array | yes |
| Hypomethylated fraction | cdr_fraction | abundance | summed dip width / array length | yes |
| Dip width | median_cdr_width | abundance | median dip width (bp) | no |
| Mean 5mC | mean_5mC | level | mean 5mC over 5-kb bins across the array | no |
| Coverage-weighted mean 5mC | mean_5mC_covwt<sup>†</sup> | level | the same, weighted by read coverage | no |

<sup>†</sup> Supplementary sensitivity variants, outside the Bonferroni correction; cdr\_pos\_var correlates with cdr\_pos\_iqr (*r* = 0.94) and mean\_5mC\_covwt correlates with mean\_5mC (*r* = 0.998). Both give zero hits at any threshold tested.

**Extended Data Fig. 1:**
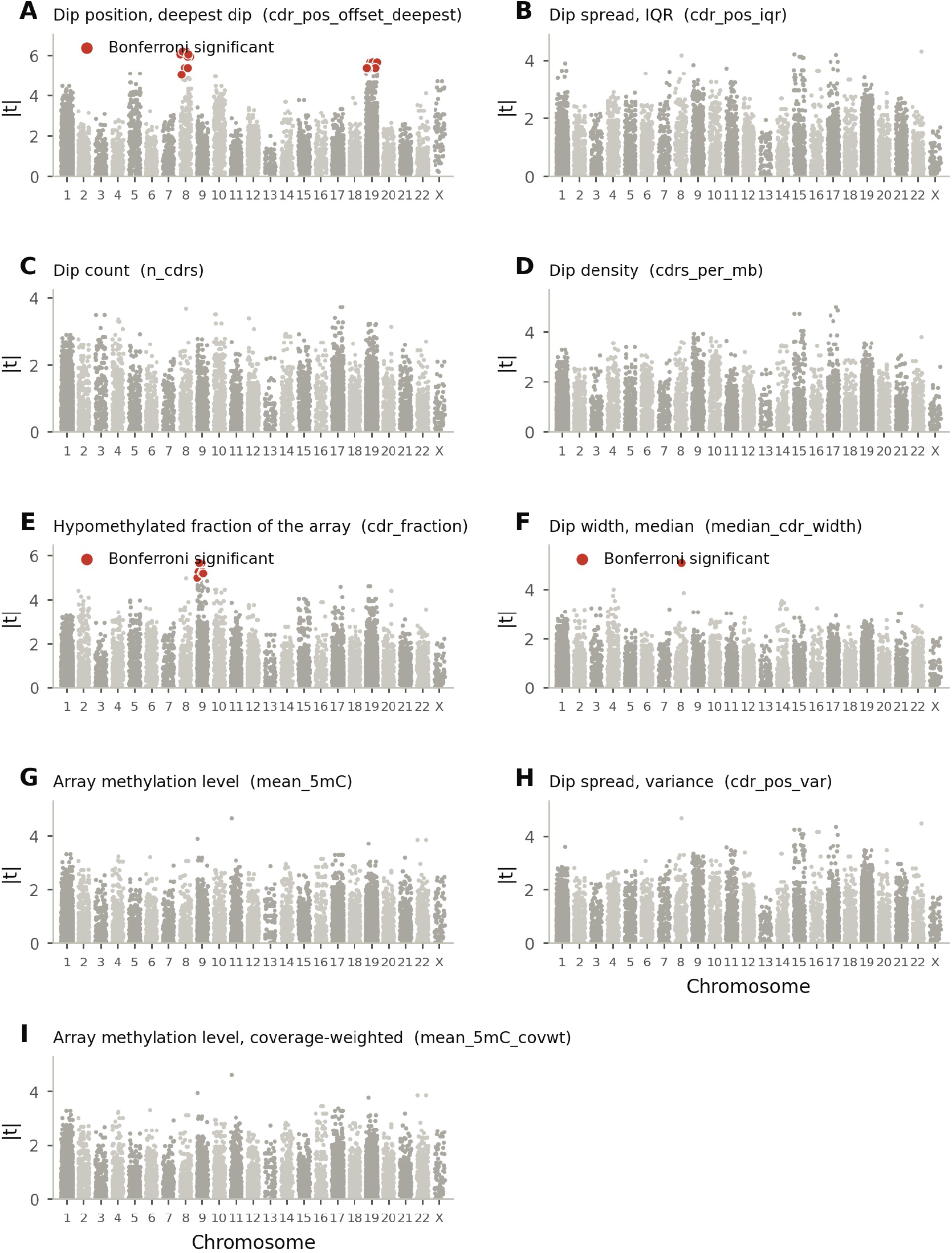
Centromeric SV association analysis for seven main-analysis dip phenotypes and two sensitivity variants. Association of every centromeric SV locus with each phenotype beyond Figure 4, over the 23 tested chromosomes (11,129 loci per phenotype). The y-axis shows *t* ; significance is determined from the permutation *P* value (Methods). Red points mark the tests reaching study-wide Bonferroni significance (*α* = 5.6 10*^−^*^7^) at 10^7^ permutations. Each panel has its own y-axis scale, and two uncolored tests in (a) lie above the lowest colored one. **a** Dip position, deepest dip: 25 tests are significant at *α*, recovering the chromosome-8 cluster. **b** Dip spread (IQR), **c** dip count, and **d** dip density give every *P* above *α*, the strongest being dip density at *P* = 1.0 10*^−^*^6^. **e** The hypomethylated fraction of the array: every point significant at *α* is the chromosome-9 cluster, excluded because array length is tested directly. **f** Dip width, median: one test is significant at *α* (*P* = 2 10*^−^*^7^) and is excluded at screening (Methods). **g** Array methylation level, the chemistry-insensitive phenotype. Its strongest *P* = 3.1 10*^−^*^6^ is above *α*. **h** Dip spread (variance) and **i** coverage-weighted array methylation level are the sensitivity variants of (b) and (g); in both, all *P > α*.

**Extended Data Fig. 2:**
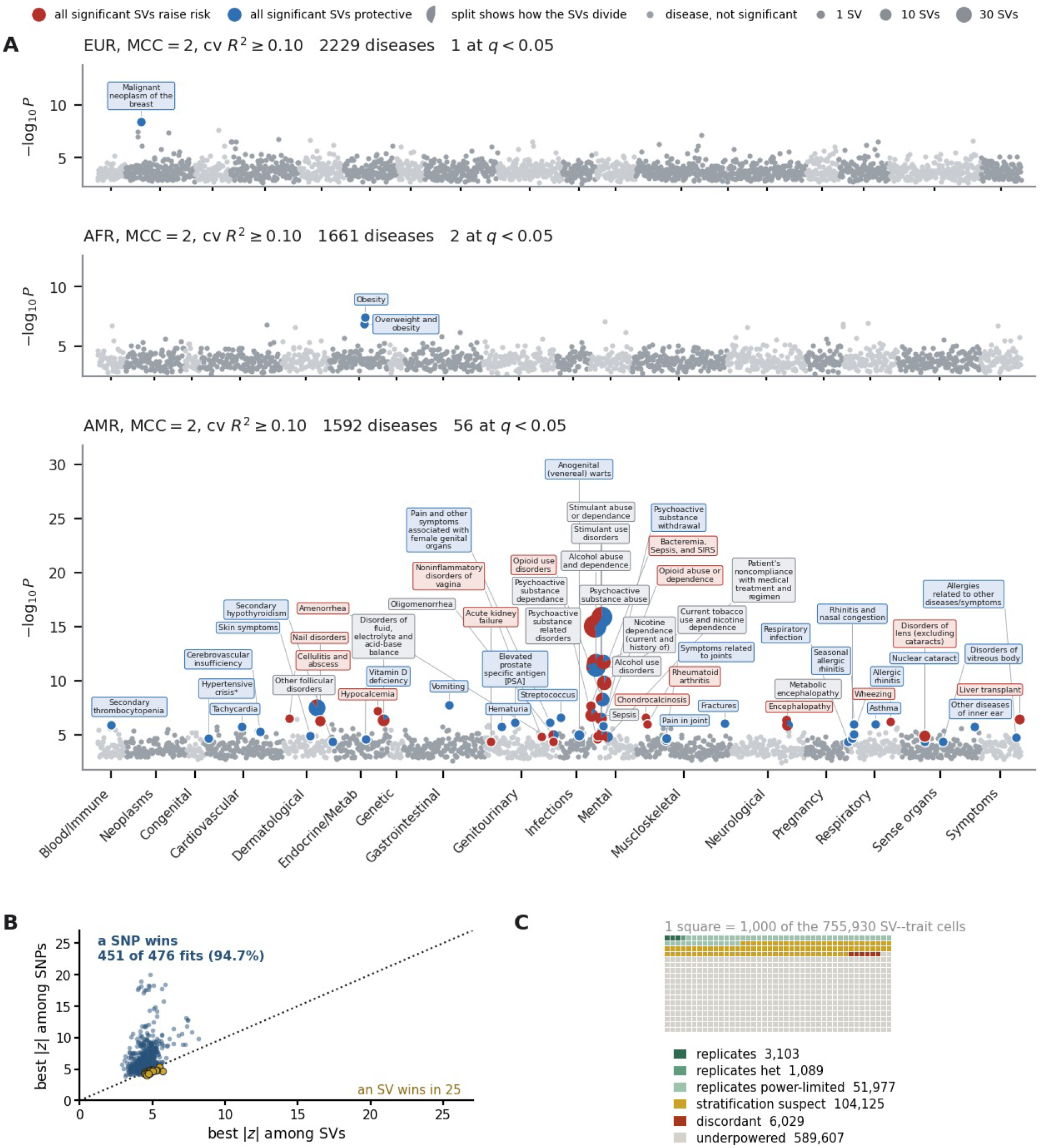
Ancestry-stratified All of Us association analysis and summary-level fine-mapping comparison. **a** Phenome-wide association analysis within each of six genetic-ancestry groups at *R*^2^ *≥* 0.10 (Methods), sizes EUR 177,230, AFR 64,726, AMR 60,984, EAS 7,144, SAS 3,944, and MID 1,176. Three ancestry groups with at least one phecode at *q <* 0.05 are shown. The admixed-American group has 56 significant phecodes, the European 1, and the African 2. **b** Best *|z|* among SNPs against best *|z|* among SVs, one point per fine-mapping fit. The dotted line marks equality. A SNP has the larger statistic in 451 of 476 fits (an SV in 25), by a median margin of 1.90. **c** Cross-ancestry classification across the 755,930 SV–trait pairs of the quantitative panel, one square per 1,000 pairs rounded to the nearest thousand. Each classification follows from independent per-ancestry fits; 56,169 SV–trait pairs are consistent, 589,607 underpowered, and the rest disagree across ancestry groups (Supplementary Note 8; Methods).

**Extended Data Fig. 3:**
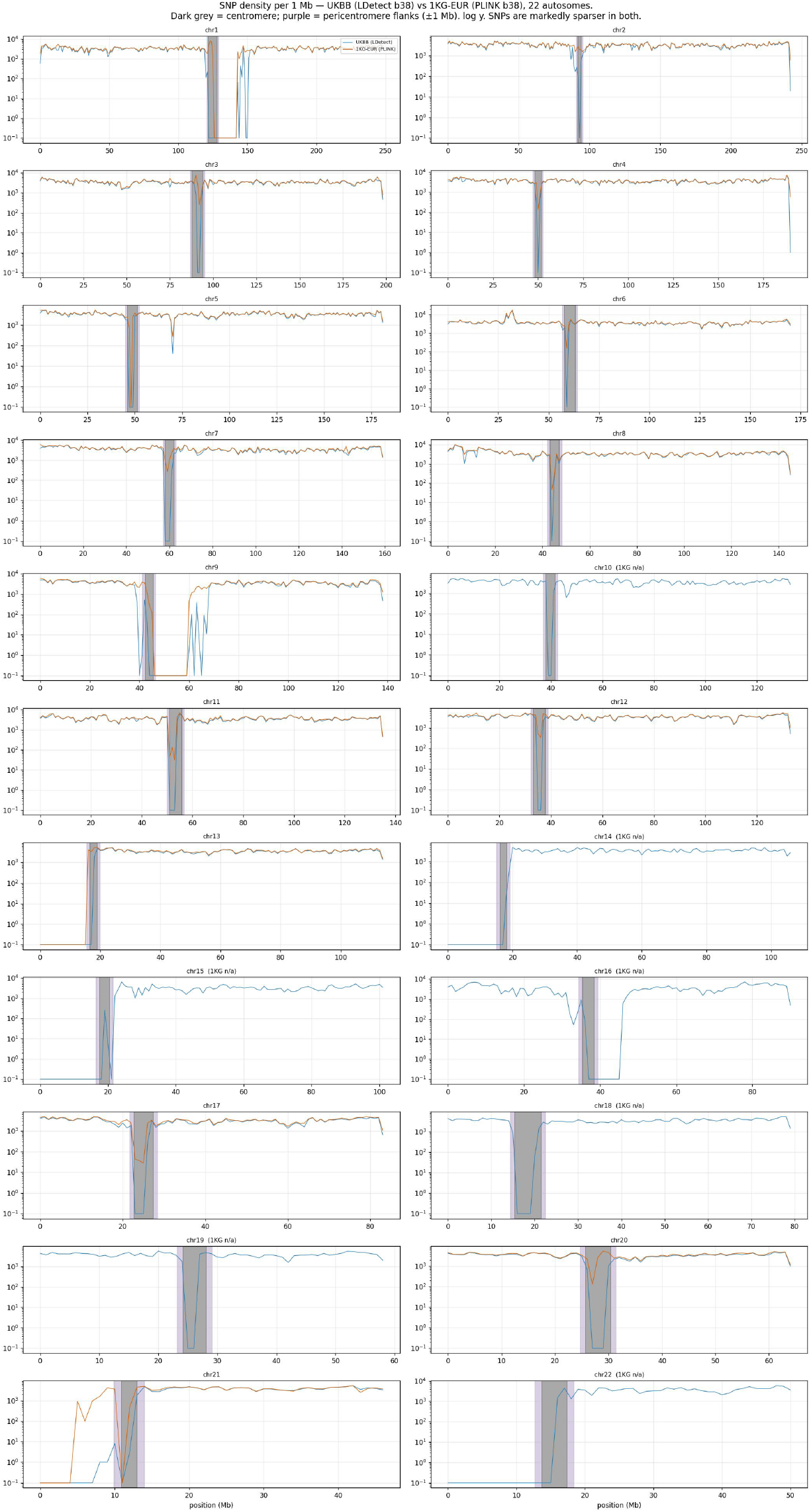
Pericentromeric regions have sparse SNP coverage in association-study panels. SNP density per Mb across the 22 autosomes for the UK Biobank genotyping panel and the 1000 Genomes European reference, logarithmic ordinate. Dark shading marks each centromere, light shading its 1 Mb flanks. Pericentromeric density averages 0.35 of the genome-wide value in the UK Biobank panel and 0.70 in 1000 Genomes. On chromosome 1 the UK Biobank panel contains 24.6 SNPs per Mb across the pericentromere compared with 2,971 per Mb elsewhere, with an absence of variants inside the centromere.

**Extended Data Fig. 4:**
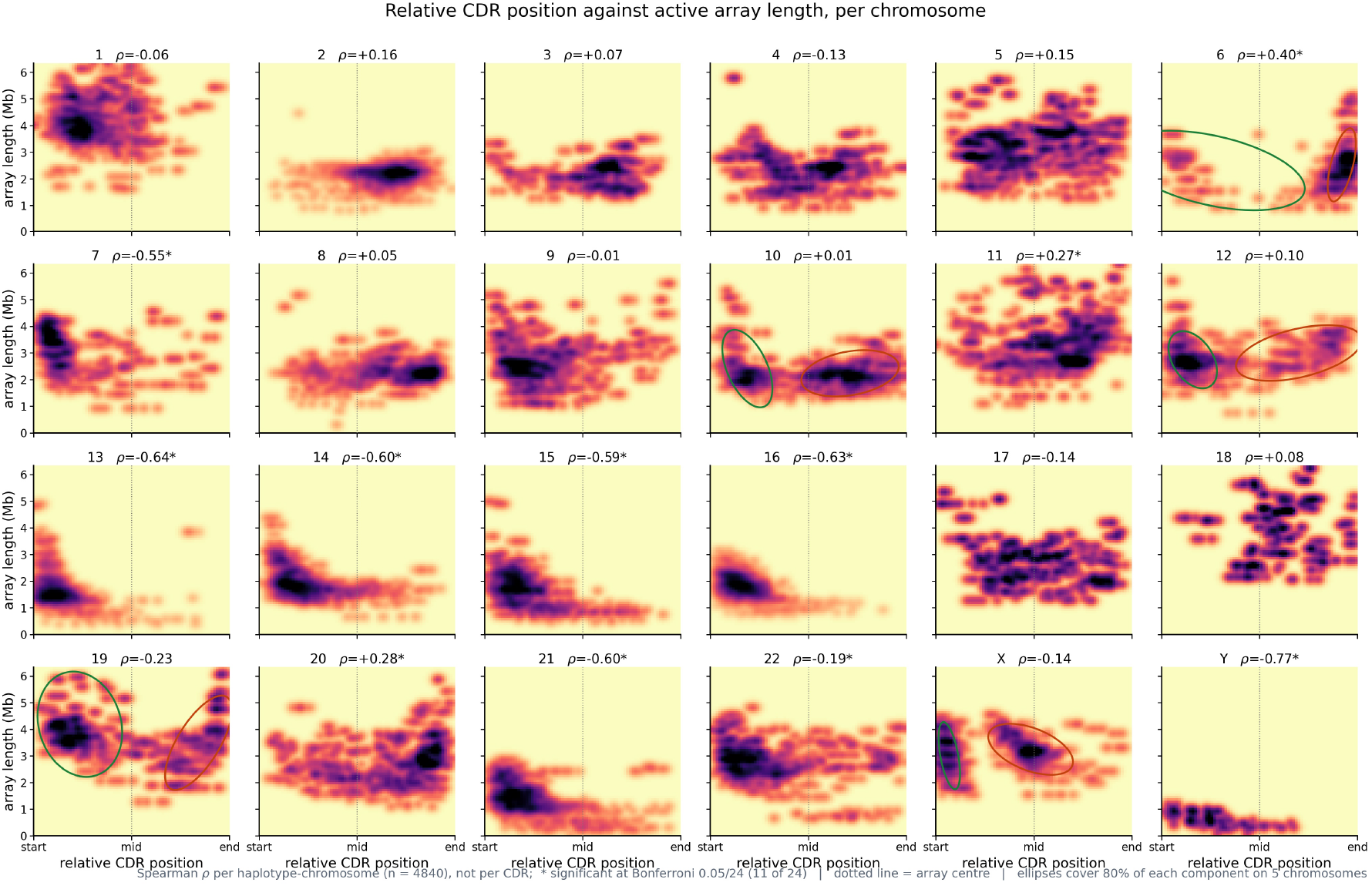
Relative dip position against active array length, per chromosome. Two-dimensional density of centromere dip regions over relative position within the active *α*-satellite HOR array (0 = array start, 0.5 = center, 1 = end) against array length, for the 24 chromosomes with an active array. We computed Spearman *ρ* per haplotype-chromosome with a called dip (*n* = 4,840; Methods); asterisks mark the 11 of 24 chromosomes reaching Bonferroni 0.05*/*24. Chromosome 8, which harbors the mapped locus, is one of the 13 unmarked chromosomes (*ρ* = +0.05, *P* = 0.43). The dotted line marks the array center, and the axis is clipped to 6.34 Mb, the 99.5th percentile of array length, leaving 105 of 21,597 dips (0.49%) undrawn. Array length correlates with relative dip position on the 11 of 24 asterisked chromosomes, most strongly on chromosomes Y (*ρ* = 0.77), 13 (*ρ* = 0.64), and 16 (*ρ* = 0.63). Ellipses mark the two preferred positions on chromosomes 6, 10, 12, 19 and X (Supplementary Table S10).

**Extended Data Fig. 5:**
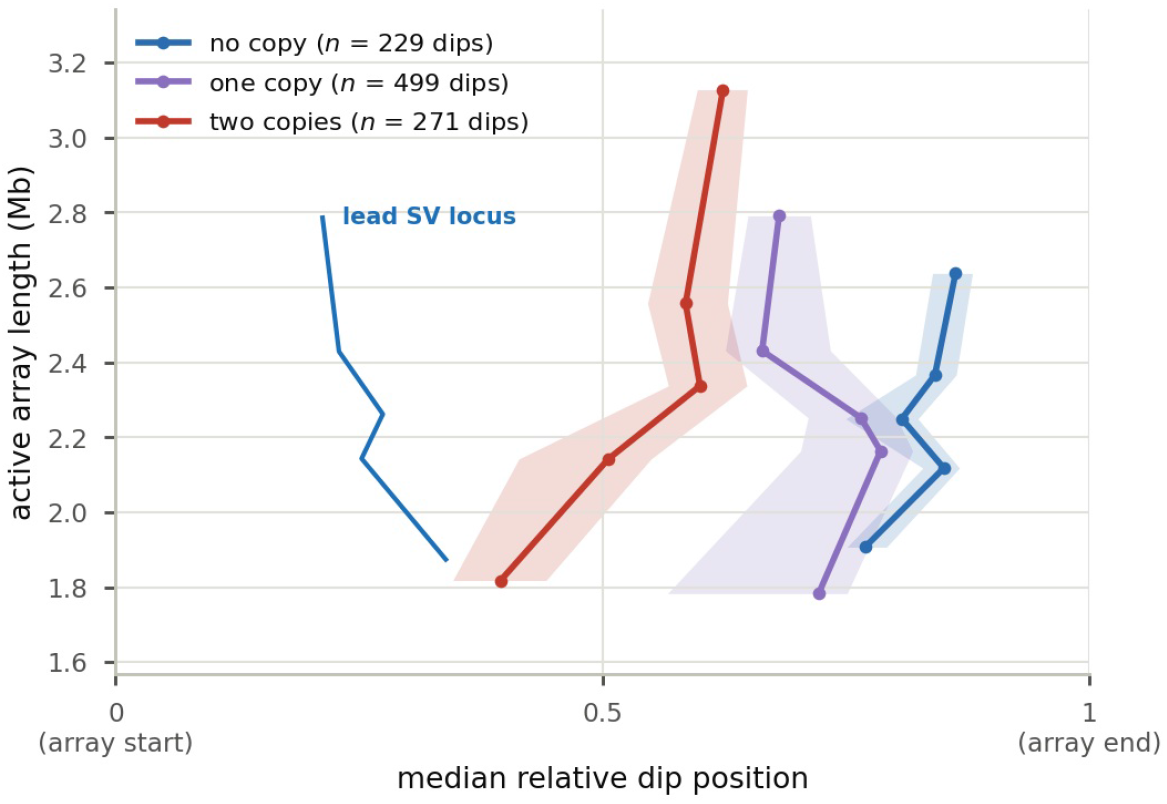
Dip position is associated with lead-deletion dosage. Median relative dip position against array-length quintile, one curve per dosage class, bootstrap 95% intervals on each median (800 resamples). Array length does not differ significantly across the three dosage classes (Kruskal– Wallis *P* = 0.23). A blue line marks the lead deletion’s locus, matched in relative position across classes (*P* = 0.63). The median dip lies 0.57, 0.46 and 0.30 from that locus across the three classes, with carriers’ dips nearer to it at every array length.

**Extended Data Fig. 6:**
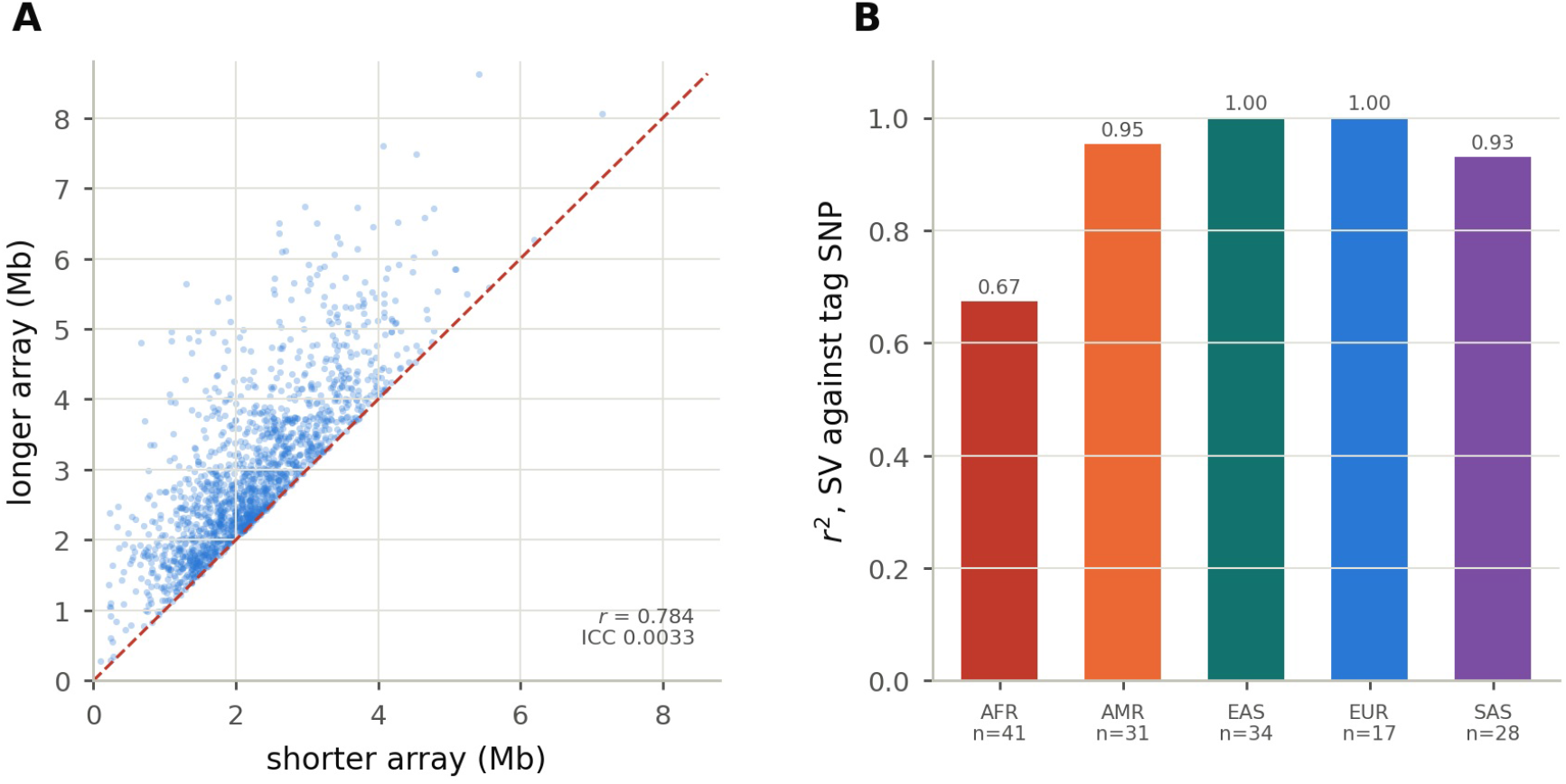
Array length varies between homologs, and SV–tag-SNP LD differs by ancestry. **a** Active array length against the paired array length of the same individual, *z*-scored within chromosome, 1,563 pairs (Methods). The intraclass correlation is 0.003 (*P* = 0.87), indicating little correlation in array length between homologs within individuals. **b** Linkage between the lead SV and its best tag SNP, by superpopulation. The pair is collinear in East Asian and European samples (*r*^2^ = 1.00) and separable in African samples (*r*^2^ = 0.67). The deletion shares the association with linked SNPs (Supplementary Table S11).

**Extended Data Fig. 7:**
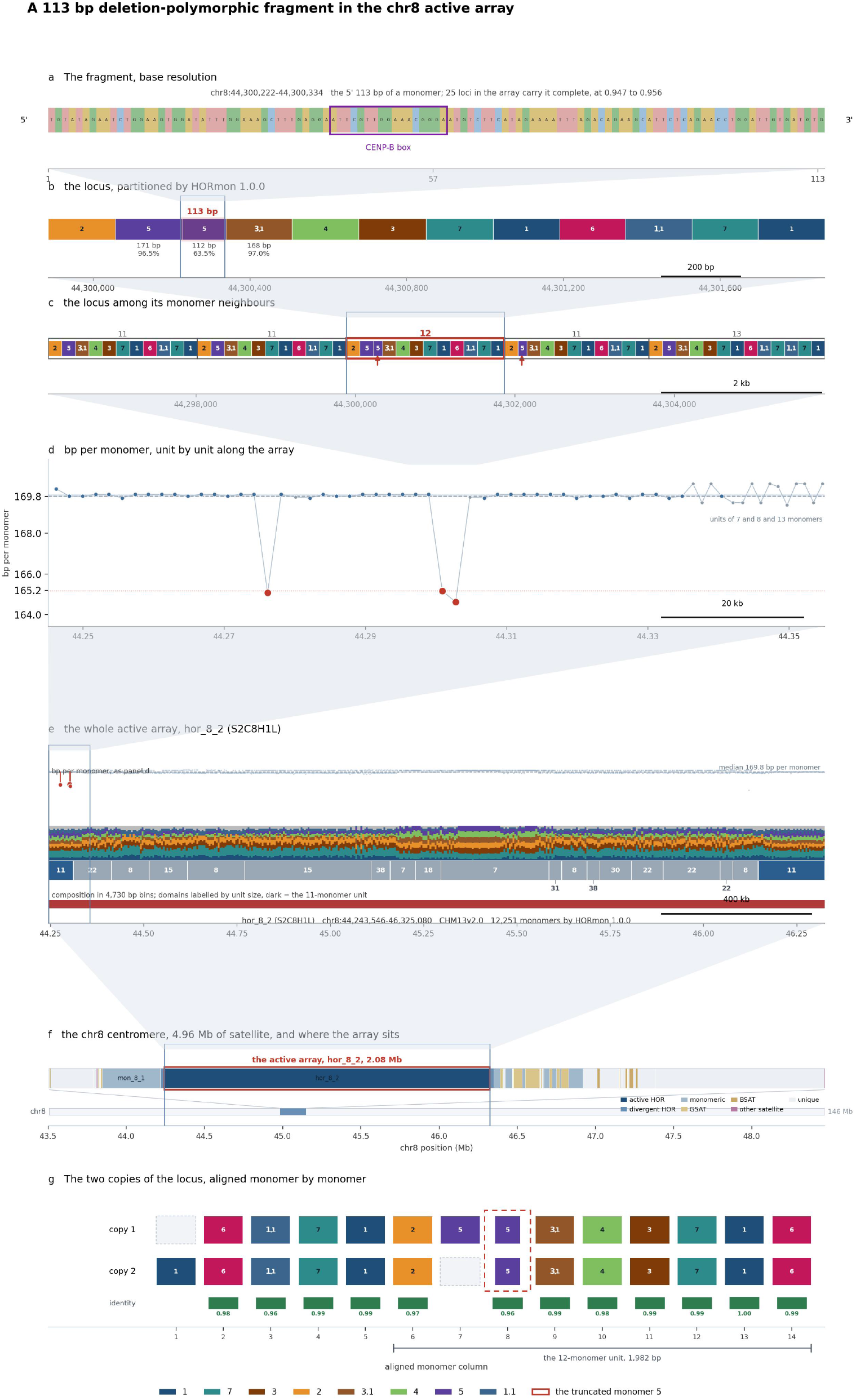
The chromosome-8 CDR-associated variant in the CHM13 reference. All panels show the chromosome-8 active array, 12,251 monomers, of which HORmon supplied the classes. We cut the class string at each occurrence of the boundary class to define repeat units, giving 1,514 units that cover the array (Methods). **a** The fragment at base resolution. **b** The locus, colored by HORmon monomer class. **c** The locus among its monomer neighbors. **d** Length in bp per monomer, unit by unit along the array. **e** The active array, hor_8_2 (S2C8H1L). **f** The chromosome-8 centromere. **g** Two copies of the locus.

## Supplementary Figures

**Figure S1:**
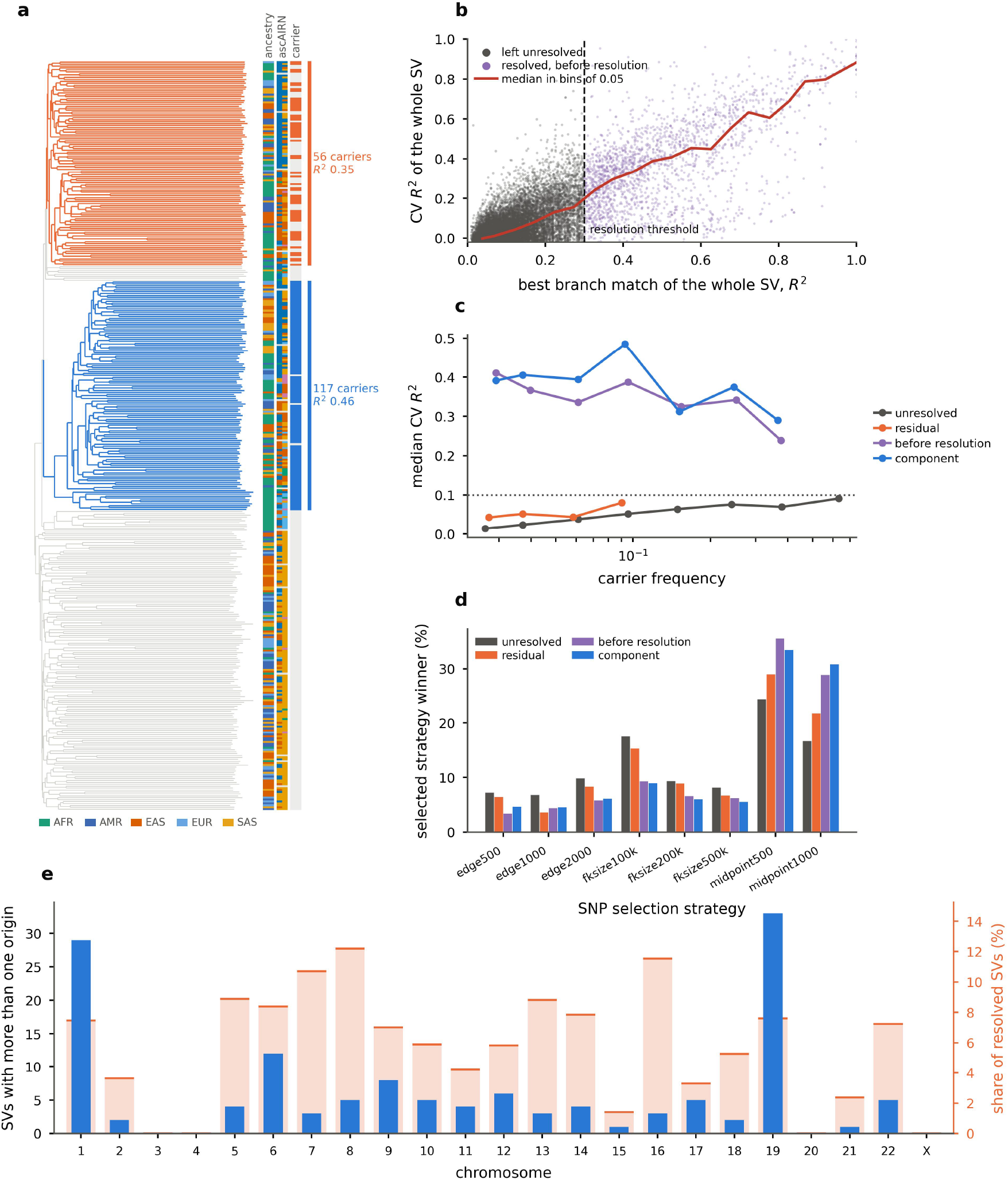
Haplogroup resolution of centromeric structural variants. **a** A 681 bp deletion at chr8:46,181,428 whose carriers form two clades of the chromosome-8 neighbor-joining tree over 392 haplotypes. The first clade spans 120 haplotypes of which 117 have the deletion (*R*^2^ 0.46); the second spans 107 of which 56 have the deletion (*R*^2^ 0.35). Each clade draws carriers from all five superpopulations, although the clustering did not use superpopulation labels. Ribbons give 1000 Genomes superpopulation, ascairn cluster and carrier state; ascairn assigns an unordered pair of clusters to each individual, so an individual’s two haplotypes have the same pair, and the nine individuals it did not type are left gray. Per-haplotype detail is in the CenSV browser. **b** Cross-validated *R*^2^ of each SV against the best branch match of its whole carrier set, for SVs left unresolved and for resolved SVs before resolution. Accuracy rises continuously through the *R*^2^ = 0.3 threshold. **c** Median CV *R*^2^ in bins of carrier frequency: monophyletic components maintain similar accuracy across the range, and unresolved SVs stay below 0.1. **d** Share of records selecting each of the eight SNP selection strategies. Midpoint strategies account for 64% of monophyletic components. **e** SVs with more than one origin per chromosome, as a count (bars, left axis) and as a share of that chromosome’s resolved SVs (blocks, right axis). 135 of 2,103 resolved SVs have more than one origin. All monophyletic SV trees can be viewed at CenSV (https://yikaidong-git.github.io/CenSV).

**Figure S2:**
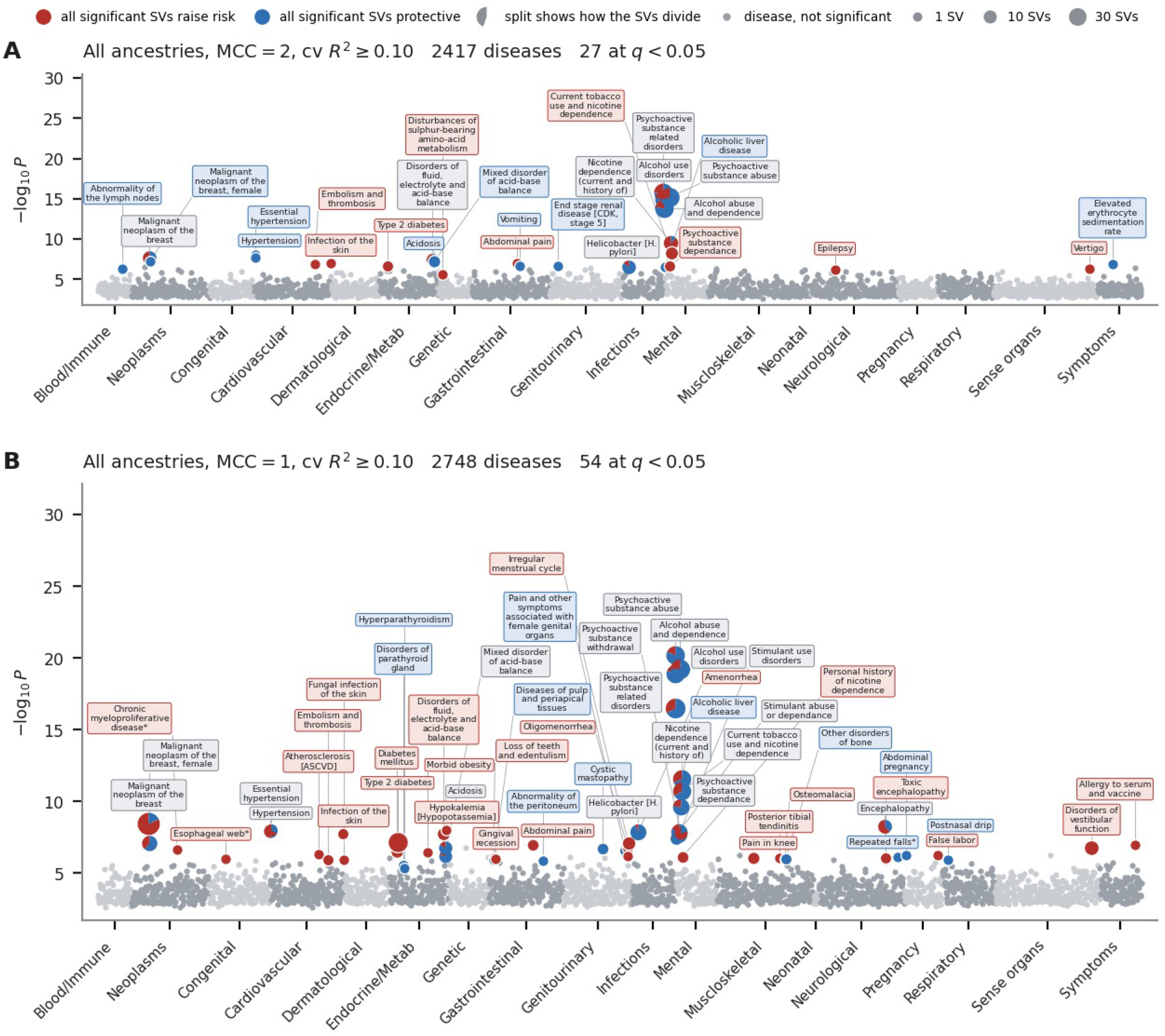
The phenome-wide association analysis under both case definitions. **a** Two occurrences of a phecodeX code required, the main definition and the one used in Figure 6a: 2,417 phecodes tested, 27 at *q <* 0.05. **b** One occurrence sufficient: 2,748 phecodes tested, 54 at *q <* 0.05. Relaxing the definition raises the sample size and makes 331 further phecodes testable. 20 of the 27 strict phecodes remain significant, 7 do not, and 34 are new. Each point is one phecode at its strongest predictor; the circle area indicates the number of distinct SVs associated with each significant phecode; red and blue sectors show the proportions of SVs that increase or decrease risk.

**Figure S3:**
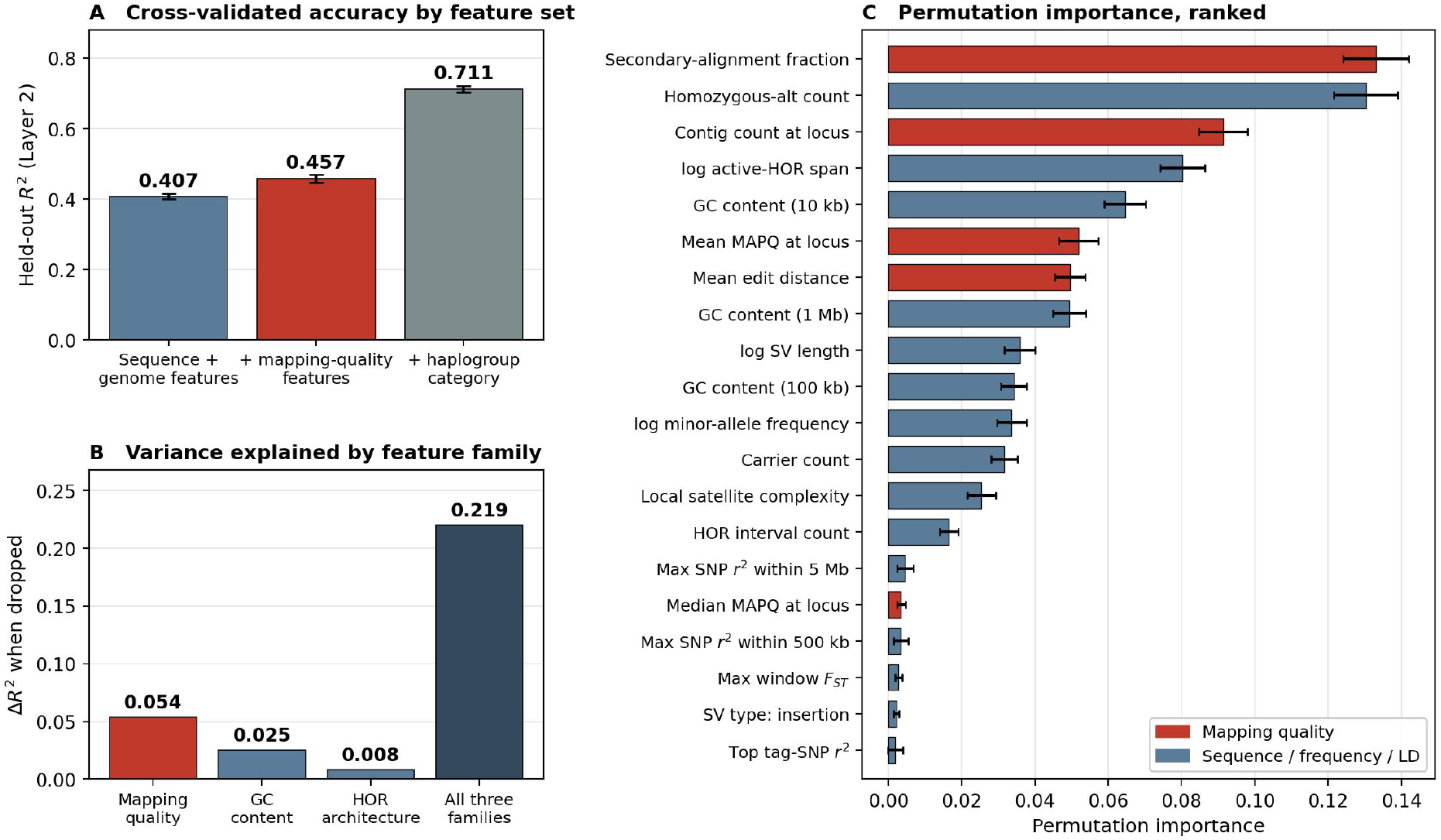
Locus mappability is the leading determinant of imputation accuracy. (Supplementary Note 3). **a** Held-out *R*^2^ of the Layer-2 model predicting per-SV cross-validated *R*^2^. Adding contig-level alignment features to the sequence- and genome-composition set raises prediction accuracy from 0.407 to 0.457. **b** Grouped drop-column analysis of three feature families. Removing the mapping-quality features reduces *R*^2^ by more than twice the reduction from removing GC-content features, and the three families together account for roughly half the modeled variance. **c** Permutation importance for the twenty leading features of the model including alignment features, of 43 in total. The fraction of secondary alignments at the locus ranks first, and four of the top seven features are mapping-quality measures (red); GC content over 10 kb ranks fifth. The leading 14 features account for 96% of the total, and every feature beyond them contributes less than 0.005. The three summaries of local SNP linkage disequilibrium rank 15th, 17th, and 20th.

**Figure S4:**
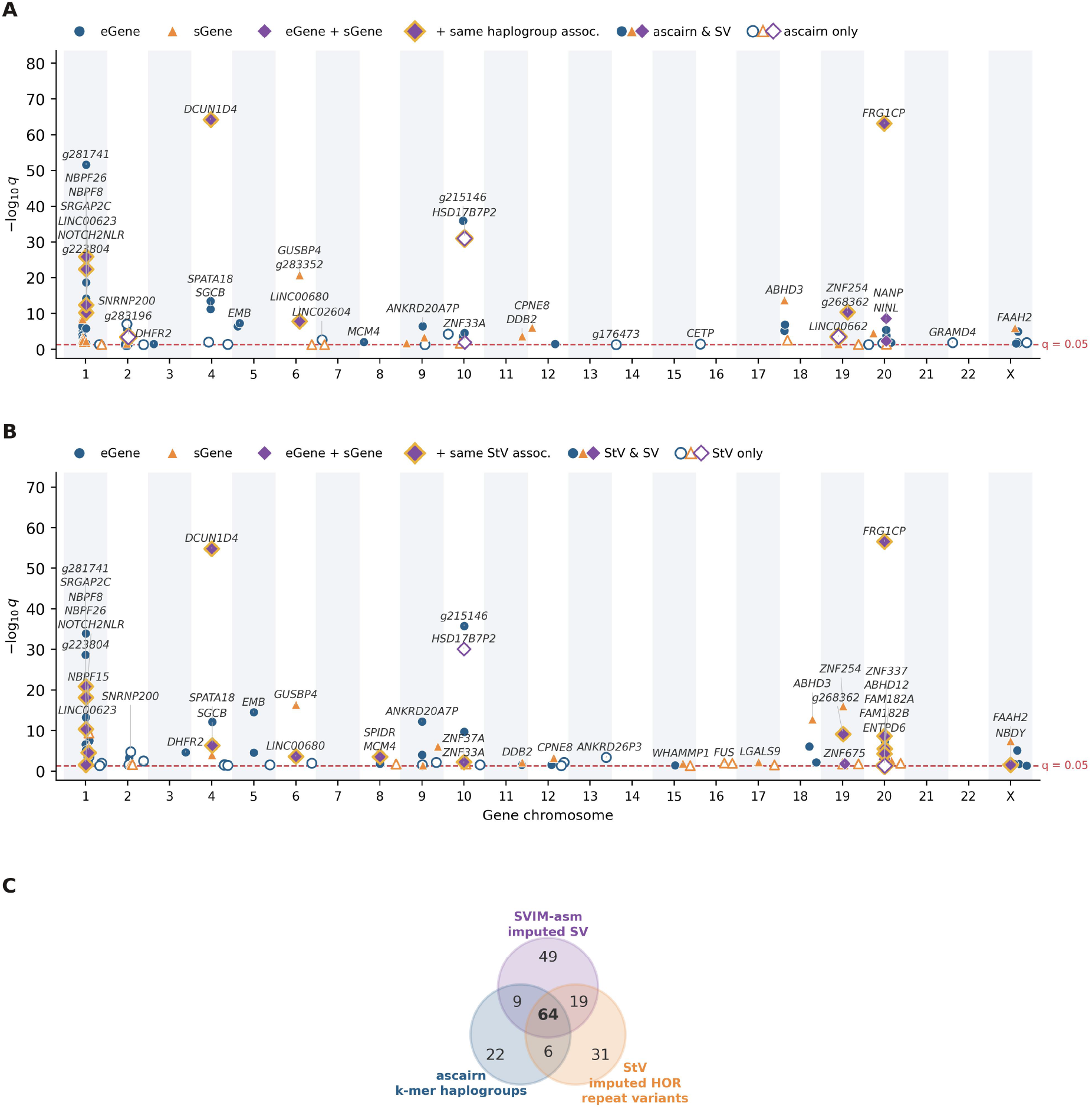
Cis-QTL genes from two alternative approaches to centromeric variation. Layout as in Figure 5a. **a** ascairn, an independent k-mer genotyping approach (*n* = 728): 101 cis genes, 73 among the 141 SV cis genes of the main analysis. **b** StV, an alternative variant definition on the same 630 individuals: 118 of 120 cis genes placed, 83 among the 141 SV cis genes of the main analysis. Filled markers indicate genes the main analysis also reports, open markers those specific to the method shown; gold outlines mark genes significant in expression and splicing on one shared variant. **c** Cis genes shared among the three methods: 64 reach significance under all three, over a union of 200.

**Figure S5:**
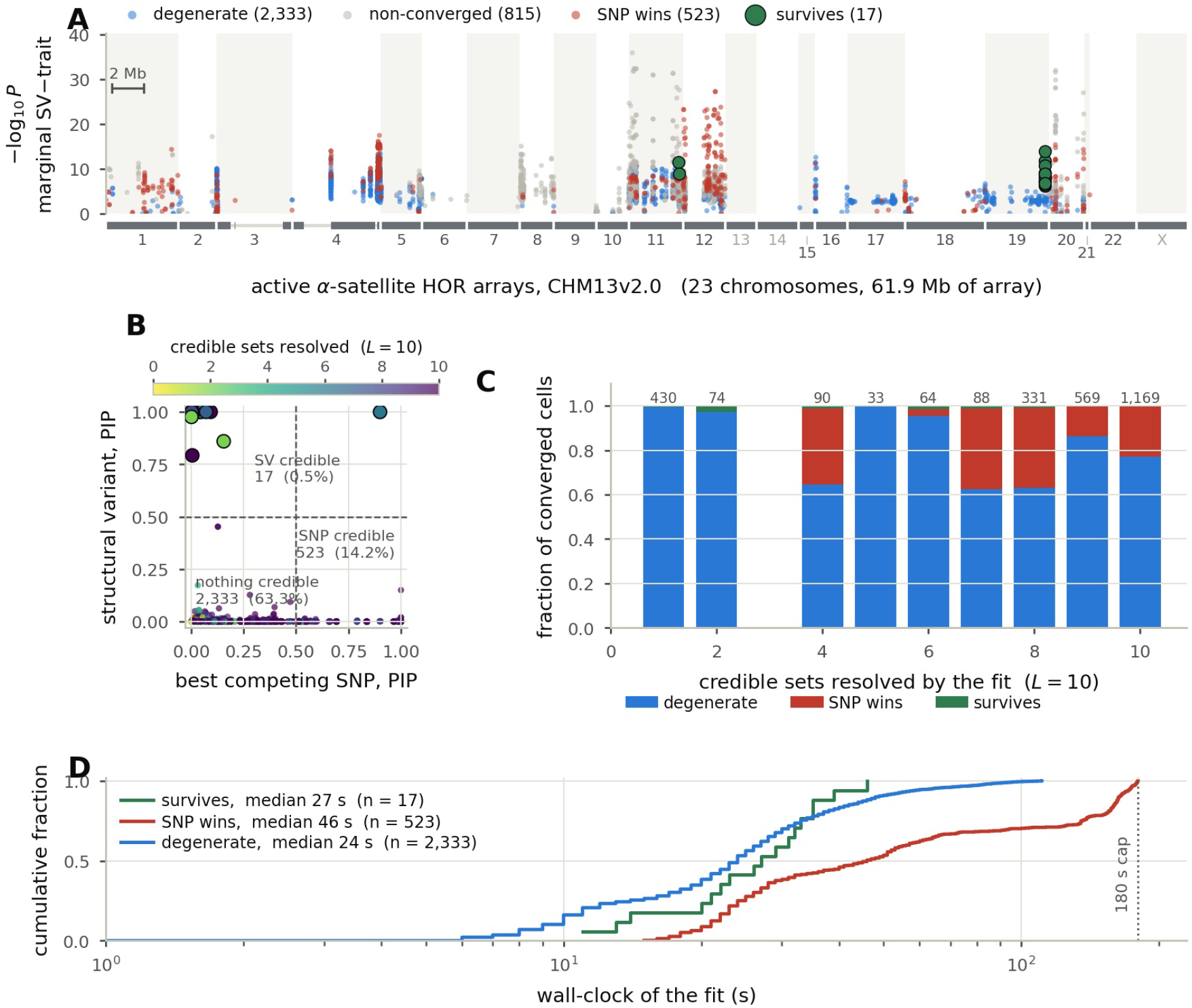
Imputed centromeric structural variants lead a credible set in 17 of 3,688 SV–trait pairs. **a** The 3,688 SV–trait pairs included at CV *R*^2^ 0.3 over 130 UK Biobank and FinnGen GWAS, drawn along the active *α*-satellite higher-order-repeat arrays (CenSat v2.1). Color marks the outcome of each SV–trait pair under a dense (12,000-SNP) re-fine-map. Three SVs lead a credible set, namely a chromosome-19 insertion at CV *R*^2^ = 0.999 and two chromosome-11 deletions. **b** SV posterior against the posterior of its best competing SNP for every SV–trait pair, colored by the number of credible sets resolved. Unresolved signal covers 63.3% of SV–trait pairs, a specific SNP 14.2%, and the SV 0.5% (17 pairs); non-convergence accounts for a further 22.1%. **c** Outcome of each SV–trait pair by the number of credible sets resolved. Fits resolving a single set leave 99.3% of SV–trait pairs unresolved, two sets 97.3%, and the *L* = 10 ceiling 77%. In 90.4% of unresolved SV–trait pairs the SV lies inside a broad credible set. **d** Wall-clock time by fit class. Unresolved fits are the quickest converged class, median 24 s.

**Figure S6:**
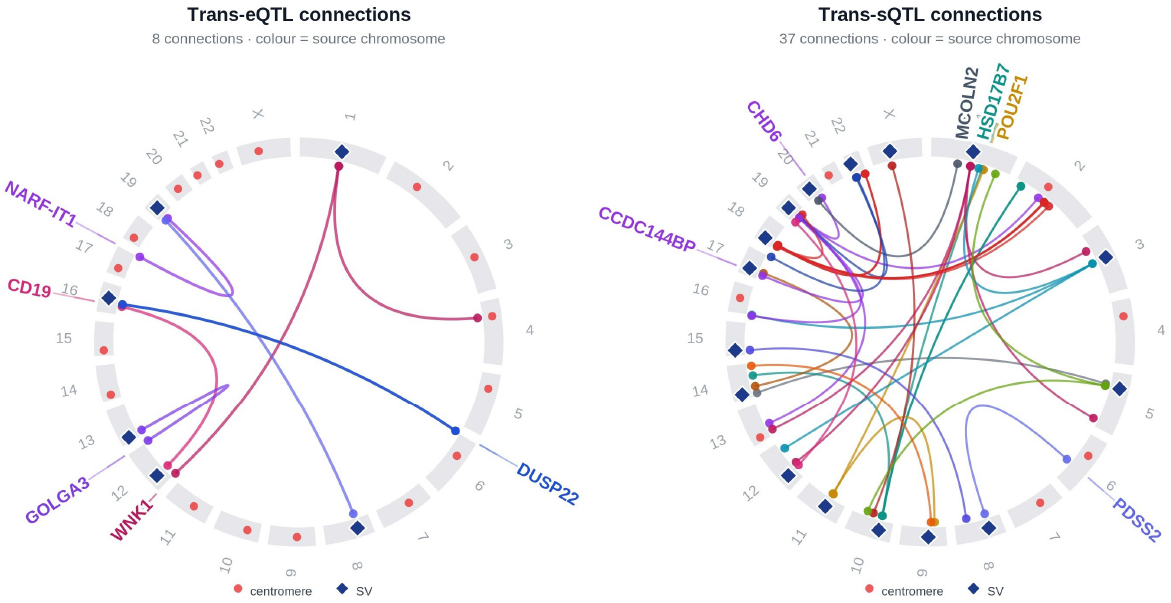
*Łraлs*-QTL connections of centromeric SVs across chromosomes. Diamonds mark each SV at its source-chromosome centromere; source-colored ribbons link them to target genes (circles) at *P <* 5 10*^−^*^8^. Of 46 target genes, 12 pass Benjamini–Hochberg at *q <* 0.1 over 1.26 10^9^ tests and 2 pass Bonferroni. Cross-mappability with a pericentromeric paralog removes *HSD17B7* [Saha and Battle, 2018a]; a non-reference paralog absent from GRCh38 removes *DUSP22*. Both Bonferroni-significant genes have paralogs. Empirical false-discovery rate 59.6% across the trans-QTL analysis (Supplementary Note 6).

**Figure S7:**
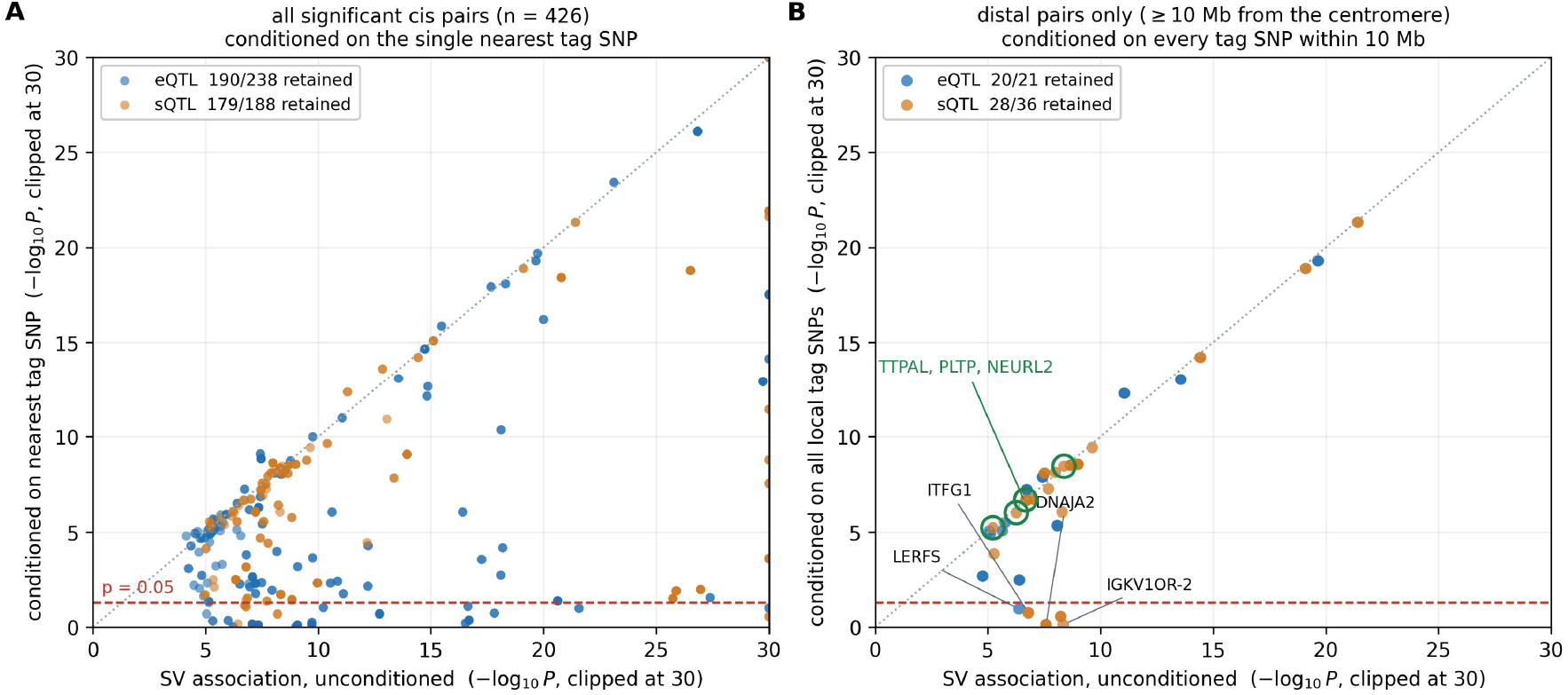
Cis associations conditioned on their local tag SNPs. **a** All 430 significant pairs under conditioning on the single nearest tag SNP; identifiable fits (*n* = 426): 190 of 238 eQTL pairs (79.8%) and 179 of 188 sQTL pairs (95.2%) retain significance. **b** Distal pairs under conditioning on all local tag SNPs (*n* = 57 non-degenerate fits): 20 of 21 eQTL (95.2%) and 28 of 36 sQTL (77.8%). We classified fits by the *R*^2^ of dosage on conditioners (informative *<* 0.90, marginal 0.90–0.99, degenerate 0.99), and excluded degenerate fits (Supplementary Note 6).

**Figure S8:**
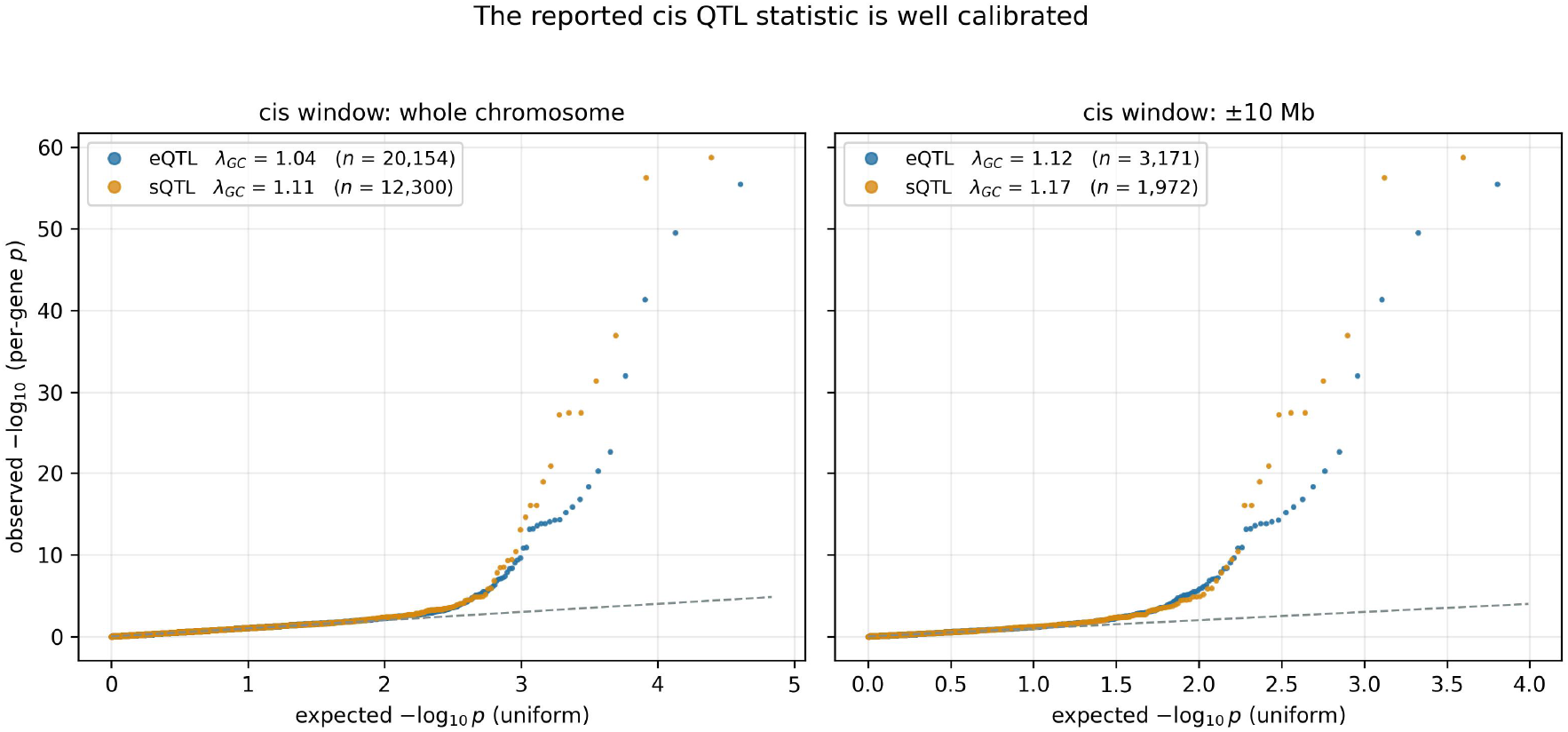
Calibration of cis-QTL permutation *P* values. Per-gene Beta-approximated permutation *P* value quantiles against the uniform expectation, pooled across the 23 chromosomes, for each cis region. Reported *q* values derive from these *P* values. Genomic-control *λ* is 1.04 (expression) and 1.11 (splicing) in the whole-chromosome region, and 1.12 and 1.17 in the 10 Mb region. Most quantiles follow the diagonal, with deviations concentrated among the smallest *P* values.

**Figure S9:**
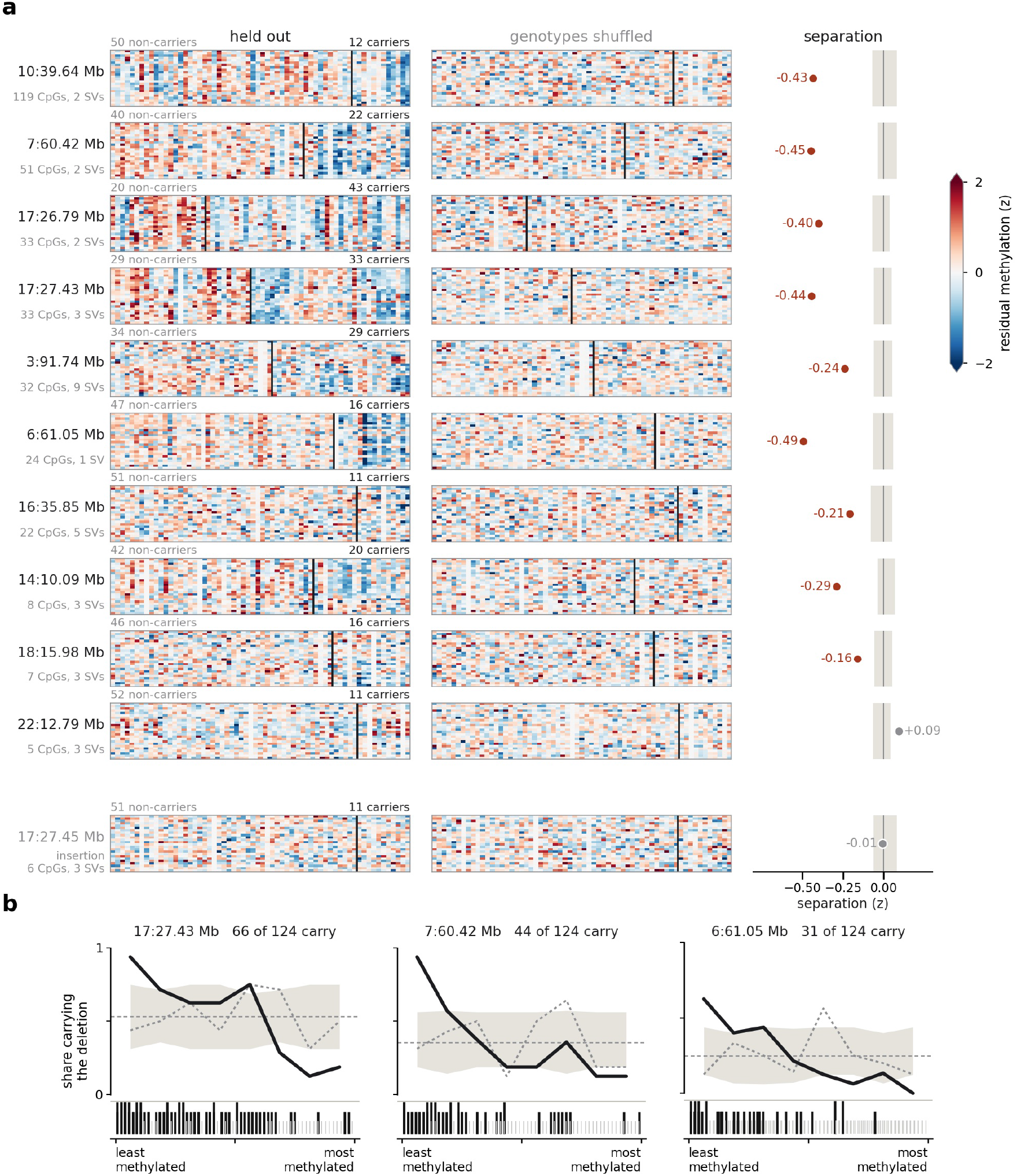
Out-of-sample prediction of carrier status from local methylation. **a** At each site, we selected 24 CpGs in one half of the 124 individuals and display the held-out half; columns are the displayed individuals ordered by deletion dosage, and the selected CpGs distinguish non-carriers from carriers. The middle column repeats the selection against a permuted dosage vector, on the same individuals in the same order. Gray spans the 5th to 95th percentile of 200 permutations. Nine of the ten deletion sites fall below their own band; chr22:12,790,199 and the insertion control lie within or above theirs. **b** Individuals ranked by methylation at CpGs chosen in the half of the cohort they are not part of, at the three sites with whole-region measurement. The line is the proportion of individuals with the deletion along that ranking, the band the same share with carriers assigned at random, and the dotted line the same people ranked on random CpGs from the region.

**Figure S10:**
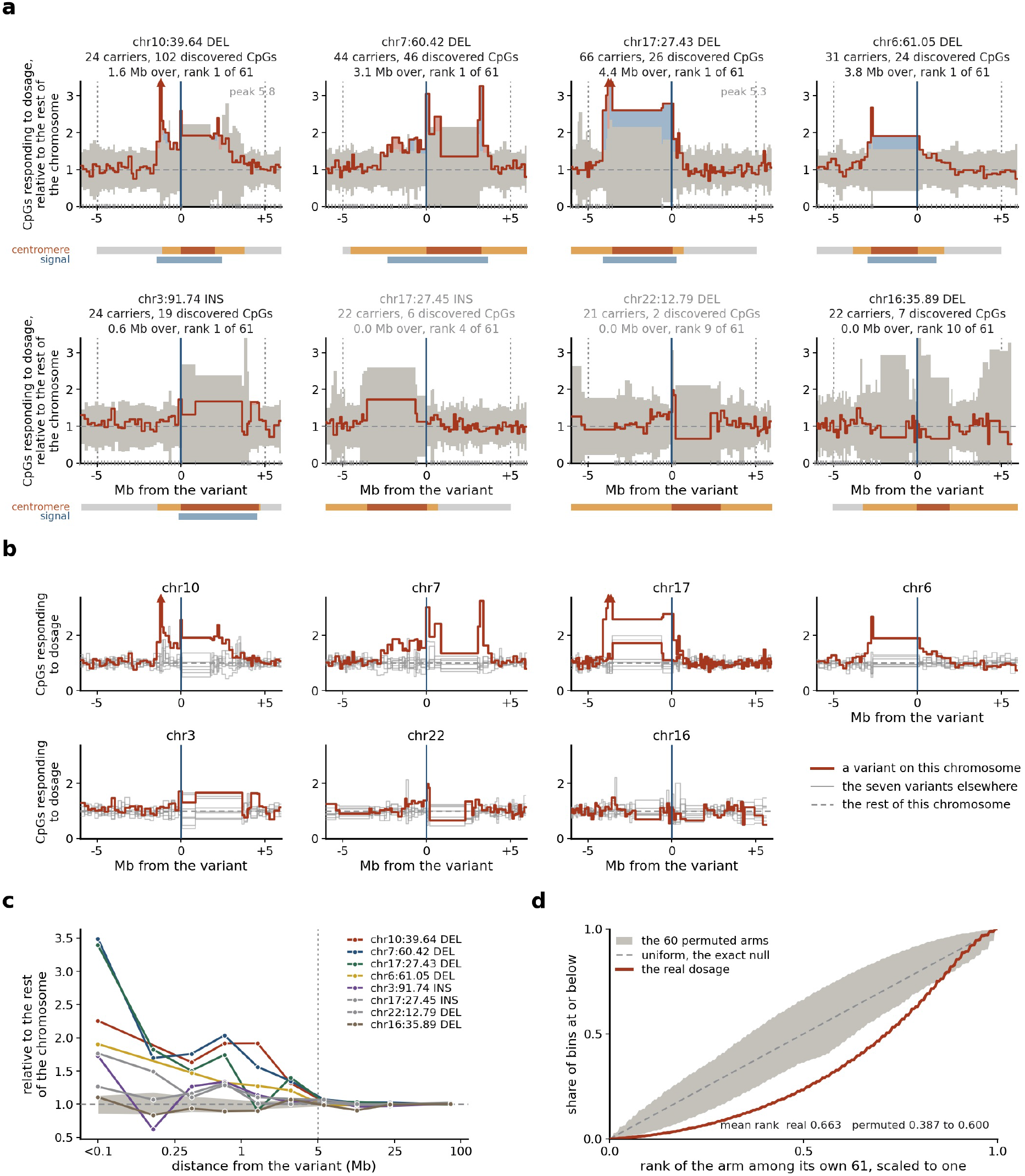
Methylation associations around each variant, across all measured CpGs. **a** Share of CpGs whose methylation is associated with deletion dosage, in bins of 2,200 CpGs, compared with the same share beyond 20 Mb on the same chromosome, so a value of 1 denotes the association frequency in the rest of that chromosome in the same individuals. Gray marks the envelope of 60 permuted dosage vectors as whole tracks; shading marks the parts above it, blue where deletion carriers have lower methylation. Each title gives the SHED width, above the envelope within 5 Mb, and its rank among the observed track and 60 permuted tracks. Bars beneath each panel show the CHM13 CenSat centromere, colored by its active HOR array (red), other centromeric sequence (orange) and the pericentromeric flanks (light gray), and the extent of the signal. **b** The same construction at every chromosome against all eight dosage vectors: red for chromosomes on which the variant is located, and gray for the seven other dosage vectors. **c** The same quantity by distance, every measured CpG in one band. **d** The same quantity in 65,823 bins more than 25 Mb from any source variant. The observed dosage vector ranks higher than any permuted one.

**Figure S11:**
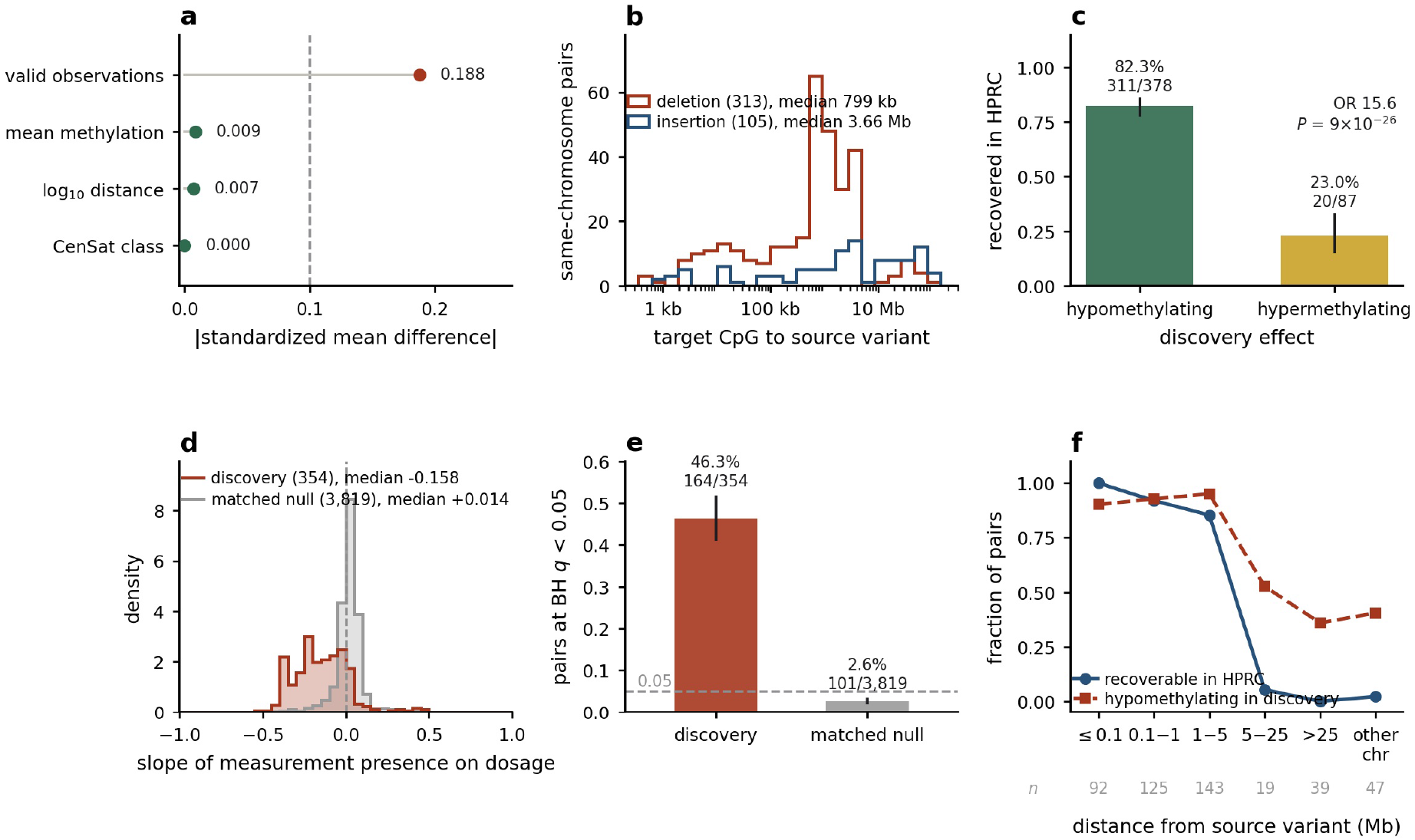
Measurement availability at candidate CpGs depends on carrier status; CpG measurement in HPRC. (Human Pangenome Reference Consortium Release 2; 156 individuals, 311 haplotype assemblies, 150 with assembly-derived genotypes, disjoint from the 124 discovery individuals). **a** Balance of the matched null, twenty control CpGs per discovery CpG matched on source SV, order of distance, CenSat class, mean methylation, and valid-observation count, excluding a region around the source event (Methods); 6,280 matched pairs over 4,715 control CpGs. Valid-observation count remains imbalanced, 0.188 against the 0.1 line. **b** Distance from each same-chromosome target CpG to its source variant, medians marked. Targets fall outside the deleted interval of their source SV, 418 of 418; missing calls therefore cannot be explained by deletion of the target CpG. A further 47 lie on another chromosome. Deletions lie 4.6-fold closer to their targets than insertions. **c** CpG measurement in HPRC by the sign of the discovery effect, Wilson 95% intervals. Fisher odds ratio 15.55, *P* = 8.9 *×* 10*^−^*^26^. **d** Per-pair slope of measurement presence on carrier dosage, negative in 302 of the 354 testable discovery pairs (85.3%). **e** The same comparison under one shared Benjamini–Hochberg family. The matched-null rate lies below nominal. **f** Recoverability and discovery direction against distance. The pericentromeric region assayed for methylation spans 6.8 Mb per contig, and this window excludes the more distal pairs at which the discovery effect was hypermethylating.

**Figure S12:**
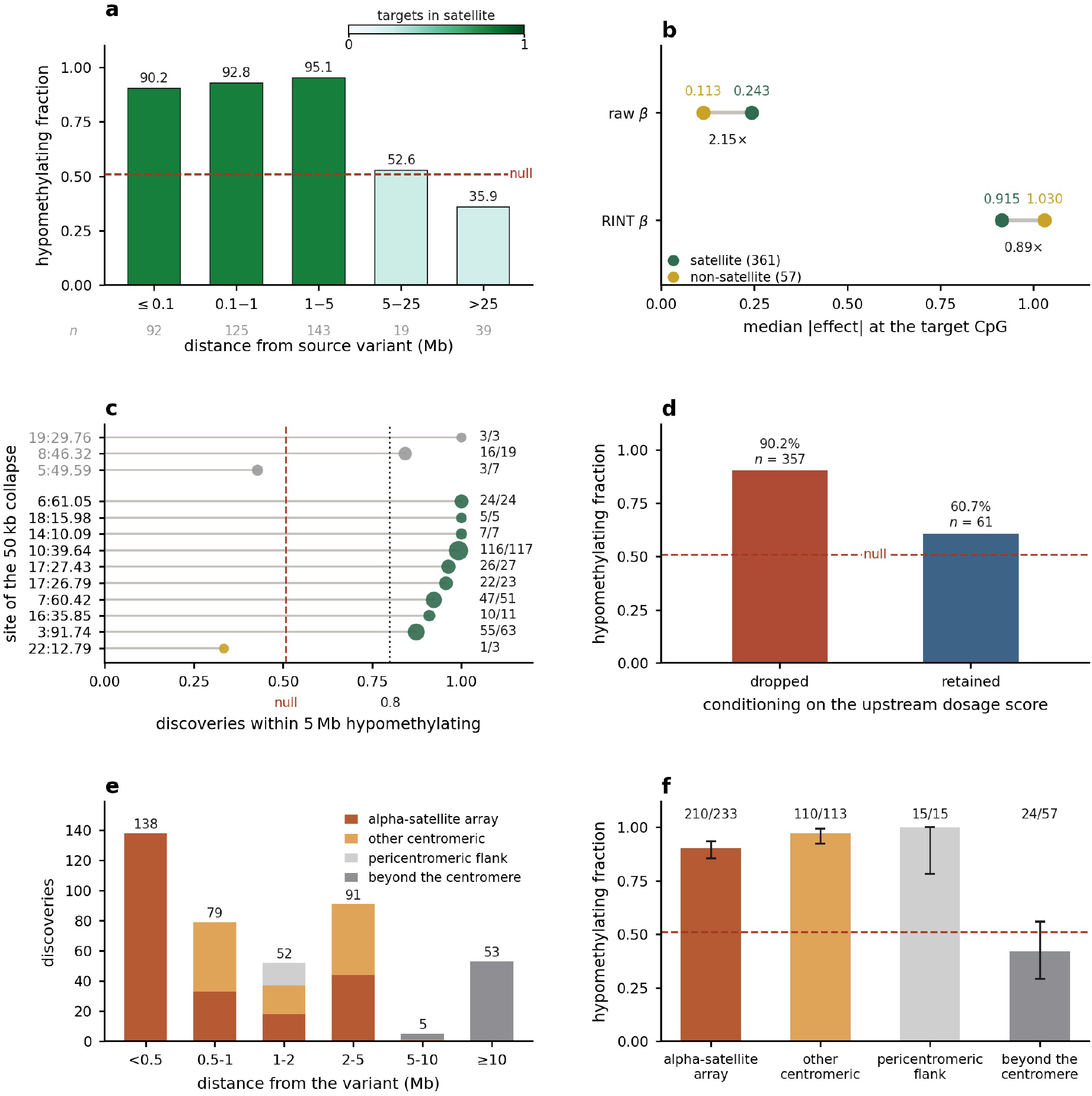
Direction and effect scale of the 418 same-chromosome discoveries. (361 in satellite sequence, 57 outside it). **a** Hypomethylating fraction by distance. Fill marks the satellite content of each bin, and a dashed line the matched null at 50.9%. Satellite-resident targets lie within 6.49 Mb of their source variant, non-satellite targets beyond 8.66 Mb. The observed fraction is 85.9% overall; within satellite 92.8%, outside it 42.1%. **b** Median absolute effect for satellite against non-satellite targets, on the untransformed reporting scale and the rank-inverse-normal testing scale. The transformed scale reverses the raw ordering (Methods). **c** Direction by site, with discoveries merged within 50 kb, among the 360 discoveries within 5 Mb, the unit of analysis in this panel and in Supplementary Fig. S9a: a chromosome can contain two sites and one site can contain nine variants. Marker area scales with the number of pairs; reference lines mark the matched null and 0.8. Nine of the ten deletion sites are at least 80% hypomethylating, chr22:12.79 Mb at 1 of 3. Insertion sites are gray, chr5:49.59 Mb at 3 of 7. **d** Direction split by retention of effect after conditioning on the upstream elastic-net dosage score (Supplementary Fig. S14d). The consistent hypomethylating direction is concentrated among pairs that are no longer significant after conditioning. **e** Discoveries by distance from the variant, colored by CHM13 CenSat region, the *α*-satellite array holding the variant, other centromeric sequence, the pericentromeric flanks and sequence beyond the centromere. **f** Hypomethylating fraction by region, with 95% intervals and the matched null.

**Figure S13:**
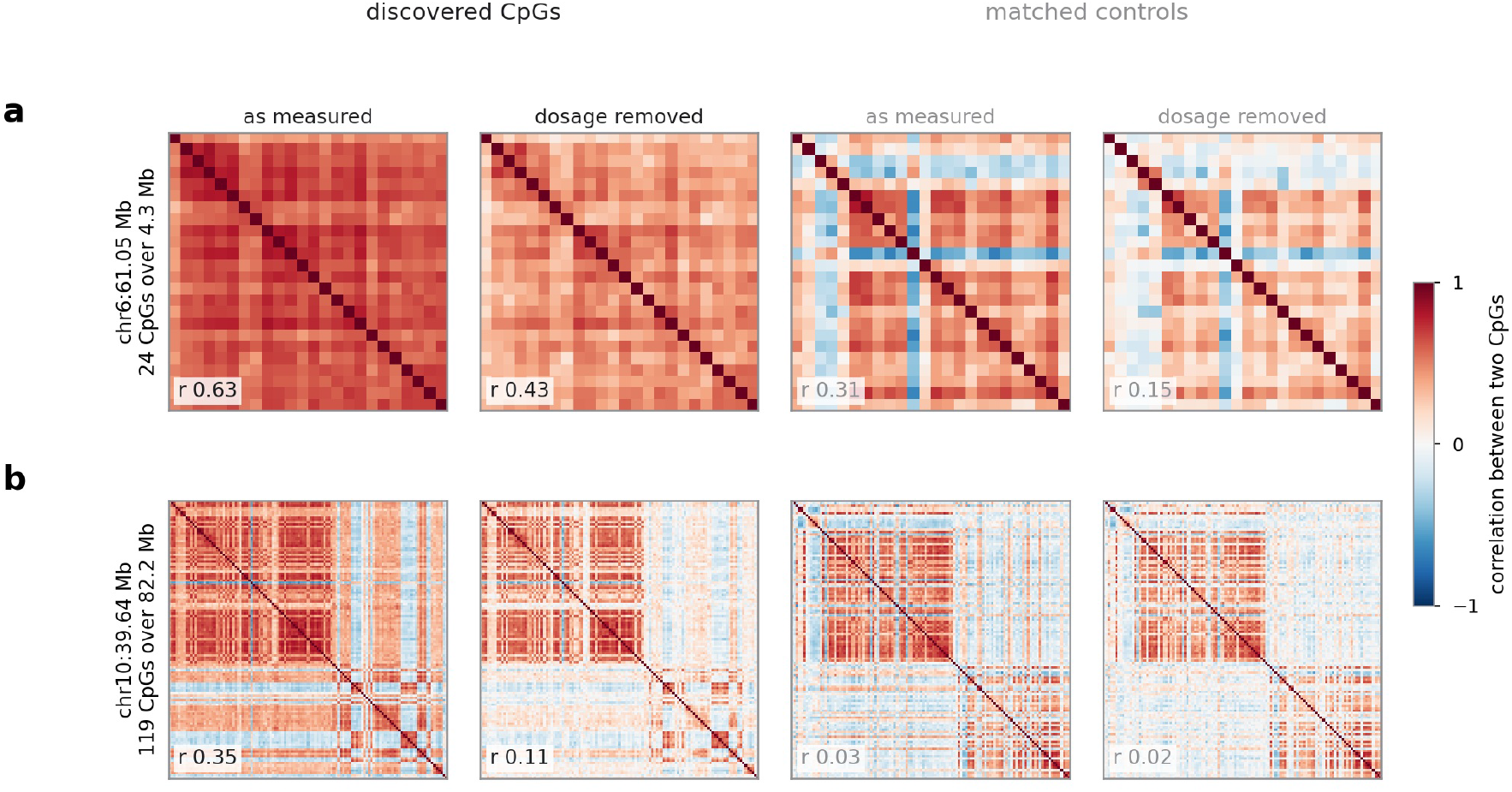
Co-methylation among the CpGs of one source variant. Correlation between the CpGs that share one source variant, z-scored and residualized on the association model’s sixteen covariates. Left, as measured and with the lead deletion’s dosage regressed out, so that the remaining correlations are assessed after adjustment for the genotype used to select the CpGs. Right, the same two treatments applied to CpGs that were tested at the same site but were not significantly associated, matched nearest at least 500 bp away, within 5 Mb, and measured in at least 80 percent as many individuals. With the dosage removed the discovered set correlates at 0.43 versus 0.15 at chromosome 6 and 0.11 versus 0.02 at chromosome 10. The CpGs share a variant and are physically dispersed: they span 4.3 and 82.2 Mb.

**Figure S14:**
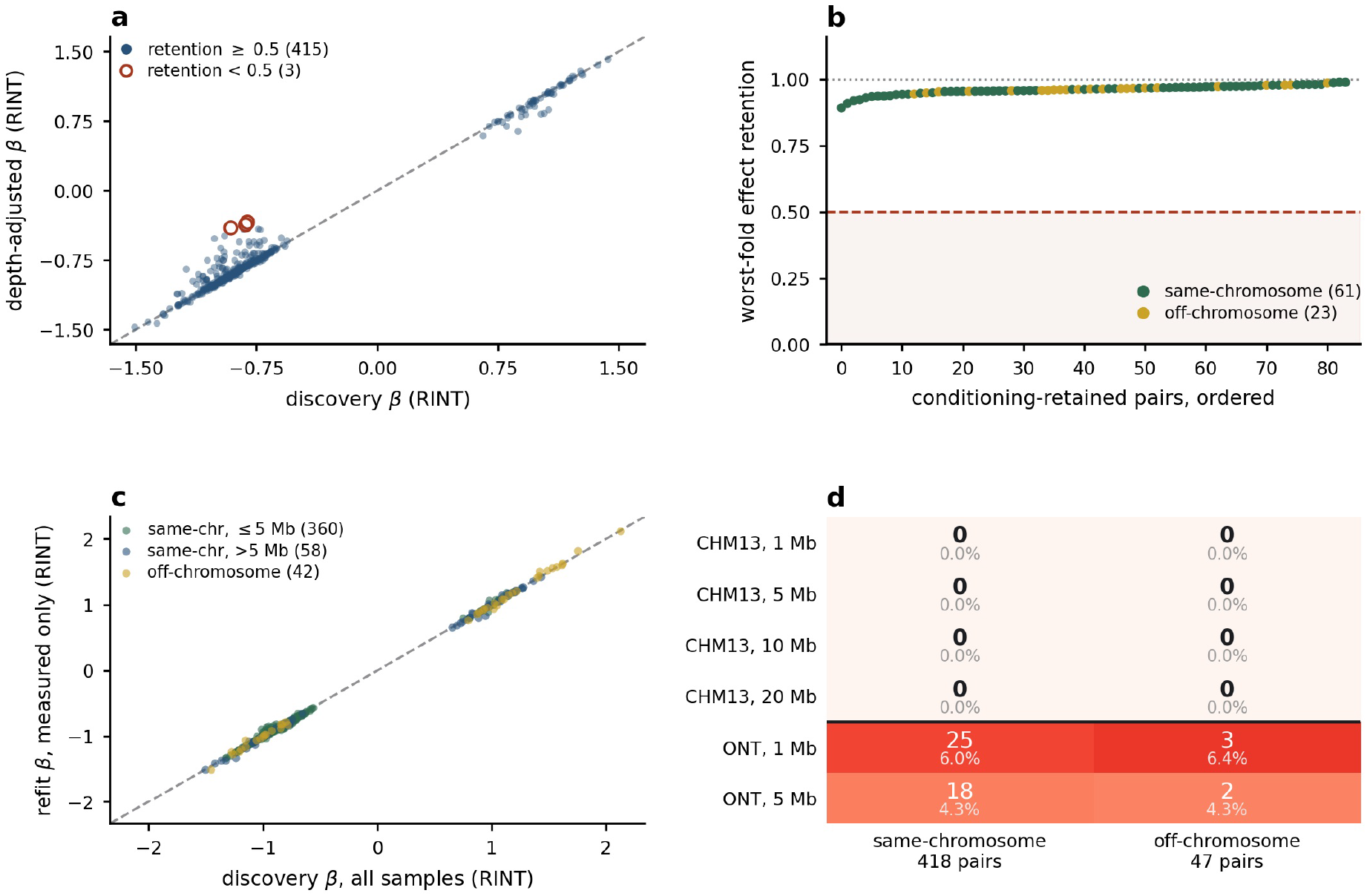
The discovery set under three refits, and conditioning on local SNPs. Effects in **a** and **c** are on the rank-inverse-normal scale; dashed lines mark the identity. **a** Per-sample CpG read depth added as a covariate. Median effect retention 0.997, all 418 pairs below *P* = 0.05 and significant under coverage weighting, 99.5% under a count-aware model. Coverage is associated with dosage at 379 of the 418 targets at *q <* 0.05. **b** Leave-one-carrier-out for the 84 conditioning-retained pairs of (d), showing the minimum effect retention across folds for each pair; the dashed line and shaded band mark 0.5. Every pair retains its effect direction in every fold; the minimum effect-retention ratio is 0.892. **c** Refit on genotype-complete samples; integer dosages equal the measured genotypes, dropping a median of 3, 8.5 and 13 samples per pair in the three approaches. Median retention 1.00 in every approach, sign agreement 460 of 460: the 418 same-chromosome pairs with 42 of the 47 off-chromosome discoveries. **d** Conditioning support for all 465 discoveries in two independent SNP panels at four distances from the source SV, as supported pairs and percentages. Seven pairs reach support, each in exactly one panel. Conditioning instead on the upstream elastic-net dosage score and its non-zero tags retains 84 pairs, at median *r*^2^ 0.346 of the observed dosage.

**Figure S15:**
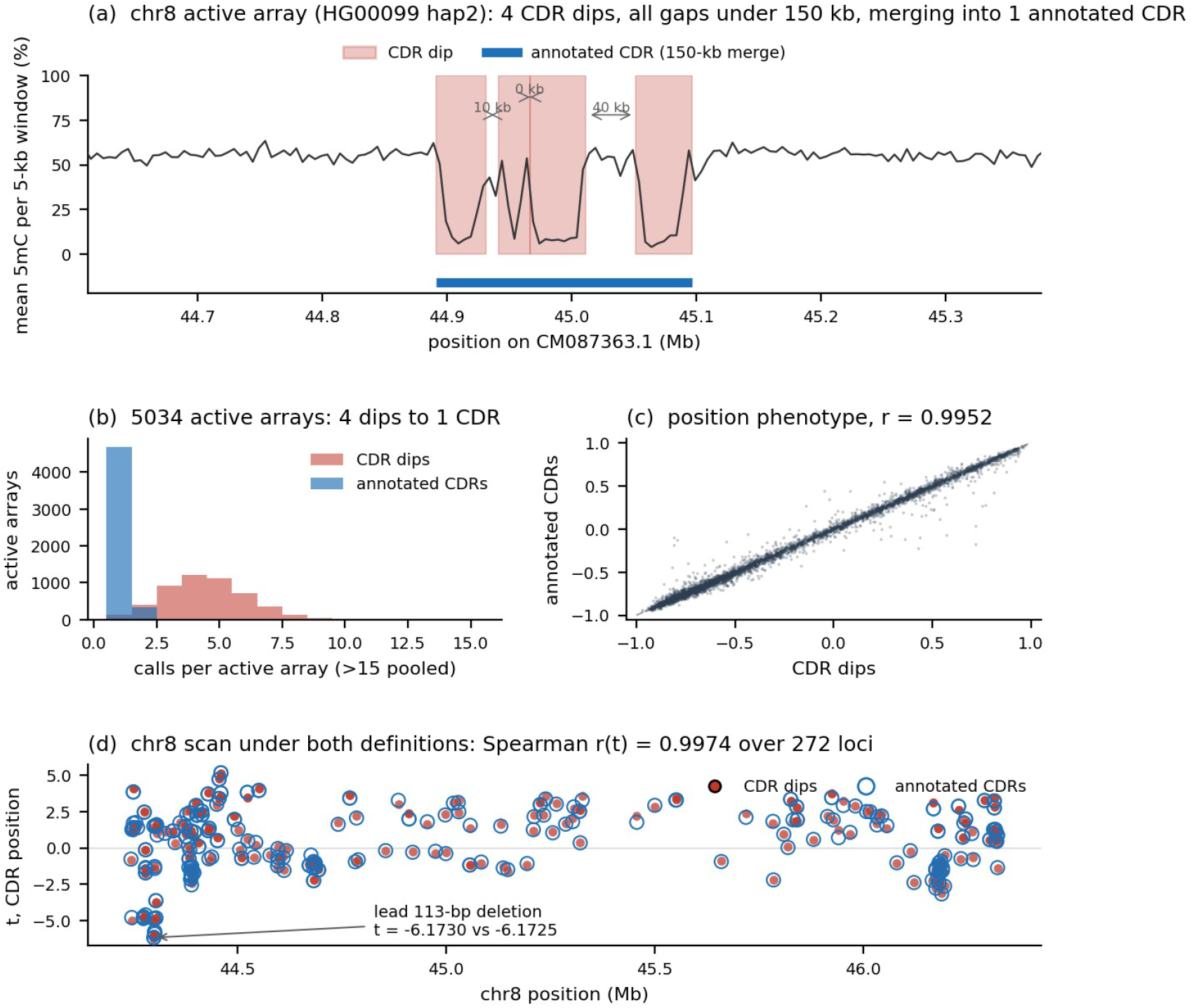
CDR calls, relative-position phenotype, and the chromosome-8 association analysis under two CDR definitions. **a** Four CDR dips on one chromosome-8 active array, gaps below 150 kb merging them into one annotated CDR. **b** Median CDR calls per array across the 5,034 active arrays of the call set, falling from 4 to 1. **c** Relative-position phenotype under the two definitions (*r* = 0.995). **d** Chromosome-8 association analysis under both definitions, the lead 113-bp deletion moving from *t* = *−*6.1730 to *t* = *−*6.1725 (CDR width: Supplementary Fig. S18).

**Figure S16:**
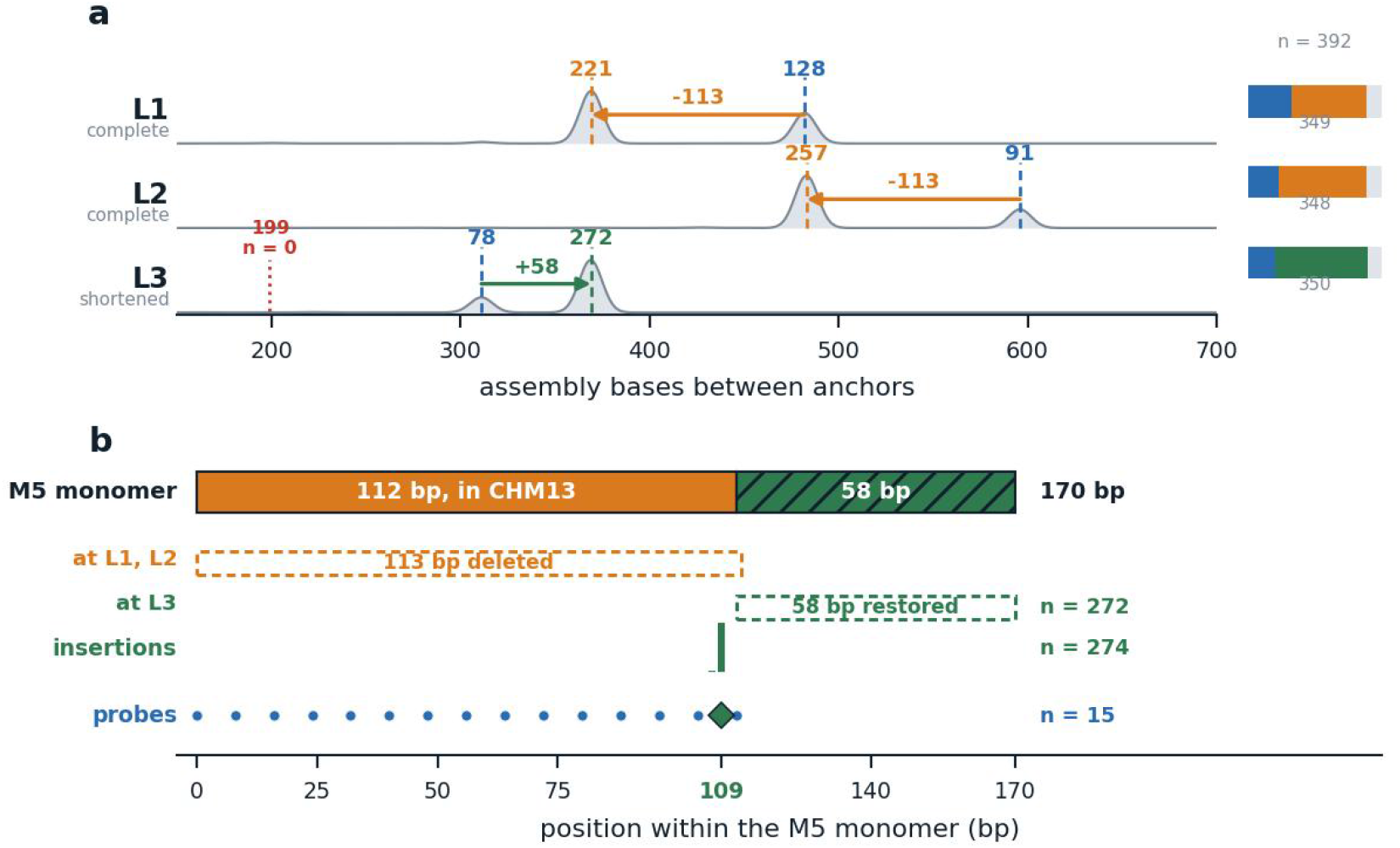
A shortened *α*-satellite monomer at three copies of the chromosome-8 arrangement. **a** Span at the three copies across 392 haplotype assemblies, with the outcome for each of the 392 assemblies shown at right. **b** The M5 monomer as a coordinate axis, showing the 113 bp absent at the first two copies, the 58 bp present at the third, insertion positions in 274 haplotypes, and 15 probes of known position. Methods; Supplementary Note 7.

**Figure S17:**
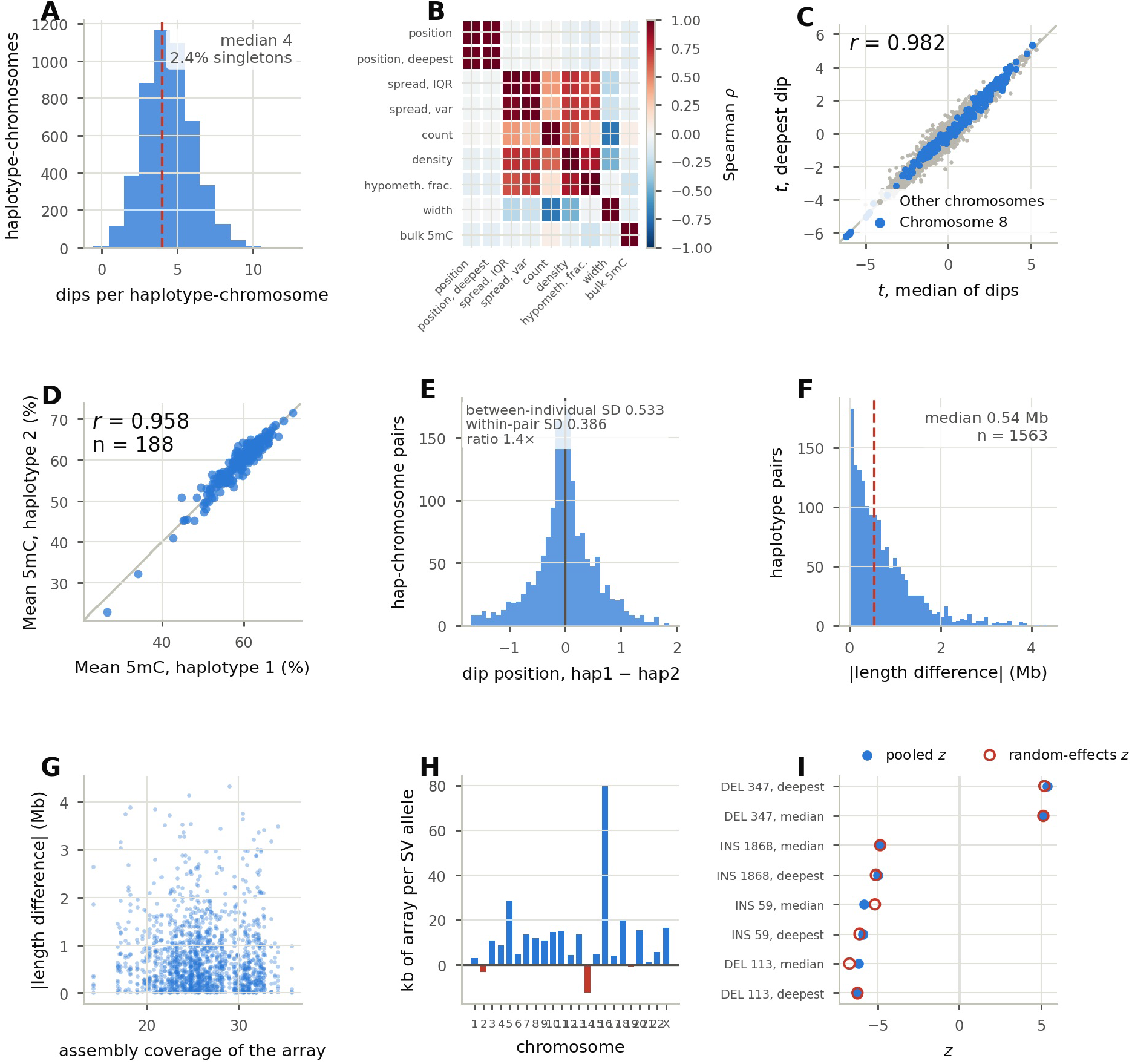
Nine measurement checks on the dip phenotype family. **a** Dips called per haplotype-chromosome over the 4,851 haplotype-chromosomes of the phenotype cohort: median 4, 2.4% singletons, maximum 12. **b** Spearman correlation among the nine tested phenotypes. The two position summaries reach *ρ* = 0.98; position against hypomethylated fraction, *ρ* = 0.11. Associations follow the phenotype correlation structure. **c** Association statistic per tested predictor using the median over all dips versus the single deepest dip as the CDR, *r* = 0.982 over 11,119 predictors. Blue points mark chromosome-8 predictors at both extremes and on the diagonal. **d** Mean 5mC of the active array (*n* = 188): between-individual spread (SD 6.0 percentage points) exceeds within-pair spread (SD 1.3), *r* = 0.958. **e** Dip position, haplotype repeatability: between-individual SD 0.539 against within-pair SD 0.390, a ratio of 1.4. **f** Active-array length, median difference 0.54 Mb over 1,563 pairs. **g** Assembly coverage against that length difference. Longer arrays are assembled from more sequence (Methods). **h** Change in active-array length per SV allele by chromosome, positive on 20 of 23. Sign and size vary across chromosomes. **i** Pooled against random-effects *z* for each of the 8 lead SV–phenotype associations. The two agree, maximum *I*^2^ = 0.35, and every lead association retains its effect direction.

**Figure S18:**
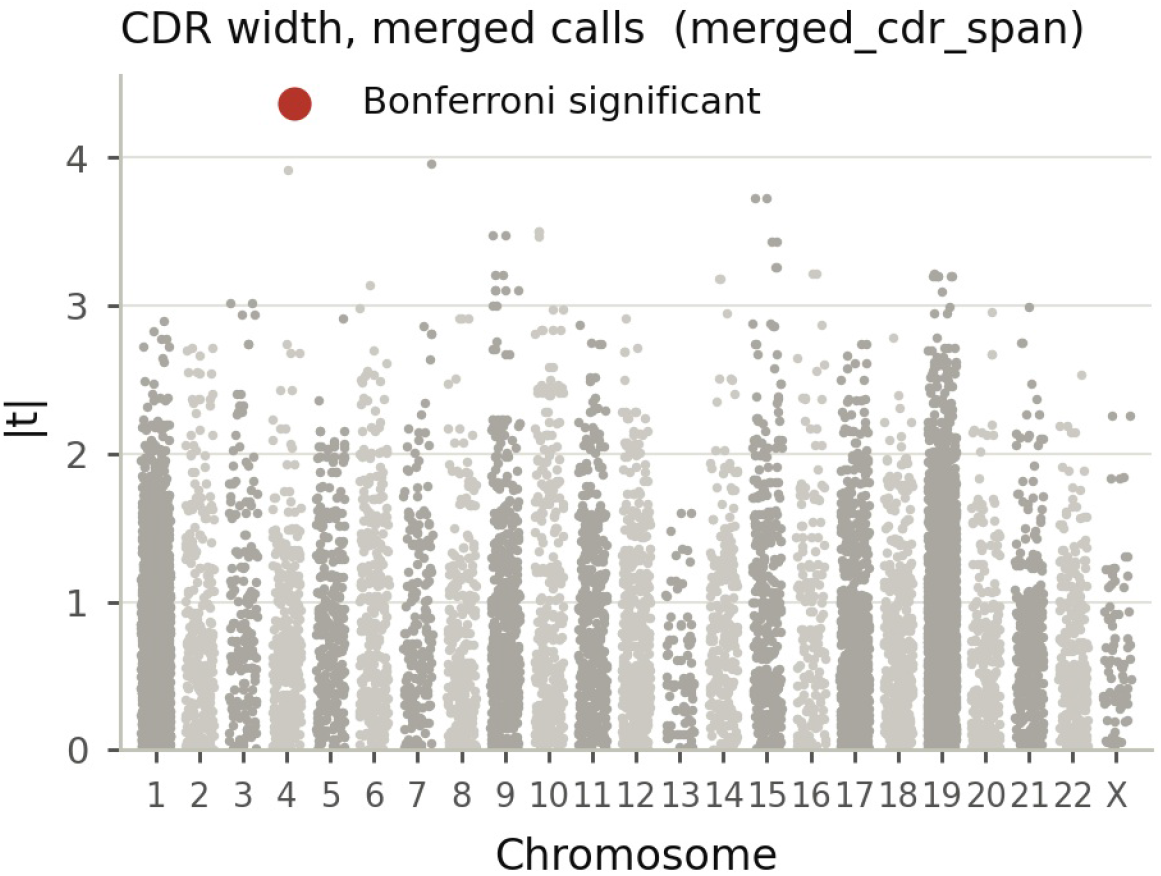
CDR width under the merged-call definition. Association of every centromeric SV locus with CDR width over the 23 tested chromosomes (11,119 loci, of an 11,129-locus catalog), on the genotypes, covariates and permutation procedure of Extended Data Fig. 1. The ordinate is *t*, with significance decided on the permutation *P* (Methods). Dips within 150 kb are joined into one region as in Gao et al. [2026], giving a median width of 215 kb against 30 kb for a single dip. The strongest association reaches *P* = 8.0 *×* 10*^−^*^5^, above the study-wide Bonferroni threshold (*α* = 5.6 *×* 10*^−^*^7^).

## Supplementary Notes

### Supplementary Note 1 SV-caller comparison inside active *α*-satellite HOR arrays

To choose a caller for the catalog, we compared three assembly-based callers on the same HG002 HPRC Release 2 haplotype assemblies against CHM13 v2.0 [Nurk et al., 2022], with every callset restricted to the 62.25 Mb of active HOR array in CenSat v2.1 [Altemose et al., 2022]. SVIM-asm v1.0.3 [Heller and Vingron, 2021] ran per haplotype; PAV v2.4.6 [Ebert et al., 2021], applied genome-wide by HPRC Release 2 [Lucas et al., 2026], ran in a default and a tuned configuration; dipcall v0.3 [Li et al., 2018a] ran at default parameters, with no -x and no -z. PAV and dipcall each align the assemblies themselves, so assemblies, reference and HOR restriction are held constant; the alignments are not. SVIM-asm reports the most SVs at every cutoff through *≥* 100 kb (Table S1): 6,517 at *≥* 50 bp against 4,793 (PAV default) and 2,506 (dipcall), and 49 at *≥* 100 kb against 2 and 0. At *≥* 500 kb every caller reports zero non-breakend calls.

The tuned PAV run (-r 50000,500000, inv_region_limit 5 Mb, sized for HOR cores above 1.2 Mb) called fewer SVs than defaults at every cutoff from 1 to 50 kb (Table S1), and both configurations call the same two variants of at least 100 kb.

Within the arrays dipcall reports no SVs on chr1, chr5, chr6, chr13, chrX and chrY, although chr1, chr5, chr6 and chr13 contain 1,659, 1,420, 1,330 and 920 dipcall indels *≥* 50 bp genome-wide. In high-identity arrays its alignment pileup either leaves the tandem repeats unaligned or GAP-filters the calls.

We applied the same SVIM-asm pipeline to the near-complete Q100 HG002 v1.1 maternal and paternal assemblies and compared the resulting calls with those from the HPRC Release 2 HG002 assemblies. In active HOR arrays the two callsets showed a positional agreement of 0.864 and Truvari [English et al., 2022] bench precision/recall/F1 of 0.842/0.824/0.833 (Table S2), a measure of reproducibility across assemblies of one individual. Against simulated SVs of known sequence, the same Truvari settings give precision/recall/F1 of 0.27/0.61/0.38 (Supplementary Note 13). HGSVC reported dipcall as the assembly-based caller with the highest false-positive rate [Logsdon et al., 2025].

We therefore used SVIM-asm for the catalog. The comparison is based on one individual, HG002, and the callers merge haplotypes differently, so differences in call counts should be interpreted qualitatively. SVIM-asm reclassifies events above its maximum SV size as breakends, 143 records in active HOR (*∼*71 unique pairs).

### Supplementary Note 2 Assembly contiguity limits centromeric SV detection

Centromeric SV calling requires a contig that spans the HOR array; where an assembly terminates inside the array, SVIM-asm cannot emit a variant. We compared the HiFi HPRC Release 2 assemblies against the haplotype-resolved ONT R9.4.1 assemblies released by the 1000 Genomes ONT Sequencing Consortium [Gustafson et al., 2024], produced with Shasta [Shafin et al., 2020]. We measured the ONT assemblies to be 25–40*×* more fragmented than HiFi (ONT 2,503–3,300 contigs per haplotype, N50 *≈*26–35 Mb; HiFi 74–120 contigs, N50 *≈*134–149 Mb; Table S3). On chromosome 9 this reduced the median per-haplotype SV yield *∼*6-fold (*∼*25 versus *∼*150 calls), with tandem-array classes most depleted (DUP:TANDEM 13.9*×* fewer, breakends 12.4*×* fewer, versus 3.8*×* for all classes). This depletion is consistent with *α*-satellite array collapse. For a given locus and individual, when near-T2T HiFi assemblies of the same samples resolve the array as a single contig, we recover the haplogroups. HOR SV maps therefore require HiFi (or comparably contiguous) assemblies.

### Supplementary Note 3 Locus mappability is the dominant determinant of imputation accuracy

We used two models to distinguish whether an SV is imputable from how accurately it is imputed. The first model, classifying all 192,363 catalog SVs as imputable (5.6%) or not (Table S5), distinguished the two groups with a 5-fold ROC-AUC of 0.989 and relied primarily on sequence-composition and chromosome features (GC content, active HOR span, satellite complexity), not allele frequency. In the second model, adding contig-level alignment features (mean MAPQ, fraction of secondary alignments, contig count, and edit distance at the locus) raised the held-out *R*^2^ of the imputation-accuracy model from 0.407 to 0.457. The fraction of secondary alignments became the single most important predictor (permutation-importance 0.133; Supplementary Fig. S3, Table S6), while the top sequence-composition feature (GC content over 10 kb) fell from first to fifth in importance. In grouped drop-column tests the mapping-quality family contributed the largest single-group loss (Δ*R*^2^ = *−*0.054, 2.2*×* that of GC), and the three feature families together explained roughly half the modeled variance. GC content is therefore largely a proxy for mapping ambiguity in repetitive sequence. In the Q100 HG002 assembly, we reproduced imputable SVs at a higher rate than unimputable ones (0.904 versus 0.848, +0.055), but a chromosome-block bootstrap CI included zero ([*−*0.011, +0.124]).

### Supplementary Note 4 Comparison with a reference-panel copying model (Beagle)

To compare imputation accuracy, we benchmarked the per-SV elastic net against Beagle 5 [Browning et al., 2018], a Li–Stephens copying model [Li and Stephens, 2003], on the identical SVs, pericentromeric SNPs and shared SNP set. Reconstructing SNP–SV phase from the assembly-to-reference alignments raised the SV–SNP *r*^2^ from 0.58 to 0.86 (SNP–SNP control 0.99), and held-out common SNPs imputed at *r*^2^ 0.93–0.97 as a positive control. We ran Beagle per centromere under five-fold cross-validation with a low-recombination map and effective population size *n_e_* = 2000, as `beagle ref=PANEL gt=FOLD map=MAP ne=2000 impute=true gp=true ap=true nthreads=4`, with leave-one-out affine recalibration of its posterior dosage, which we assessed in raw, affine and isotonic dosage-*R*^2^ forms at a minor-allele-count floor of 20.

Across all 23 centromeres the two methods gave comparable accuracy (centromere-level block bootstrap, mean gap *−*0.003 in Beagle’s favor, 95% CI [*−*0.013, +0.008], *P* = 0.59; monophyletic class *P* = 0.96; elastic net ahead in 12 of 23 centromeres at the allele-count floor and 13 of 23 in the monophyletic class, including a single-centromere *∼*1.3*×* lead on chromosome 17 in Pearson *r*^2^, Table S7). The elastic net was more robust than Beagle when each superpopulation was held out in turn. Beagle requires the assembly-co-phased panel, built from the same assemblies it imputes, while the elastic net uses SNP genotypes alone.

### Supplementary Note 5 Expression and splicing cis-QTL from ascairn k-mer haplogroups and StV repeat variants

To test whether the QTL signal depends on how variants are defined, we repeated the QTL mapping with a genotyping route that reads centromere haplogroups directly from short-read k-mers (ascairn [Shiraishi et al., 2026]). We called haplogroups with ascairn v0.3.0 against the published ascairn_paper_2025 reference panel of assembled active-HOR haplotypes (HPRC, HGSVC and T2T assemblies) under the distributed defaults. For each chromosome the workflow reports the most likely active-HOR haplogroup pair, the nearest proxy haplotypes from the panel, and marker-level probability tables. ascairn types directly from short reads and bypasses the imputation step, so we typed and tested *n* = 728 MAGE individuals, exceeding the 630 in the main analysis. Using covariates matched to the main analysis, we tested haplogroup dosages for associations with expression and splicing, including chromosome X, and identified 66 cis-eGenes and 49 cis-sGenes, 46 and 35 of them within the main-analysis 86 and 74 (Table S8; Supplementary Fig. S4a; *λ* = 1.016 for expression and 1.062 for splicing; 101 unique cis genes). Of these 101, 73 lie among the 141 cis genes of the main analysis, and 64 genes are common to the main, ascairn and StV routes, whose union spans 200 genes.

#### Concordance under an *α*-satellite HOR variant definition

StV [Altemose et al., 2022] called 17,786,321 higher-order repeat instances of 22,652 types across the 384 phased haplotype assemblies, 2,992 of them common. Tested in a QTL analysis that shared the assemblies, the SNP panel, the elastic-net imputation, the 630 individuals and the tensorQTL configuration of the main analysis (Methods), these dosages identified 78 cis-eGenes and 63 cis-sGenes, of which 54 of the 78 and 42 of the 63 fall within the main-analysis 86 and 74 (Supplementary Fig. S4b).

### Supplementary Note 6 Distal cis associations and controls

We asked whether distal cis and trans associations survive controls for tag SNPs and paralogs.

#### Two distance metrics in cis

Every centromeric SV is anchored at its chromosome’s centromere midpoint for cis-region construction, so the distance the QTL mapping reports is distance to the centromere. Because an imputed dosage is a linear function of its tag SNPs, we also computed distance to the nearest tag SNP of its elastic-net model, restricted to the 3,905 of 3,949 model SNPs (98.9%) present in the 1000 Genomes phased panel; SNPs absent from the panel are represented as constant columns and cannot contribute. Of all 166 significant cis gene associations across the two imputation ensembles, 35 lie *≥* 10 Mb from the centromere, and 14 lie *≥* 10 Mb from the centromere and from every tag SNP, at 11 independent loci, with distances ranging 12.2–132.4 Mb.

We refit every significant cis pair conditioning on the model’s tag SNPs (Supplementary Fig. S7). The refit reproduces the main analysis (Pearson *r* = 0.997 between refit and original *—* log_10_ *P* ; significance recovered in 430 of 430 pairs). Conditioning on all of the model’s local tag SNPs is degenerate by construction, so we classified each fit by the *R*^2^ of the dosage on its conditioners (informative *<* 0.90, marginal 0.90–0.99, degenerate *≥* 0.99), and excluded degenerate fits. Conditioning on all local tag SNPs leaves 42 of 124 testable eQTL pairs (33.9%) and 45 of 100 sQTL pairs (45.0%) significant; conditioning on the single nearest tag SNP leaves 190 of 238 (79.8%) and 179 of 188 (95.2%). All 14 genes distal under both definitions survive on informative fits (*R*^2^ 0.000–0.142). Conditioning leaves the three chromosome-20 associations intact, shifting them from *P* = 4.16 *×* 10*^−^*^9^ to 3.58 *×* 10*^−^*^9^ (*TTPAL*), 5.42 *×* 10*^−^*^7^ to 9.78 *×* 10*^−^*^7^ (*PLTP*) and 5.96 *×* 10*^−^*^6^ to 6.07 *×* 10*^−^*^6^ (*NEURL2*).

#### The paralog control

GRCh38 cross-mappability covers only paralogs present in GRCh38, so a complete-assembly search, also applied to the trans candidate *DUSP22* below, was run on the 14 genes distal under both distance metrics, asking whether each has a high-identity copy in the pericentromere of its own chromosome; every query recovered its own locus at identity *≥* 0.98. Thirteen have no such copy. The exception is *WHAMMP1* on chromosome 15, a transcribed processed pseudogene and the most significant of the 14 (*q* = 7.8 *×* 10*^−^*^4^). It has six high-identity copies on chromosome 15 (99.49% at 28.26 Mb, 96.0–96.4% at 20.33, 20.37, 26.51 and 26.55 Mb, 93.4% at 80.67 Mb), two within 3 Mb of the *α*-satellite array and all at mapping quality 0. We therefore exclude it, leaving 13 distal genes at 10 loci. None of the three chromosome-20 genes has an additional copy anywhere on chromosome 20.

We computed genomic-control *λ* from the per-gene Beta-approximated permutation *P* value, pooled across the 23 chromosomes. For expression it is 1.04 in the whole-chromosome region and 1.12 in the *±*10 Mb region; for splicing, 1.11 and 1.17 (Supplementary Fig. S8), with Storey *π*_0_ [Storey and Tibshirani, 2003] of 0.98, 0.95, 0.95 and 0.93 respectively.

#### Trans associations

Across 1.26 *×* 10^9^ SV–phenotype pairs tested in trans, the four controls (multiple testing, cross-mappability, conditioning and the paralog check) remove all 46 target genes.

Over the union of the two imputation ensembles (8 eQTL and 38 sQTL target genes, 46 in total), Benjamini–Hochberg at *q <* 0.1 applied across all trans tests retains 12 trans target genes and Bonferroni retains 2, whereas a fixed *P <* 5 *×* 10*^−^*^8^ threshold expects *∼*63 false positives across these tests, with an empirical false-discovery rate of 59.6% under permutation calibration.

#### Both Bonferroni-significant genes have paralogs

In the GRCh38, GENCODE v26 cross-mappability tables [Saha and Battle, 2018a,b], *HSD17B7* and its pseudogene *HSD17B7P2* show a symmetric cross-mappability of 999.5 k-mers. *HSD17B7P2* lies inside the chromosome-10 pericentromere, and in our analysis of that table the cross-mapping is 30.8-fold stronger than with the pericentromere of any other chromosome. *DUSP22* has a 63.9 kb copy, *DUSP22B*, that is 99.86% identical, lies 0.82 Mb from the chromosome-16 *α*-satellite array in CHM13, and is absent from GRCh38, which instead maps that sequence onto chr6:287,496–356,341 at mapping quality 60. Short reads from the unreferenced copy are assigned to chromosome-6 *DUSP22*, so chromosome-16 pericentromeric SVs linked to that copy appear as trans regulators. The lncRNA overlapping the *DUSP22* promoter (ENSG00000287265) has a 99.78%-identical copy at the same chromosome-16 locus and shows the same association, whereas *EXOC2*, a chromosome-6p25.3 neighbor 0.2 Mb away with no chromosome-16 paralog, has no chromosome-16 SNP below *P* = 1 *×* 10*^−^*^5^. The same mismapping mechanism accounts for an estimated 6.1% of trans-eQTLs in this cohort [Aqil et al., 2026].

Of the 46 target genes, 12 remain after Benjamini–Hochberg, 2 after Bonferroni, 1 after cross-mappability, which removes *HSD17B7*, 1 after conditioning, and 0 after the non-reference-paralog check, which removes *DUSP22*. No trans association survives all four controls. *POU2F1* fails Benjamini–Hochberg, fails Bonferroni, and survives conditioning in 0 of 6 pairs; *MCOLN2* fails both thresholds and cross-maps at 20.5 with *PXMP4*.

### Supplementary Note 7 Supporting analyses for the chromosome-8 centromere-dip-position locus

The Li–Ji effective number of independent phenotypes is 6.0, and the significant lead variants reach the 10*^−^*^7^ permutation floor and the significant set is unchanged when correction uses seven, eight or ten phenotypes. We asked whether this association is robust, whether the deletion can be distinguished from linked SNPs, and what sequence underlies it.

#### Robustness of the chromosome-8 association

##### A chromosome-8 locus is associated with CDR position

The lead deletion lies in a 213.6 kb interval at chr8:44.25–44.46 Mb.

It contains every study-wide-significant chromosome-8 lead from all eight CDR phenotypes. Within it, five distinct genotype vectors on four SVs produce nine significant (genotype-group *×* phenotype) associations (Extended Data Table 1; Figure 4a, b): four vectors under *both* position phenotypes, and a 1,868-bp insertion under the deepest-dip phenotype alone; the insertion at chr8:44,302,147 reaches the threshold under two independently derived genotype vectors. Outside this interval, leads on chromosomes 5, 9 and 19 also reach study-wide significance under the canonical covered-length frame; each loses it in the robustness analysis (Extended Data Fig. 1).

##### The chromosome-8 association is concordant across five superpopulations

It is permutation-calibrated at 10^7^ draws over the complete catalog, stable when the array endpoints are modeled, homogeneous across five superpopulations (concordant in all five, *t* from *−*2.06 to *−*4.58; inverse-variance meta-analysis *z* = *−*6.56, *P* = 5.3 *×* 10*^−^*^11^; Cochran *Q P* = 0.50) and specific to dip position among eight phenotypes. The association identifies the locus but does not distinguish among its linked variants. At *r* = 0.95 the 113-bp deletion and its linked chromosome-8 pericentromeric variants share the same signal in this cohort, and dip position is measured relative to the annotated array.

##### The locus has a single signal

The 113-bp deletion and its tag SNP show similar associations with CDR position (*P ≈* 10*^−^*^7^ for each). In African-ancestry samples linkage partially breaks down (*r*^2^ = 0.67) and the SNP survives joint fitting while the deletion loses significance (*P* = 0.033, *n* = 41); in East Asian and European samples the pair is collinear. The interval contains a single independent signal. Conditioning each pre-specified lead on the top lead leaves three with a nominal conditional association (*P* = 0.019–0.041), the strongest the 347-bp deletion at chr8:44,460,941 (*|r|* = 0.62). Calibrated against the set screened, the best of these is *P* = 0.10 over the pre-specified leads and *P* = 0.61 over all 69 genotype vectors in the interval.

#### Specificity to dip position

##### The chromosome-8 association is specific to dip position

Of the four aspects of CDR organization measured (Extended Data Table 2), only position shows a significant association. The chromosome-8 leads show no association with dip spread, dip count, dip width, the hypomethylated fraction or bulk methylation (lead deletion *P* = 0.59 for hypomethylated fraction, 0.15 for mean 5mC, 0.52 for dip width, 0.24–0.96 for dip count; Extended Data Fig. 1).

Dip position is dosage-dependent at the chromosome-8 leads: the median relative dip position is 0.82, 0.71 and 0.55 across the three dosage classes of the lead deletion (Figure 4c).

##### Chromosome-8 array length is independent of dosage

The chromosome-8 locus alone has this property (*|t| ≤* 1.00, *P ≥* 0.32). Total SV burden predicts array length (*t* = +11.95), no burden association remains significant once length is in the model (Figure 4e), and length variation is uncorrelated with assembly coverage (*r* = *−*0.03, *n* = 181).

For mean 5mC across the array (Supplementary Fig. S17), the SV association analysis shows little genomic inflation (*λ* = 0.948, median *P* = 0.49), and aggregate burden, associated with mean 5mC at *P* = 1.8 *×* 10*^−^*^9^, reaches *P ≈* 0.08 after array-length adjustment, with a smaller individual-level component at *P* = 0.003. Contig count predicts mean 5mC and array length (*t* = +4.5, +6.3) and explains *R*^2^ = 0.02 of burden.

In analyses partitioned by individual and chromosome, dip position has an intraclass correlation of 0.000 and mean 5mC of 0.91, and adjusting either for array length leaves both unchanged; dip position can differ between homologs within an individual, consistent with a *cis*-acting effect.

Partitioning variation in the same way distinguishes two contributions of array length: it accounts for 54% of the between-superpopulation difference in the hypomethylated fraction and 42% in dip density, the two phenotypes that divide by it, and none of the difference in mean 5mC, which remains an individual-level trait that differs among superpopulations after adjustment (*R*^2^ = 0.083, *P* = 0.004).

No chromosome-8 SV reaches the threshold at any of the three interval widths of the localmethylation test (Supplementary Note 11), on a set that tests all 272 of its SVs at *n* = 130. The mediation analysis provides no evidence that local methylation level explains the chromosome-8 association with dip position. Across all four leads the direct effect equals the total, the SV-to-methylation step has *P* = 0.39–0.89, and the indirect effect’s confidence interval spans zero.

##### Three further loci associate on arrays whose length or boundaries vary with dosage

Leads on chromosomes 5, 9 and 19 rank highest outside the chromosome-8 interval, though the chromosome-19 lead loses significance once the array endpoints are added as covariates. Their arrays change length with dosage, and normalizing by array length cannot distinguish a shift in dip position from a change in total array length. The chromosome-5 lead is the only one whose association weakens after adjustment (*t* = +5.32 *→* +4.09, *P* = 1 *×* 10*^−^*^6^ *→* 4.2 *×* 10*^−^*^5^), and the basis of this association remains unclear.

#### Cis effects on the haplotype with the deletion

##### Dip position differs between homologs by deletion status

In the 15 individuals heterozygous for the lead deletion, the two homologs of a single nucleus have relative dip positions of 0.516 and 0.819 (paired difference *−*0.309, Wilcoxon *P* = 3.0 *×* 10*^−^*^4^; the homolog with the deletion closer to the center of the array in 13 of the 15). Using the two centromere groups in place of the deletion gives 27 heterozygotes and the same result (difference *−*0.253, Wilcoxon *P* = 4.9 *×* 10*^−^*^7^, 24 of 27), equal in size to the difference between unrelated individuals.

##### Dip position differs by SV-defined centromere group

Grouping haplotypes by neighbor-joining on cosine distances over the chromosome-8 SV matrix gives two SV-defined centromere groups, with the 113-bp deletion present on every haplotype in one group. Nineteen haplotypes in the other group also have the deletion, allowing a within-group comparison. Using assembly sequence between unique flanking 31-mers, we genotyped the deletion and found a mean relative dip position among carriers of 0.834, indistinguishable from the 64 non-carriers of the same group (0.837). The deletion’s effect within a group is estimated at *−*1% of the between-group difference (bootstrap 95% CI *−*22% to +10%), and dip position is associated with the SV-defined group. Because every haplotype in the first group has the deletion, only sufficiency on the alternative background is testable, not necessity. A further 12 carriers in the same group had no dip called, at a rate indistinguishable from non-carriers (61% against 73%, Fisher *P* = 0.26).

#### Sequence at the chromosome-8 locus

##### Class assignment of the partial monomer

We assigned the 112 bp monomer to a class by globally aligning its first 112 bases to the first 112 bases of each class consensus, with identity taken as matches over alignment length. Its identity to the M5 consensus exceeded that of all 927 other-class monomers among 1,000 drawn from the same array (*P* = 0.001); in two additional null comparisons a 112 bp prefix reaches a median identity of 0.97 to its best class and the observed identity difference from the second-best class lies below the null median. The prefix-based assignment identifies a monomer’s own class in 94.2% of the 1,000 draws and in every M5 draw. Dvorkina et al. [2021] previously reported a partial monomer of this length in this array.

##### The monomer across haplotype assemblies

We measured the span between fixed anchors at each of the three copies in 392 HPRC Release 2 haplotype assemblies (Methods). At the first two copies the two states differ by 113 bp, which spans the 112 bp monomer and one adjacent base, and CHM13 has the longer state at both (482 against 369 bp, 128 and 221 haplotypes; 596 against 483 bp, 91 and 257). At the third copy the states differ by 58 bp and CHM13 has the shorter one (311 against 369 bp, 78 and 272). The 199 bp span expected if the monomer were absent at the third copy was not observed in any of the 350 resolved haplotypes. We resolved the three spans in 349, 348 and 350 of 392 haplotypes, respectively. Aligning each extracted span to the reference span places the 58 bp within the shortened monomer in all 274 haplotypes with the longer state, at monomer position 109 in 267 of them and 107 in the remaining 7, and none of the flanking M2 or M3.1 monomers contains it; the shortened monomer occupies 112 of the 311 bases of the measured span. The 58 bp matches M5 monomers of this array from position 109, and M5 accounts for the 100 highest-scoring placements. Since 112 and 58 sum to 170, the modal M5 length of this array, the two states at the third copy correspond to a shortened and a full-length monomer. Probes of the same length drawn from full-length M5 monomers at known positions each recovered the position they were drawn from, which establishes that the placement resolves position within a monomer (Supplementary Fig. S16). At the 113-bp deletion itself, assembly sequence agreed with the SVIM-asm genotype in 330 of the 337 haplotypes with an unambiguous allele assignment (97.9%).

##### Shortened monomers across the reference

To compare the chromosome-8 count with other centromeres we applied the same decomposition and the same length threshold to all 26 active arrays of the reference, giving 255,245 monomers, and tested the class distribution of the pooled short monomers against class abundance by permutation with 200,000 draws. The class distribution differs from that expected from monomer abundance (*P* = 5 *×* 10*^−^*^6^), with class M5 at 7.2 times its expected count and M4 at 7.6.

##### Operational definitions

We classified monomers shorter than 150 bp as truncated. On the chromosome-8 array the four such monomers, including the three partial-monomer copies, fall between 112 and 124 bp and the next shortest measures 161 bp. We identified CENP-B boxes as matches to NTTCGNNNNANNCGGGN on either strand. Four of the array’s 5,263 boxes have adenine at the first position of the motif, and two of the three truncated copies contain one each.

#### Dip measurement

##### Dip calling and dip counts

Calling the CDR within each sample’s active HOR array [Gao et al., 2026] gives *∼*22,000 dips, 57 per haplotype across a median of 13 assembled centromeres and *∼*4 per centromere, and merging dips within 150 kb gives a mean of 1.07 annotated CDRs per active array (93.1% with exactly one). A concurrent survey reported one kinetochore site in 94% of centromeres genome-wide and in 98% of chromosome-8 haplotypes, each spanning a mean of 4.3 methylation dips [Gao et al., 2026]. The position association is retained with a single main dip per haplotype (*P* = 7.5 *×* 10*^−^*^4^) and with the deepest dip (*t* = *−*6.19). Over the whole centromeric satellite block the same caller and parameters produce *∼*324,000 dips (median 880 per haplotype, *∼*50 per centromere), against a literature expectation of one to three; at thresholds 0.5/0.5 the R10.4.1 subset gives 2.2–2.5 dips per centromere.

##### Chromosome-specific dip position

Where the dip falls along the array differs by chromosome: 19 of the 23 chromosomes place it preferentially on one side, and all five acrocentrics place it on the p side (Supplementary Table S9). On chromosomes 6, 10, 12, 19 and X the dips resolve into two preferred positions, recovered in independent halves of the cohort. The lower falls between 0.07 and 0.19 on all five and the upper between 0.74 and 0.94 on chromosomes 6, 10, 12 and 19, placing both within a few hundred kilobases of an array end (Supplementary Table S10).

##### Modification-call filtering and ONT chemistry

With methylation computed from 5mC calls alone (Methods), dip counts agree between the R9.4.1 and R10.4.1 cohorts (rate ratio 1.04, 95% CI 0.99–1.10; array methylation *P* = 0.15 adjusting for ancestry).

##### Trans association analysis

We tested each locus against every off-chromosome phenotype, 2,045,280 trans tests in 192 chromosome–phenotype pairs. Because a Bonferroni threshold at that test count (2.4 *×* 10*^−^*^8^) is below the minimum *P* value attainable with the available permutations, the main statistic is the global maximum *|t|* over 100,000 permutations shared across the whole panel. The analysis is calibrated (median *λ* = 1.013; 19,852 sub-threshold candidates at *P ≤* 0.01 against 20,453 expected), its cis positive control places the chromosome-8 leads at the top of 88,952 tests, and every trans test has family-wise error *≥* 0.05 and *q ≥* 0.05, the strongest being an insertion at chr20:28,704,151 against chromosome-21 dip width (*P* = 4.1*×*10*^−^*^7^, familywise error 0.14).

### Supplementary Note 8 Supporting analyses for the biobank-scale disease screen

The score test gave conservative or well-calibrated results relative to the exact Firth fit within each carrier-count bin and predictor set.

#### One hundred seventy associations, most of them protective

At *q <* 0.05, 170 SV– phecode associations are significant, spanning 61 distinct SVs and 27 phecodes, with a further 85 associations at 0.05 *≤ q <* 0.10 (Figure 6a).

Of the 170 associations, 110 were protective. The 61 significant SVs have a median cross-validated imputation *R*^2^ of 0.23, with 38 below 0.30 and 11 at 0.45 or better (accounting for 45 associations).

Chromosomes 11 and 19 contribute 1,022 and 1,058 SV–phecode pairs at *P ≤* 10*^−^*^4^, of which two and none, respectively, are significant, compared with 134 of 407 for chromosome 16.

A set of 3,097 pericentromeric SVs with no summary-level signal shows no association at *q <* 0.05 and none at *q <* 0.10 across 7,485,449 tests (Figure 6b). Its smallest *P* is 4.5 *×* 10*^−^*^8^ against 1.3 *×* 10*^−^*^7^ expected for the minimum of that many tests under the null, a value reached by chance with probability 0.29. These predictors are imputed as well as the discovery predictors, at a median cross-validated *R*^2^ of 0.30 against 0.21. The procedure is therefore calibrated on pericentromeric dosages, while the discovery predictors give 99 significant associations from a comparable 8,256,472 tests.

The 3,097 negative-control predictors occupy 11 chromosomes, including 1,884 on chromosome 19 and none on chromosomes 16 or X. We selected them for a null result in the summary-level association analysis (Extended Data Fig. 3).

Twenty-eight SV–phecode pairs from the range 0.05 *≤ q <* 0.10 are significant in more than one ancestry group, over 27 SVs at 14 loci, and all 28 survive restriction to unrelated participants.

#### Strong quantitative-trait associations are consistent across ancestry groups

Of 755,930 SV–trait pairs, 56,169 are consistent across ancestry groups and 589,607 underpowered. Among SV–trait pairs reaching meta *P <* 10*^−^*^4^, consistency runs from 102 of 104 systolic blood-pressure pairs (98.1%) and 253 of 273 heart-rate pairs (92.7%) to 70 of 83 diastolic pairs (84.3%) and 67 of 89 hip-circumference pairs (75.3%) (Extended Data Fig. 2c). On the same anthropometric trait the negative-control predictors are consistent in 166 of 300 SV–trait pairs (55.3%), against 239 of 308 (77.6%) for discovery predictors, and the comparison gives OR = 2.80 (*P* = 6.7 *×* 10*^−^*^9^). Strong quantitative-trait associations are therefore more consistent across ancestry groups than those of the negative-control predictors.

All 11 chromosome-X associations at *q <* 0.10 remain significant under each of the three ploidy models.

#### Fine-mapping

We fine-mapped every candidate association against its own tag SNPs in the unrelated subset (*n* = 291,668), producing 476 fits under two panel constructions and three methods (Methods). The imputed SVs are strongly correlated with their local SNPs: the multiple *R*^2^ of an SV on its own SNP panel has median 0.900, and 0.807 when the SNPs that built the dosage are excluded.

#### Credible-set composition

Among the single-SV credible sets that contain an SV, 12 contain the SV alone and 42 contain it together with a SNP outside its imputation predictors, while 7 contain it with only its own imputation predictors, which cannot distinguish the SV dosage from the SNPs used to predict it. We included five negative-control predictors in four fits, and two of those fits placed a control SV in a credible set, showing that credible-set assignment can arise without signal. Imputation accuracy is highest where an SV is included in a credible set (median 0.314) and lowest where the fit does not resolve (median 0.204), with accuracy and panel collinearity positively correlated (*r* = +0.179).

Of 46 localization instances, 15 arise under both panel constructions, 21 occur under the single-SV panel alone and 10 under the joint panel alone. Seven of the 10 are candidates on chromosomes 16 and X with marginal support from *P* = 1.2 *×* 10*^−^*^6^ to 6.2 *×* 10*^−^*^13^ that have higher posterior support than other SVs in the model; of the three outside the candidate set, a chromosome-16 SV with alcohol abuse and dependence reaches joint posterior 1.00 at marginal *P* = 0.94 within a cluster of collinear SVs, and two show marginal signal (*P* = 2.2 *×* 10*^−^*^8^ and 2.4 *×* 10*^−^*^6^) at negligible posterior.

### Supplementary Note 9 Measurement availability at candidate CpGs depends on carrier status

Because the methylation-QTL analysis estimates each CpG effect among individuals with a measurement, we tested whether measurement depended on genotype at the associated SV.

Supplementary Fig. S11 measures this dependence in 150 HPRC Release 2 individuals with assembly-derived genotypes, all distinct from the 124 discovery individuals. The per-pair slope of measurement presence on carrier dosage is negative in 302 of the 354 testable discovery pairs (85.3%), against a matched null of twenty control CpGs per discovery CpG (6,280 pairs over 4,715 control CpGs) whose rate falls below nominal under one shared Benjamini–Hochberg family. Measurement availability is associated with the sign of the discovery effect (Fisher odds ratio 15.55, *P* = 8.9 *×* 10*^−^*^26^). The target CpGs lie outside the deleted interval of their source SV in 465 of 465 pairs, so missing methylation calls cannot be explained by deletion of the target CpG. The assayed pericentromeric region spans 6.8 Mb per contig, and this window excludes the more distal pairs at which the discovery effect was hypermethylating.

A within-individual test comparing the between-haplotype methylation difference of SV heterozygotes with that of homozygotes detected an association at *∼*40% of the SVs shared between the methylation and expression analyses, a pattern that R9.4.1 5mC miscalling would also produce.

We estimated per-CpG effects in the discovery cohort alone because estimates from the HPRC cohort would include bias from differential measurement availability.

### Supplementary Tables

**Table S1:** SV-caller comparison inside active HOR arrays. (Supplementary Note 1). Counts from three callers for benchmark individual HG002 (HPRC R2 freeze_2 v1.0.1) across the 62.25 Mb of CHM13 active *α*-satellite HOR array. SVIM-asm in our configuration, PAV at defaults and tuned, dipcall at defaults and restricted to its own confident region (conf. BED). SVIM-asm reclassifies events above its maximum SV size as breakends, and breakends (BNDs) are counted separately. dipcall emits INS and DEL, with sizes above 500 kb left unevaluated (–).

|  | SVIM-asm v1.0.3 |  | PAV v2.4.6 |  | dipcall v0.3 |  |
|---|---|---|---|---|---|---|
| Size class | DEL+INS+INV+DUP | BND | default | tuned | default | conf. BED |
| ≥ 50 bp | 6,517 | 143 | 4,793 | 4,690 | 2,506 | 2,404 |
| ≥ 100 bp | 6,461 | 143 | 4,767 | 4,664 | 2,498 | 2,396 |
| ≥ 500 bp | 3,788 | 143 | 3,025 | 2,925 | 1,740 | 1,675 |
| ≥ 1 kb | 2,327 | 143 | 1,882 | 1,803 | 1,063 | 1,040 |
| ≥ 5 kb | 269 | 143 | 201 | 177 | 62 | 61 |
| ≥ 10 kb | 139 | 143 | 67 | 51 | 22 | 21 |
| ≥ 50 kb | 66 | 143 | 8 | 3 | 6 | 6 |
| ≥ 100 kb | 49 | 143 | 2 | 2 | 0 | 0 |
| ≥ 500 kb | 0 | 143 | 0 | 0 | 0 | 0 |
| ≥ 1 Mb | 0 | 95 | 0 | 0 | – | – |
| ≥ 5 Mb | 0 | 0 | 0 | 0 | – | – |

Tuned PAV calls fewer variants than default PAV at every cutoff below 1 kb and from 1 to 50 kb inside the active HOR (Supplementary Note 1). Both PAV runs report the same two ≥ 100 kb HOR calls. Genome-wide and outside the HOR, PAV reports more calls than SVIM-asm at ≥ 500 kb: 33 default PAV, 31 tuned PAV, and 0 SVIM-asm, which reclassifies these as breakends. dipcall reports the fewest calls at every cutoff up to 10 kb, and 0 inside the HOR arrays of chr1, chr5, chr6, chr13, chrX, and chrY. Counts on chrX and chrY under default diploid vcfpair are lower bounds. the dipcall kit bundles minimap2 2.20-r1061 against 2.30 in our pipeline.

**Table S2:**
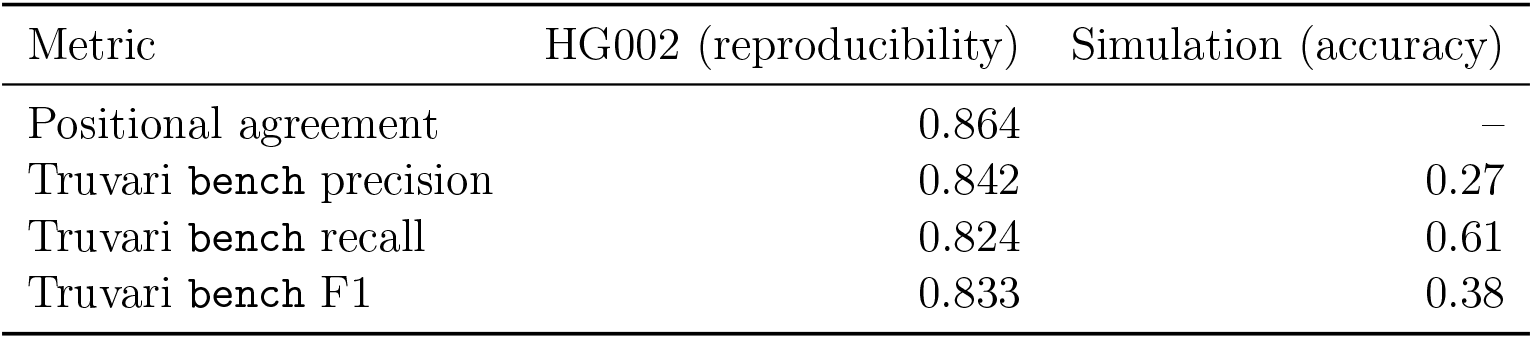
SVIM-asm reproducibility between two HG002 assemblies and accuracy in simulation. (Supplementary Notes 1 and 13). HG002: calls from the HPRC Release 2 assemblies compared with calls from the same pipeline on the Q100 HG002 v1.1 assemblies. Simulation: calls compared with the 891 simulated SVs within Truvari’s default size range (50 bp–50 kb) in 69 simulated chromosomes. Truvari bench settings are identical in both columns (–refdist 500 –pctseq 0.7 –pctsize 0.7 –pick single).

**Table S3:** Assembly contiguity limits centromeric SV detection. (Supplementary Note 2). Perhaplotype SV yield at chromosome 9, HiFi HPRC R2 against ONT-R9.4.1 Shasta.

| Metric | HiFi (HPRC R2) | ONT-R9.4.1 (Shasta) |
|---|---|---|
| Contigs per haplotype | 74–120 | 2,503–3,300 |
| Contig N50 | ∼134–149 Mb | ∼26–35 Mb |
| Median chr9 SV calls | ∼150 | ∼25 |
| *Depletion in ONT, by class* |  |  |
| DUP:TANDEM |  | 13.9× fewer |
| Breakends |  | 12.4× fewer |
| All classes |  | 3.8× fewer |

**Table S4:** Haplogroup resolution by chromosome. (one row per centromere). For each resolved SV, at least one branch of the chromosome’s haplotype tree explains its carrier set; *components* counts the resulting monophyletic components. *Later origins* are the components found after the first branch match was masked, and *imputed* the number of those with a fitted elastic-net model. Multi-origin share is the fraction of that chromosome’s resolved SVs with more than one origin (Supplementary Fig. S1e).

| Chromosome | Resolved SVs | > 1 origin | Components | Later origins | Imputed | > 1 origin (%) |
|---|---|---|---|---|---|---|
| 1 | 388 | 29 | 417 | 29 | 5 | 7.5 |
| 2 | 55 | 2 | 57 | 2 | 1 | 3.6 |
| 3 | 18 | 0 | 18 | 0 | 0 | 0.0 |
| 4 | 32 | 0 | 32 | 0 | 0 | 0.0 |
| 5 | 45 | 4 | 49 | 4 | 0 | 8.9 |
| 6 | 143 | 12 | 155 | 12 | 2 | 8.4 |
| 7 | 28 | 3 | 31 | 3 | 2 | 10.7 |
| 8 | 41 | 5 | 46 | 5 | 2 | 12.2 |
| 9 | 114 | 8 | 123 | 9 | 0 | 7.0 |
| 10 | 85 | 5 | 91 | 6 | 3 | 5.9 |
| 11 | 95 | 4 | 99 | 4 | 0 | 4.2 |
| 12 | 103 | 6 | 109 | 6 | 6 | 5.8 |
| 13 | 34 | 3 | 37 | 3 | 0 | 8.8 |
| 14 | 51 | 4 | 55 | 4 | 1 | 7.8 |
| 15 | 71 | 1 | 72 | 1 | 0 | 1.4 |
| 16 | 26 | 3 | 30 | 4 | 0 | 11.5 |
| 17 | 152 | 5 | 157 | 5 | 1 | 3.3 |
| 18 | 38 | 2 | 40 | 2 | 0 | 5.3 |
| 19 | 434 | 33 | 473 | 39 | 7 | 7.6 |
| 20 | 27 | 0 | 27 | 0 | 0 | 0.0 |
| 21 | 42 | 1 | 43 | 1 | 0 | 2.4 |
| 22 | 69 | 5 | 74 | 5 | 2 | 7.2 |
| X | 12 | 0 | 12 | 0 | 0 | 0.0 |
| All | 2,103 | 135 | 2,247 | 144 | 32 | 6.4 |

**Table S5:** Determinants of imputability. (Supplementary Note 3). Layer 1 classifies whether an SV is imputable; Layer 2 predicts imputation accuracy.

| Quantity | Value |
|---|---|
| Catalog SVs classified (Layer 1) | 192,363 |
| Imputable fraction | 5.6% |
| Layer 1 separability (5-fold ROC-AUC) | 0.989 |
| Layer 2 held-out *R*<sup>2</sup>, sequence/chromosome features | 0.407 |
| Layer 2 held-out *R*<sup>2</sup>, adding mapping-quality features | 0.457 |
| Top permutation importance (fraction secondary alignments) | 0.133 |
| Grouped drop-column loss, mapping-quality family | Δ*R*<sup>2</sup> = −0.054 |
| Q100 reproduction, imputable SVs | 0.904 |
| Q100 reproduction, unimputable SVs | 0.848 |
| Difference (chromosome-block bootstrap 95% CI) | +0.055 [−0.011, +0.124] |

**Table S6:** Mapping-quality features are the strongest predictors of imputation accuracy. (Supplementary Note 3). Permutation importance (mean s.d.) for the Layer 2 model, built without the haplogroup-category feature and with contig-level alignment features. marks the mapping-quality features; the top sequence-composition feature (GC over 10 kb) ranks fifth.

| Rank | Feature | Permutation importance |
|---|---|---|
| 1 | Fraction of secondary alignments at locus<sup>†</sup> | 0.133 ± 0.009 |
| 2 | Homozygous-alternate count | 0.130 ± 0.009 |
| 3 | Contig count at locus<sup>†</sup> | 0.091 ± 0.007 |
| 4 | log active HOR span | 0.080 ± 0.006 |
| 5 | GC content, 10 kb | 0.065 ± 0.006 |
| 6 | Mean MAPQ at locus<sup>†</sup> | 0.052 ± 0.005 |
| 7 | Mean edit distance at locus<sup>†</sup> | 0.050 ± 0.004 |
| 8 | GC content, 1 Mb | 0.049 ± 0.005 |
| 9 | log SV length | 0.036 ± 0.004 |
| 10 | GC content, 100 kb | 0.034 ± 0.004 |

**Table S7:** Elastic net matches a Beagle 5 copying model genome-wide. (Supplementary Note 4). Assembly-co-phased benchmark panel; genome-wide centromere-level block bootstrap.

| Comparison | Value |
|---|---|
| Positive control (held-out common SNPs, Beagle *r*<sup>2</sup>) | 0.93–0.97 |
| Effective population size *n*<sub>*e*</sub> | 2,000 |
| All 23 centromeres, mean *r*<sup>2</sup> gap | −0.003 |
| 95% CI | [−0.013, +0.008] |
| *P* | 0.59 |
| monophyletic class, *P* | 0.96 |
| Centromeres where elastic net leads, MAC ≥ 20 | 12 / 23 |
| monophyletic class | 13 / 23 |
| chr17 alone, elastic net/Beagle ratio (Pearson *r*<sup>2</sup>) | ∼1.3× |

**Table S8:** Concordance with k-mer centromere-haplogroup QTL. (Supplementary Note 5). We typed haplogroups from short-read k-mers with ascairn.

| Quantity | Value |
|---|---|
| MAGE individuals typed | 728 |
| Haplogroup variants tested | 107 / 209 |
| cis-eGenes | 66 |
| cis-sGenes | 49 |
| Distinct cis genes | 101 |
| Genomic inflation λ | 1.016 (expression), 1.062 (splicing) |
| Cis genes among the 141 imputed-SV cis genes | 73 / 101 |

**Table S9:** Dip position along the *α*-satellite array, by chromosome. (Supplementary Note 7). 21,515 dip calls within active arrays on single contigs, chromosomes 1–22 and X: 4,795 haplotypes across 188 samples. CDR annotations merge dips within 150 kb. Dip position *x* runs from 1 (p-side end) through 0 (center) to +1 (q-side end). Each haplotype contributes the median *x* over its dips; median offset is the median *x* across haplotypes. A two-sided sign test against an even split identifies a preferred side at *P <* 0.05*/*23.

| Chromosome | Morphology | CDR dips<br>/ annot. | Haplotypes | Median<br>position | p side | Median<br>\|offset\| | Sign-test<br>*P* | Lean |
|---|---|---|---|---|---|---|---|---|
| chr16 | metacentric | 1,391 / 281 | 269 | −0.669 | 99.3% | 0.669 | 7.7 × 10<sup>−77</sup> | p |
| chr13 | acrocentric | 1,156 / 265 | 262 | −0.760 | 98.5% | 0.761 | 5.3 × 10<sup>−71</sup> | p |
| chr15 | acrocentric | 874 / 225 | 218 | −0.632 | 94.0% | 0.632 | 1.4 × 10<sup>−45</sup> | p |
| chr14 | acrocentric | 1,163 / 299 | 288 | −0.581 | 91.0% | 0.587 | 3.0 × 10<sup>−50</sup> | p |
| chr7 | submetacentric | 650 / 154 | 140 | −0.714 | 89.3% | 0.733 | 8.9 × 10<sup>−23</sup> | p |
| chr21 | acrocentric | 1,191 / 292 | 284 | −0.590 | 88.7% | 0.594 | 1.5 × 10<sup>−43</sup> | p |
| chr1 | metacentric | 1,344 / 279 | 245 | −0.427 | 84.9% | 0.448 | 4.5 × 10<sup>−30</sup> | p |
| chr9 | submetacentric | 1,039 / 294 | 278 | −0.473 | 80.9% | 0.527 | 2.1 × 10<sup>−26</sup> | p |
| chrX | submetacentric | 890 / 212 | 206 | −0.280 | 78.2% | 0.344 | 1.7 × 10<sup>−16</sup> | p |
| chr22 | acrocentric | 1,000 / 280 | 270 | −0.526 | 77.8% | 0.610 | 1.1 × 10<sup>−20</sup> | p |
| chr12 | submetacentric | 1,161 / 251 | 232 | −0.480 | 65.5% | 0.669 | 2.6 × 10<sup>−6</sup> | p |
| chr19 | metacentric | 837 / 169 | 158 | −0.319 | 60.1% | 0.599 | 0.013 | — |
| chr5 | submetacentric | 1,059 / 226 | 192 | −0.124 | 56.2% | 0.405 | 0.097 | — |
| chr4 | submetacentric | 584 / 138 | 133 | 0.007 | 49.6% | 0.330 | 1.00 | — |
| chr17 | submetacentric | 437 / 94 | 88 | 0.051 | 47.7% | 0.357 | 0.75 | — |
| chr10 | submetacentric | 1,055 / 218 | 211 | 0.204 | 37.4% | 0.532 | 3.2 × 10<sup>−4</sup> | q |
| chr11 | submetacentric | 1,058 / 288 | 240 | 0.298 | 28.3% | 0.394 | 1.4 × 10<sup>−11</sup> | q |
| chr20 | metacentric | 905 / 246 | 210 | 0.498 | 27.1% | 0.573 | 2.5 × 10<sup>−11</sup> | q |
| chr3 | metacentric | 505 / 113 | 111 | 0.244 | 27.0% | 0.321 | 1.4 × 10<sup>−6</sup> | q |
| chr18 | submetacentric | 247 / 72 | 60 | 0.236 | 26.7% | 0.300 | 3.9 × 10<sup>−4</sup> | q |
| chr8 | submetacentric | 1,048 / 240 | 235 | 0.413 | 22.1% | 0.469 | 2.7 × 10<sup>−18</sup> | q |
| chr6 | submetacentric | 551 / 144 | 139 | 0.848 | 17.3% | 0.863 | 1.9 × 10<sup>−15</sup> | q |
| chr2 | submetacentric | 1,370 / 361 | 326 | 0.359 | 11.0% | 0.369 | 1.8 × 10<sup>−50</sup> | q |

**Table S10:** Five chromosomes have more than one common CDR location. (Supplementary Note 7). On five of the 24 chromosomes with an active array the CDR locations form two separated major clusters; on the remaining 19 they form one continuous major cluster. Position is given on the relative axis *x* of Supplementary Table S9 rescaled to (*x* + 1)*/*2, so that 0 is the p-side end of the active array and 1 the q-side end. “Apart” converts the distance between the two positions to megabases at that chromosome’s median array length. “In the gap” is the share of CDR annotations whose position falls in the middle 30 % of the interval between the two, with the share expected under a uniform distribution over the same array in parentheses. More than one preferred position is supported on all five by excess mass calibrated against a Beta distribution fitted to that chromosome and resampled over haplotypes (*P* 0.004), and both positions are recovered independently in each half of the cohort in 99–100 % of 300 random divisions of the 188 individuals.

| Chromosome | CDR annotations | Lower position | Upper position | Apart (Mb) | Annotations at each | In the gap |
|---|---|---|---|---|---|---|
| chrX | 212 | 0.07 | 0.47 | 1.3 | 90 / 122 | 3 % (12 % expected) |
| chr19 | 169 | 0.19 | 0.87 | 2.5 | 102 / 67 | 7 % (20 % expected) |
| chr10 | 218 | 0.17 | 0.74 | 1.2 | 74 / 144 | 8 % (17 % expected) |
| chr12 | 251 | 0.12 | 0.86 | 2.0 | 166 / 85 | 10 % (22 % expected) |
| chr6 | 144 | 0.09 | 0.94 | 2.1 | 26 / 118 | 1 % (26 % expected) |

**Table S11:** Chromosome-8 leads for dip position and their best tag SNP. Each row is one haplogroup resolution of a physical SV, with its own carrier set, in CHM13v2.0 coordinates; positions repeat across resolutions. *P* alone fits the structural variant on its own; *P* joint adds its best tag SNP; *r* tag is the linkage between the two. Bold marks the lead, tagged at *r* = 0.95. The 1,868-bp insertion, tagged at 0.40, retains its association in the joint fit (*P* = 0.019).

| Position | Type | Length | AF | CV *R*<sup>2</sup> | \|*r*\| tag | *P* alone | *P* joint |
|---|---|---|---|---|---|---|---|
| 44,300,221 | DEL | 113 | 0.533 | 0.93 | **0.95** | 7.0e-09 | 0.920 |
| 44,302,147 | INS | 59 | 0.639 | 0.65 | 0.79 | 1.9e-08 | 0.109 |
| 44,302,147 | INS | 59 | 0.675 | 0.65 | 0.75 | 2.7e-08 | 0.074 |
| 44,460,941 | DEL | 347 | 0.238 | 0.46 | 0.66 | 1.1e-06 | 0.116 |
| 44,247,324 | INS | 1,868 | 0.119 | 0.38 | 0.40 | 1.8e-06 | 0.019 |
| 44,300,221 | DEL | 113 | 0.619 | 0.93 | 0.78 | 2.2e-06 | 0.920 |
| 44,460,941 | DEL | 347 | 0.305 | 0.46 | 0.55 | 4.5e-04 | 0.441 |
| 44,300,221 | DEL | 113 | 0.086 | 0.93 | 0.26 | 1.7e-01 | 0.951 |

**Table S12:** Genotype composition and Hardy–Weinberg fit of every discovery variant. One row per dosage profile, ordered by Hardy–Weinberg *p*; genotypes are observed calls only, with mean-imputed dosages excluded. *n* counts individuals with an observed call, 0/1/2 their genotype counts, carriers the heterozygotes and alternate homozygotes together. The last three columns count the discoveries each variant contributes, split at 5 Mb on the same chromosome and on any other chromosome; they sum to the 360, 58 and 42 pairs of the three refit approaches. ^a^chr3:91,750,609 and chr3:91,750,618, 336-bp deletions 9 bp apart, have an identical dosage vector and were tested once; they share this row, so the 45 rows cover 46 calls.

| Variant | Type | Length | *n* | 0/1/2 | Carr. | HWE *P* | ≤5 Mb | >5 Mb | Other |
|---|---|---|---|---|---|---|---|---|---|
| chr17:26,788,109 | INS | 167 | 109 | 100/6/3 | 9 | 8.8 × 10<sup>−7</sup> | 1 | 9 | 14 |
| chr18:20,747,890 | INS | 2,881 | 103 | 88/12/3 | 15 | 6.2 × 10<sup>−3</sup> | 0 | 0 | 1 |
| chr7:60,420,588 | INS | 1,360 | 124 | 54/65/5 | 70 | 7.0 × 10<sup>−3</sup> | 5 | 0 | 0 |
| chr6:61,053,641 | DEL | 54 | 122 | 93/23/6 | 29 | 0.010 | 24 | 0 | 0 |
| chr17:26,799,131 | DEL | 168 | 101 | 39/56/6 | 62 | 0.015 | 22 | 1 | 0 |
| chr17:27,481,433 | INS | 506 | 121 | 78/43/0 | 43 | 0.017 | 1 | 0 | 0 |
| chr17:27,431,973 | DEL | 167 | 105 | 58/34/13 | 47 | 0.034 | 26 | 0 | 0 |
| chr22:12,790,199 | DEL | 1,364 | 117 | 103/12/2 | 14 | 0.035 | 1 | 1 | 0 |
| chr10:39,636,872 | DEL | 337 | 124 | 78/45/1 | 46 | 0.043 | 17 | 0 | 0 |
| chr19:29,757,702 | INS | 328 | 124 | 88/36/0 | 36 | 0.059 | 3 | 0 | 0 |
| chr16:35,891,049 | DEL | 2,036 | 118 | 102/14/2 | 16 | 0.086 | 6 | 1 | 0 |
| chr7:60,423,386 | DEL | 340 | 121 | 80/33/8 | 41 | 0.087 | 46 | 0 | 0 |
| chr18:15,994,275 | DEL | 342 | 122 | 75/45/2 | 47 | 0.100 | 1 | 0 | 0 |
| chr5:49,591,352 | INS | 679 | 112 | 82/30/0 | 30 | 0.102 | 7 | 0 | 0 |
| chr18:15,967,546 | DEL | 1,695 | 124 | 107/15/2 | 17 | 0.106 | 0 | 0 | 2 |
| chr3:91,779,442 | DEL | 339 | 123 | 92/31/0 | 31 | 0.110 | 7 | 0 | 0 |
| chr14:10,098,447 | DEL | 1,363 | 124 | 84/39/1 | 40 | 0.120 | 1 | 0 | 0 |
| chr3:91,778,320 | DEL | 1,011 | 123 | 93/30/0 | 30 | 0.123 | 0 | 1 | 0 |
| chr3:91,750,609<sup>a</sup> | DEL | 336 | 124 | 85/38/1 | 39 | 0.140 | 16 | 0 | 0 |
| chr10:39,640,346 | DEL | 338 | 124 | 100/21/3 | 24 | 0.157 | 100 | 2 | 0 |
| chr16:35,850,459 | DEL | 2,032 | 124 | 67/44/13 | 57 | 0.167 | 4 | 0 | 0 |
| chr2:94,630,228 | INS | 679 | 123 | 98/25/0 | 25 | 0.210 | 0 | 1 | 0 |
| chr8:46,321,659 | INS | 338 | 124 | 104/18/2 | 20 | 0.255 | 6 | 2 | 0 |
| chr22:12,803,133 | DEL | 171 | 113 | 47/48/18 | 66 | 0.336 | 1 | 0 | 0 |
| chr18:15,979,817 | DEL | 675 | 124 | 92/31/1 | 32 | 0.352 | 2 | 0 | 0 |
| chr16:35,904,295 | DEL | 339 | 113 | 100/12/1 | 13 | 0.359 | 0 | 4 | 4 |
| chr14:10,118,311 | DEL | 1,364 | 108 | 91/17/0 | 17 | 0.375 | 0 | 0 | 2 |
| chr16:35,864,722 | DEL | 336 | 124 | 102/20/2 | 22 | 0.389 | 0 | 1 | 0 |
| chr22:12,807,563 | INS | 1,192 | 108 | 87/19/2 | 21 | 0.433 | 1 | 1 | 0 |
| chr2:94,638,534 | INS | 1,022 | 123 | 108/15/0 | 15 | 0.471 | 0 | 7 | 2 |
| chr16:35,890,476 | DEL | 3,053 | 119 | 105/14/0 | 14 | 0.495 | 1 | 5 | 7 |
| chr14:10,096,863 | INS | 342 | 124 | 81/37/6 | 43 | 0.511 | 2 | 0 | 0 |
| Variant | Type | Length | *n* | 0/1/2 | Carr. | HWE *P* | ≤5 Mb | >5 Mb | Other |
| chr14:10,094,126 | INS | 1,364 | 124 | 111/13/0 | 13 | 0.538 | 4 | 1 | 3 |
| chr17:27,445,504 | INS | 679 | 114 | 102/12/0 | 12 | 0.553 | 0 | 6 | 1 |
| chr15:16,686,871 | INS | 1,870 | 124 | 112/12/0 | 12 | 0.571 | 0 | 8 | 3 |
| chr18:15,982,437 | INS | 342 | 124 | 114/10/0 | 10 | 0.640 | 2 | 2 | 1 |
| chr3:91,748,471 | INS | 2,888 | 124 | 98/24/2 | 26 | 0.707 | 0 | 0 | 2 |
| chr3:91,771,109 | DEL | 339 | 123 | 91/30/2 | 32 | 0.791 | 4 | 1 | 0 |
| chr3:91,742,653 | INS | 1,018 | 124 | 100/23/1 | 24 | 0.797 | 19 | 0 | 0 |
| chr3:91,756,698 | DEL | 337 | 124 | 67/49/8 | 57 | 0.810 | 10 | 0 | 0 |
| chr3:91,775,969 | INS | 3,563 | 123 | 100/22/1 | 23 | 0.861 | 0 | 1 | 0 |
| chr2:94,662,333 | INS | 680 | 124 | 101/22/1 | 23 | 0.868 | 0 | 2 | 0 |
| chr8:46,321,868 | INS | 338 | 124 | 96/26/2 | 28 | 0.876 | 13 | 0 | 0 |
| chr3:91,762,776 | DEL | 339 | 124 | 60/52/12 | 64 | 0.881 | 7 | 0 | 0 |
| chr9:44,983,369 | INS | 854 | 103 | 71/29/3 | 32 | 0.985 | 0 | 1 | 0 |

**Table S13:**
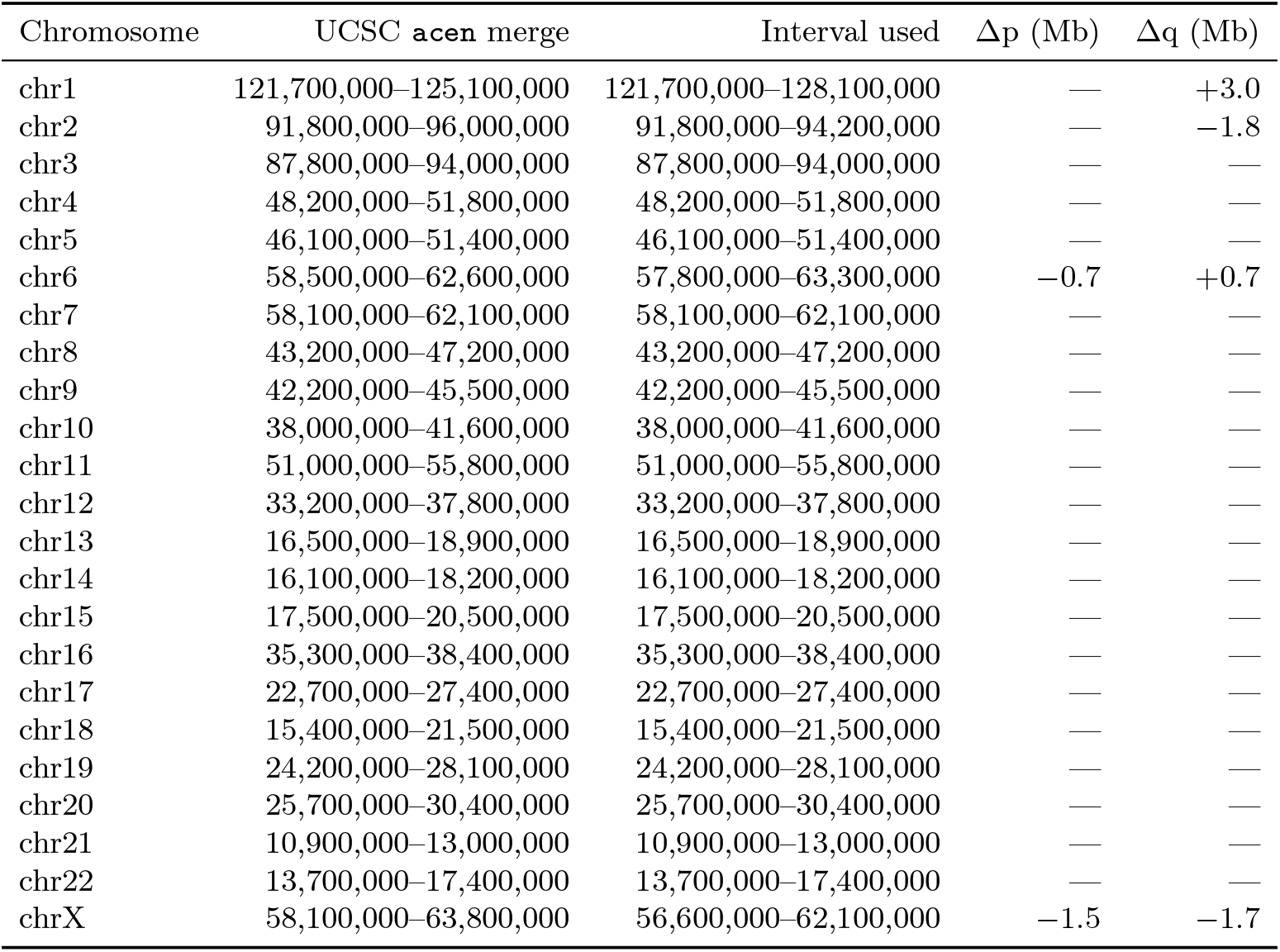
Centromere intervals used in Strategy 1. Per-chromosome merges of the p and q acen bands from UCSC hg38 cytoBand, against the intervals applied.

**Table S14:** Datasets, reference resources, software and deposited products used in this study. Structural variants, satellite annotations and centromere dip regions are on T2T-CHM13 v2.0; predictor SNPs, centromere boundary tracks, cross-mappability tables, methylation pileups, GWAS summary statistics and biobank genotypes are on GRCh38.

| Resource | Version or release | Role in this study | Accession, DOI or URL |
|---|---|---|---|
| **A. Primary datasets** |  |  |  |
| HPRC Release 2 haplotype-resolved assemblies | freeze 2; v1.0, v1.0.1 and per-sample ncbi_upload builds | 196 individuals; assembly-to-CHM13 alignment and SV discovery | s3://human-pangenomics/submissions/DC27718F-5F38-43B0-9A78-270F395F13E8--INT_ASM_PRODUCTION/<sample>/assemblies/freeze_2/; https://humanpangenome.org/hprc-data-release-2/ |
| HPRC R2 per-haplotype CenSat, as_hor and active-centromere BED | Release 2, v1 | active-array definition per haplotype | s3://human-pangenomics/submissions/DC27718F-5F38-43B0-9A78-270F395F13E8--INT_ASM_PRODUCTION/<sample>/assemblies/freeze_2/annotation/censat/ |
| HPRC R2 haplotype-resolved 5mC bedMethyl | Release 2 freeze 2 | 384 haplotypes; CDR calling, local methylation windows | s3://human-pangenomics/submissions/DC27718F-5F38-43B0-9A78-270F395F13E8--INT_ASM_PRODUCTION/<sample>/assemblies/freeze_2/annotation/methylation/ont/; https://registry.opendata.aws/hprc-epigenome/ |
| HPRC R2 Minigraph–Cactus chains | Release 2, vs CHM13 | second alignment route for local methylation windows | s3://human-pangenomics/submissions/DC27718F-5F38-43B0-9A78-270F395F13E8--INT_ASM_PRODUCTION/<sample>/assemblies/freeze_2/annotation/chains/minigraph-cactus/ |
| HPRC R2 genome-wide PAV call set | Release 2 | community comparator for the caller choice | Lucas et al. [2026] |
| HPRC sample and population metadata | year-one (46) and Release 2 (234) | ancestry labels; platform and basecaller covariates | https://github.com/human-pangenomics |
| T2T-CHM13 v2.0 analysis set | v2.0 | alignment target and coordinate system for all SV calls | s3://human-pangenomics/T2T/CHM13/assemblies/analysis_set/chm13v2.0.fa.gz |
| CenSat satellite annotation | v2.1 | active alpha-satellite HOR arrays; H1L records define the 62.25 Mb restriction | s3://human-pangenomics/T2T/CHM13/assemblies/annotation/chm13v2.0_censat_v2.1.bed |
| Q100 HG002 assembly | v1.1 | SV-call reproducibility benchmark | s3://human-pangenomics/T2T/HG002/assemblies/hg002v1.1.fasta.gz |
| Global centromere sequences and annotations | figshare v1 | AS-HOR_stv_row.ort active-array intervals; 150 kb merge convention | https://doi.org/10.6084/m9.figshare.29921990 |
| 1000 Genomes 30x NYGC high-coverage phased panel | 20220422 3202-sample release | predictor SNPs, imputation targets, EUR-525 LD reference | http://ftp.1000genomes.ebi.ac.uk/vol1/ftp/data_collections/1000G_2504_high_coverage/working/20220422_3202_phased_SNV_INDEL_SV |
| 1000 Genomes pedigree and population file | 20130606, 3202 samples | founder status, family, sex, superpopulation; LD-panel and target selection | http://ftp.1000genomes.ebi.ac.uk/vol1/ftp/data_collections/1000G_2504_high_coverage/20130606_g1k_3202_samples_ped_population.txt |
| 1000 Genomes 30x GRCh38 CRAMs | 3202-sample collection | short reads for ascairn k-mer haplogroup typing | https://www.internationalgenome.org/data-portal |
| 1000 Genomes ONT haplotype-resolved 5mC bedMethyl | modkit v0.1.11 pileups; ONT R9.4.1 | 500 LCLs; methylation phenotype (176 in cohort, 124 with long-read genotypes) | s3://1000g-ont/PROCESSED_DATA/ALIGNED_TO_HG38/MODKIT/; https://s3.amazonaws.com/1000g-ont/index.html |
| 1000 Genomes ONT CHM13-aligned read alignments | same release | input alignments for Sniffles2 re-genotyping at catalog sites | https://s3.amazonaws.com/1000g-ont/index.html |
| 1000 Genomes ONT CHM13-aligned variant calls | same release | first conditional SNP panel of the methylation-QTL analysis | https://s3.amazonaws.com/1000g-ont/index.html |
| 1000 Genomes ONT R9.4.1 assemblies (Shasta) | same release | assembly-contiguity comparator, Supplementary Note 2 | https://s3.amazonaws.com/1000g-ont/index.html |
| T2T-CHM13 1000 Genomes joint callset | SHAPEIT5-phased v1.1 | second conditional SNP panel of the methylation-QTL analysis; SNP backbone of the Beagle benchmark | s3://human-pangenomics/T2T/CHM13/assemblies/variants/1000_Genomes_Project/chm13v2.0/Phased_SHAPEIT5_v1.1/ |
| MAGE expression and splicing phenotypes | v1.0 | 630 LCLs; eGene and sGene phenotypes and their covariates | https://zenodo.org/records/10535719; https://github.com/mccoy-lab/MAGE |
| MAGE SNP-based cis-QTL summary statistics | v1.0 | comparator defining a novel gene | Taylor et al. [2024] |
| **B. Annotation and reference resources** |  |  |  |
| UCSC hg38 cytoBand | hg38 | acen bands merged per chromosome; centromere boundaries for Strategy 1 | https://hgdownload.soe.ucsc.edu/goldenPath/hg38/database/cytoBand.txt.gz |
| UCSC hg38 gap track | hg38 | heterochromatin and short_arm records for the heterochromatin-aware flanking | https://hgdownload.soe.ucsc.edu/goldenPath/hg38/database/gap.txt.gz |
| Saha and Battle cross-mappability tables | GRCh38 / GENCODE v26 | trans-QTL artefact screen | https://doi.org/10.6084/m9.figshare.7315136.v3 |
| GENCODE annotation | v26 | gene models underlying the cross-mappability tables | https://doi.org/10.6084/m9.figshare.7315136.v3 |
| ascairn reference panel ascairn_paper_2025 | 451 MB; HPRC, HGSVC, T2T | assembled active-HOR haplotypes for k-mer typing | https://github.com/friend1ws/ascairn |
| ascairn active-HOR FASTA release | Zenodo 19601002 | aHOR sequences of the reference panel | https://zenodo.org/records/19601002 |
| ssimp build database | 2018-01-18 build | summary-statistic imputation reference | https://doi.org/10.5281/zenodo.13222092 |
| LDetect / UK Biobank b38 LD blocks and LD reference | cTWAS resources | super-region partition and admission filter | https://uchicago.box.com/s/jqocacd2fulskmhoqnasrknbt59x3xkn |
| T2T-CHM13 recombination map (Lalli et al. 2025) | averagedByBase, scaled to deCODE, no mask | genetic map for the Beagle imputation benchmark, Supplementary Note 4; the published map masks the centromere core, so a flat rate was spliced inside the alpha-satellite boundary | https://zenodo.org/records/14891074 |
| **C. Association and biobank resources** |  |  |  |
| FinnGen summary statistics | R12 | 159 disease endpoints of the 464-trait joint analysis | https://www.finngen.fi/en/access_results; gs://finngen-public-data-r12/summary_stats/release/ |
| Neale lab UK Biobank GWAS | round 2 | 305 quantitative and biomarker phenotypes | http://www.nealelab.is/uk-biobank |
| All of Us Research Program | Controlled Tier, curated data release v8 | biobank replication; genotypes, EHR phenotypes, ancestry, relatedness | https://www.researchallofus.org/data-tools/workbench/ |
| phecodeX phenotype definitions | – | 2,530 definitions; 2,417 clear the case and control floor | Shuey et al. [2023] |
| UK Biobank genotyping-array marker set | controlled access | pericentromeric SNP-density comparison | UK Biobank access application |
| **D. Products deposited by this study** |  |  |  |
| cenSVhap | this study | centromere haplogroup-resolution method | https://github.com/YikaiDong-git/cenSVhap |
| cenSVimp | this study | SNP panel construction, elastic-net training, imputation | https://github.com/YikaiDong-git/cenSVimp |
| cenSV-analysis | this study | workflows built on published tools; Workbench drivers; figure scripts | https://github.com/YikaiDong-git/cenSV-analysis |
| CenSV browser | this study | interactive view of the haplogroup-resolved SVs and their associations | https://yikaidong-git.github.io/CenSV |
| **E. Software** |  |  |  |
| minimap2 | 2.30 | [Li, 2018] |  |
| SVIM-asm | 1.0.3 | [Heller and Vingron, 2021]; https://github.com/eldariont/svim-asm |  |
| PAV | 2.4.6 (88df4dbf) | [Ebert et al., 2021]; https://github.com/EichlerLab/pav |  |
| dipcall | 0.3 | [Li et al., 2018a] |  |
| Truvari | 5.4.0 | [English et al., 2022] |  |
| Sniffles2 | 2.7.1 | [Smolka et al., 2024] |  |
| SAMtools / BCFtools / HTSlib | 1.22.1 / 1.22 / 1.22.1 | [Danecek et al., 2021] |  |
| tabix | 1.11 | – |  |
| BEDTools | 2.31.1 | – |  |
| mosdepth | 0.3.14 | – |  |
| Snakemake-HumAS-SD (StV / HumAS-HMMER) | – | [Altemose et al., 2022]; https://github.com/logsdon-lab/Snakemake-HumAS-SD |  |
| HORmon | 1.0.0 | [Kunyavskaya et al., 2022]; https://github.com/ablab/HORmon |  |
| StringDecomposer | 1.1.2 | [Dvorkina et al., 2020]; https://github.com/ablab/stringdecomposer |  |
| ascairn | 0.3.0 | [Shiraishi et al., 2026]; https://github.com/friend1ws/ascairn |  |
| CDR-Finder | commit 42bf9c9 | [Mastrorosa et al., 2024]; https://github.com/koisland/CDR-Finder |  |
| modkit | 0.6.0 (0.1.11 upstream) | https://github.com/nanoporetech/modkit |  |
| Beagle | 5.5 | [Browning et al., 2018] |  |
| PLINK 2 | 2.0.0a.6.9 | [Chang et al., 2015] |  |
| tensorQTL | 1.0.10 | [Taylor-Weiner et al., 2019] |  |
| susieR | 0.14.2 | [Wang et al., 2020] |  |
| ssimp | 0.5.5 | [Rüeger et al., 2018] |  |
| scikit-learn | 1.8.0 | [Pedregosa et al., 2011] |  |
| scikit-bio | 0.7.2 | [Saitou and Nei, 1987] |  |
| qvalue | 2.42.0 | [Storey and Tibshirani, 2003] |  |
| process_neale_lab_GWAS | commit 83578d1 | https://github.com/weinstockj/process_neale_lab_GWAS |  |
| Snakemake | 9.13.7 / 8.30.0 | – |  |
| Python / R | 3.11.14, 3.12.13 / 4.5.2 | – |  |

### Supplementary Note 10 Summary-level joint fine-mapping of imputed SVs (cTWAS / SuSiE-RSS)

#### S10.1 Imputed SVs share credible sets with flanking SNPs in most joint fine-mapping fits

Jointly modeling imputed centromeric SVs and nearby SNPs tests how posterior support is distributed among these correlated predictors. We fitted each centromeric super-region’s included SVs alongside their supporting SNPs in a single SuSiE-RSS model [Zou et al., 2022], adapting the cTWAS framework [Zhao et al., 2024] to matched GWAS from UK Biobank (Neale) and FinnGen (Methods). For each matched trait, high posterior inclusion probabilities (PIP) for individual SVs were cohort-specific.

After re-fitting against a panel of *∼*12,000 SNPs from the same region (Methods), most joint candidate associations returned a credible set that spanned collinear variants. Of 473 single-SV candidate hits and 3,688 joint candidate hits, 36 (10 SVs) and 17 (3 SVs), respectively, retained PIP *>* 0.5. Among the remaining 3,671 joint SV–trait pairs (all 3,688 pairs but the 17 that retained PIP *>* 0.5), 63.6% returned unresolved credible sets in which the collinear pericentromeric LD left multiple credible variants (SV or SNP). A further 22.2% failed to converge, and in 14.2% a specific SNP received higher posterior support than the SV (Supplementary Fig. S5). The SV lay inside a credible set in 90.4% of unresolved SV–trait pairs against 67.7% of pairs in which a SNP received higher posterior support (robust to a sensitivity re-fit and a lower inclusion threshold; Supplementary Note 11). Most joint candidate associations therefore implicate a set of linked pericentromeric variants without distinguishing the SV from nearby SNPs.

Two SVs passed all three analyses, one on a single trait and one on multiple traits. The first, a chromosome-11 deletion (CV *R*^2^ = 0.41), passed on one trait. The second, an insertion in the chromosome-19 pericentromere, passed on 26, 15 and 12 traits in the three analyses at CV *R*^2^ = 0.999, with a median dense-panel PIP of 0.999 (range 0.79–1.00; *≥* 0.98 in 21 of its 26 single-SV traits). The SNPs used to impute it span a 4.5-Mb haplotype (GRCh38 chr19:23.9– 28.4 Mb) straddling the centromere and tags at least two independent association peaks *∼*3 Mb apart, and at CV *R*^2^ *≈* 1 the SV itself lies within the span of those predictor SNPs. The insertion therefore marks a recombination-suppressed block across the centromere. The trait association is localized to the haplotype block marked by the insertion. Distinguishing the insertion from other variants in this block requires direct genotyping across the region.

**Table S15:** Datasets, versions and accessions. Builds are GRCh38 throughout.

| Resource | Role | Source / accession |
|---|---|---|
| 1000 Genomes 30× (NYGC) high-coverage, phased | LD reference panel (525 EUR founders) and ancestry-matched LD | internationalgenome.org; Byrska-Bishop et al. [2022] |
| FinnGen | 159 disease/endpoint GWAS (binary) | finngen.fi; Kurki et al. [2023] |
| Neale lab UK Biobank GWAS (round 2) | 305 quantitative/biomarker GWAS | nealelab.is/uk-biobank |
| Xin He lab LDetect / UKBB b38 LD blocks | region (block) partition and inclusion-filter LD info | Berisa and Pickrell [2016]; cTWAS resources [Zhao et al., 2024] |
| ssimp build database | summary-statistic imputation reference (ssimp 0.5.5) | Zenodo 13222092; Rüeger et al. [2018] |

#### S10.2 Overview and design

We tested whether pericentromeric structural variants (SVs) are associated with human complex traits by integrating SV dosages, imputed from local single-nucleotide polymorphisms (SNPs), with publicly available GWAS summary statistics in a causal TWAS (cTWAS) framework [Zhao et al., 2024]. SV dosages are not present in GWAS cohorts; we therefore (i) trained per-SV elastic-net predictors from flanking SNPs, (ii) formed an *augmented* summary-statistic and LD representation in which each SV occupies one row/column alongside the SNPs, and (iii) jointly fine-mapped SNPs and SVs per genomic region with the “regression with summary statistics” model SuSiE-RSS [Wang et al., 2020, Zou et al., 2022].

Two properties of these data determine the analysis design. First, the imputed-SV summary statistic is a deterministic linear combination of the same SNP statistics that are included in the LD matrix, so the augmented LD matrix is rank-deficient and the SV signal is collinear with its tagging SNPs (Sections S10.6–S10.7). Second, GWAS support is frequently missing across these low-complexity intervals, and how missing summary statistics are handled materially changes the result (Section S10.9). We therefore treat calibration (Section S10.10), power and limits on distinguishing SVs from their tag SNPs (Section S10.11), and an assessment of whether associations can be attributed specifically to SVs (Section S10.13) as part of the method.

#### S10.3 Datasets and accessions

All input datasets are public; accessions and versions are given in Table S15.

The 464 GWAS used in the phenome-wide application (Section S10.12) comprise 159 FinnGen endpoints and 305 Neale-lab UK Biobank phenotypes. We obtained or lifted all summary statistics to GRCh38, indexed them with tabix, and harmonized them to the panel allele orientation as described in Section S10.9.

#### S10.4 Structural-variant analysis set and elastic-net imputation

##### SV calling

We called structural variants from the *n* = 196 assembly-based cohort (1000 Genomes NYGC and HPRC assemblies) with SVIM-asm and harmonized the calls into a haplogroup-resolution catalog (main Methods). Resolution assigns each event one of four categories, named as in the released tables (non_decomposed, monophyletic, original_decomposed, residual); by design no de-duplication is performed across categories, so a genomic event may be included in the pipeline as more than one test row.

##### Per-SV elastic-net predictors

For each SV we trained an elastic-net regression of SV dosage on local SNP genotypes. Let **x** *∈* ℝ*^p^* be the (standardized) genotypes of *p* pericentromeric SNPs and *y* the SV dosage. We fit

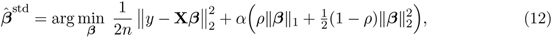

with the penalty mixing *ρ ∈ {*0.3, 0.5, 1.0*}* and strength *α* chosen by 4-fold cross-validation (`ElasticNetCV`, 50 candidate *α*, seed 42). We trained eight candidate models per SV (Section 4.5) and selected the model with the highest cross-validated *R*^2^ (CV *R*^2^) as the main predictor (best-of-eight). Two of the eight use SNPs anchored at the array midpoint, and an SV whose best-performing model was one of those two additionally has a sensitivity predictor built from the best of the remaining six (best-of-six); an SV whose two selections are identical contributes a single predictor. The unit of analysis is therefore the *predictor*, nested within the SV, and 11,129 candidate predictors are evaluated by the admissibility filter.

The coefficients ***β̂***^std^ are in standardized-genotype units and are used *directly* as the predictor weights ***β*** = ***β̂***^std^ throughout the cTWAS model (Section S10.6), paired with the reference *correlation* matrix **R**. This is the FUSION/cTWAS convention [Gusev et al., 2016, Zhao et al., 2024]: with both the predictor and **R** on standardized genotypes, the imputed-feature *z*-score 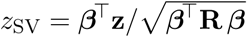 requires *no* per-SNP standard-deviation factor, which the full S-PrediXcan identity [Barbeira et al., 2018] includes for *per-dosage* PredictDB-style weights. The two parameterizations are *≈* 0.98 correlated under the null but can differ by up to *∼* 2*×* on individual realized statistics. CV *R*^2^ measures how well an SV is tagged by SNPs and therefore (Section S10.11) how distinguishable the SV is from those SNPs in fine-mapping.

##### Inclusion filter

The 11,129 candidate predictors all have CV *R*^2^ *≥* 0.1 from model selection. We applied three additional filters: autosomal only (chromosome X lacks a b38 LD reference; 79 excluded, leaving 11,050); at least one weight SNP matching the b38 LD-reference variant info on (chrom, pos, ref, alt) (4,075 excluded, leaving 6,975); and at least three matched weight SNPs within a single LDetect-b38 block (456 excluded). The filter includes **6,519 predictors on 5,149 distinct SVs** (4,195 best-of-eight, 2,324 best-of-six; 2,825 SVs by best-of-eight alone, 954 by best-of-six alone, 1,370 by both), the analysis set (Table S16).

Included predictors occupy 23 pericentromeric super-regions distributed over 21 chromosomes: chromosomes 6 and 11 each contribute two disjoint super-regions, and two chromosomes contribute none. Chromosome X is excluded by construction, and all 239 chromosome-14 candidates fail the LD-reference matching test. Their weight SNPs have a median of zero matched variants (maximum one), so no chromosome-14 predictor can be placed on the reference panel. This exclusion reflects reference-panel matching rather than low imputation accuracy: chromosome-14 candidate CV *R*^2^ has a median of 0.23 and reaches 0.95.

#### S10.5 LD reference panel

We computed the SNP–SNP correlation matrix **R** using **525 unrelated European founders** of the 1000 Genomes NYGC high-coverage panel (GRCh38), selected as samples with `FatherID=0 AND MotherID=0 AND Superpopulation=EUR`. We retained variants with minor allele frequency *≥* 0.01 and mean-imputed missing genotypes; **R** is the column-standardized Pearson correlation (unit diagonal). The LD-reference cohort is *decoupled* from the SV-imputation cohort: the elastic-net weights ***β*** are trained on the 188 HPRC individuals, whereas **R** is estimated on the EUR-525 founders, trading exact within-cohort LD for panel size and ancestry control. Its consequences for ancestry mismatch are tested directly in Section S10.13. We used the 1000 Genomes whole-genome-sequencing panel as the LD reference because pericentromeric SNPs are directly genotyped in high-coverage WGS, whereas array-based panels impute them. Coverage of these regions also limits the GWAS side: in the Neale-lab UK Biobank variant set, 9 of 22 chromosomes (2,076 of the 6,519 included predictors, 31.8%) have no flanking SNP present at any interval extension, and those predictors could not be tested regardless of which LD reference was used.

**Table S16:**
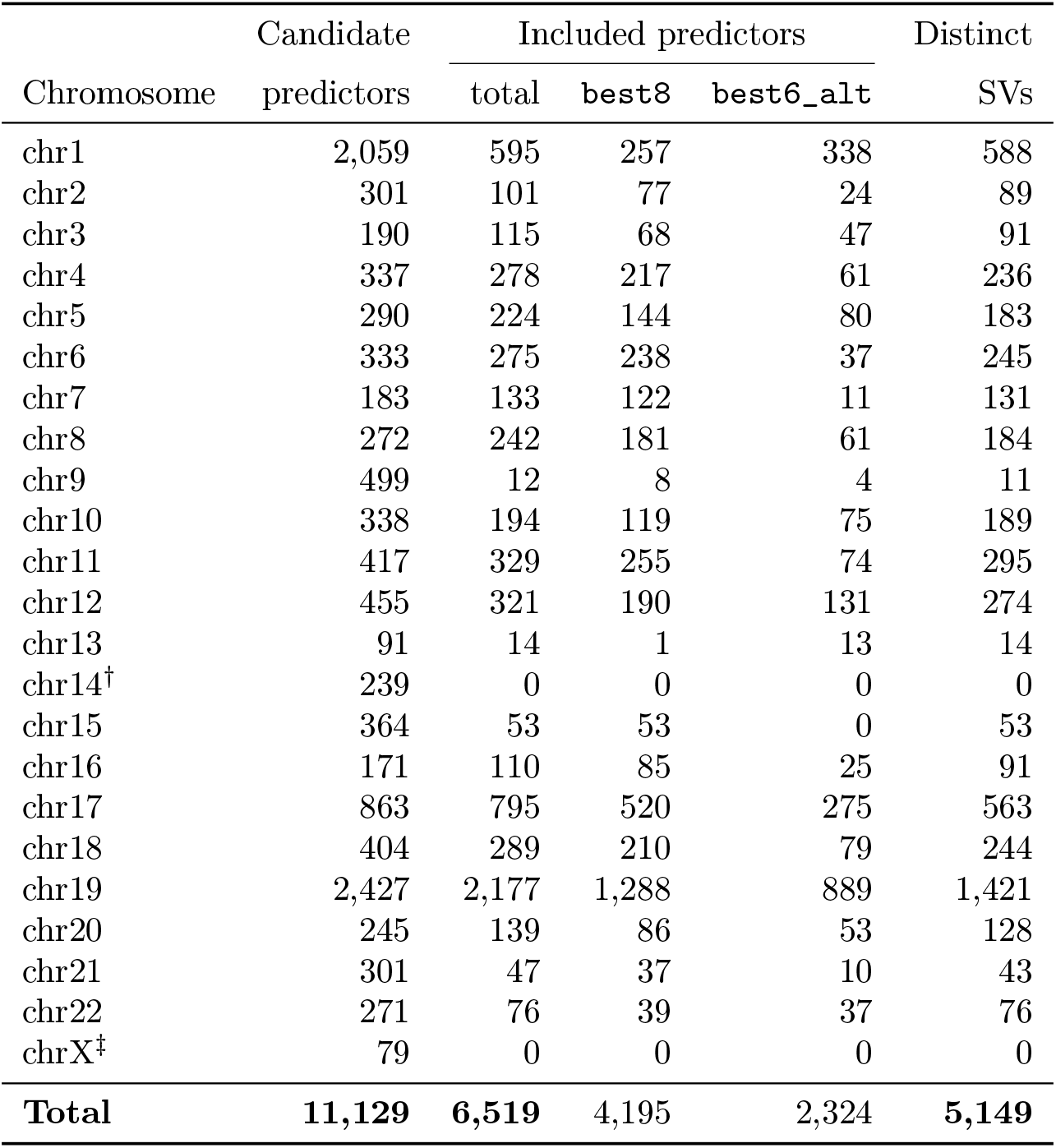
SV analysis set. Candidate predictors evaluated by the inclusion filter and those included, by chromosome. “Distinct SVs” merges the best-of-eight/best-of-six predictor pair into the underlying SV, so the total (5,149) is smaller than the predictor count (6,519). *^†^*chr14 is included nowhere because every candidate fails LD-reference matching; *^‡^*chrX is excluded for lack of a b38 LD reference.

| Candidate | Included predictors |  |  | Distinct |
|---|---|---|---|---|---|
| Chromosome | predictors | total | best8 | best6_alt | SVs |
|---|---|---|---|---|---|
| chr1 | 2,059 | 595 | 257 | 338 | 588 |
| chr2 | 301 | 101 | 77 | 24 | 89 |
| chr3 | 190 | 115 | 68 | 47 | 91 |
| chr4 | 337 | 278 | 217 | 61 | 236 |
| chr5 | 290 | 224 | 144 | 80 | 183 |
| chr6 | 333 | 275 | 238 | 37 | 245 |
| chr7 | 183 | 133 | 122 | 11 | 131 |
| chr8 | 272 | 242 | 181 | 61 | 184 |
| chr9 | 499 | 12 | 8 | 4 | 11 |
| chr10 | 338 | 194 | 119 | 75 | 189 |
| chr11 | 417 | 329 | 255 | 74 | 295 |
| chr12 | 455 | 321 | 190 | 131 | 274 |
| chr13 | 91 | 14 | 1 | 13 | 14 |
| chr14<sup>†</sup> | 239 | 0 | 0 | 0 | 0 |
| chr15 | 364 | 53 | 53 | 0 | 53 |
| chr16 | 171 | 110 | 85 | 25 | 91 |
| chr17 | 863 | 795 | 520 | 275 | 563 |
| chr18 | 404 | 289 | 210 | 79 | 244 |
| chr19 | 2,427 | 2,177 | 1,288 | 889 | 1,421 |
| chr20 | 245 | 139 | 86 | 53 | 128 |
| chr21 | 301 | 47 | 37 | 10 | 43 |
| chr22 | 271 | 76 | 39 | 37 | 76 |
| chrX<sup>‡</sup> | 79 | 0 | 0 | 0 | 0 |
| **Total** | **11,129** | **6,519** | 4,195 | 2,324 | **5,149** |

#### S10.6 The augmented cTWAS / SuSiE-RSS model

Within a super-region of *p* SNPs and *G* jointly fine-mapped SVs, the SNP GWAS *z*-scores **z** *∈* ℝ*^p^*, the SNP LD matrix **R** *∈* ℝ*^p×p^*, and the *G* predictor weight vectors ***β***_1_*, … ,* ***β****_G_* are combined into an augmented *z*-vector **z**_aug_ *∈* ℝ*^p^*^+^*^G^* and an augmented correlation matrix **R**_aug_ *∈* ℝ^(^*^p^*^+^*^G^*^)^*^×^*^(^*^p^*^+^*^G^*^)^ that treat each SV as a single “synthetic variant.” This follows the FUSION/S-PrediXcan imputed-association identity [Gusev et al., 2016, Barbeira et al., 2018] and the multi-feature canonical construction of cTWAS [Zhao et al., 2024], generalized to *G ≥* 1 correlated SVs (Supplementary Fig. S19a).

##### Imputed-SV *z*-score

Under the LD null the standardized association statistic of predicted SV dosage is the **R**-weighted burden of SNP *z*-scores,

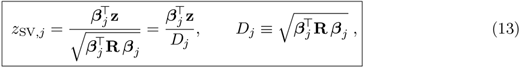

where *D_j_* is the standard deviation of the predicted dosage 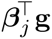 under Var(**g**) = **R**. Equation (13) is the S-PrediXcan imputed-association identity [Barbeira et al., 2018].

##### Augmented correlation matrix

Collect the weights into **B** = [***β***_1_*, … ,* ***β****_G_*] *∈* ℝ*^p×G^* and the normalizers into **D** = diag(*D*_1_*, … , D_G_*). The augmented matrix is

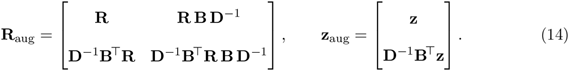

Entry-wise, the SNP*↔*SV block is the correlation of SNP *i* with predicted SV *j*,

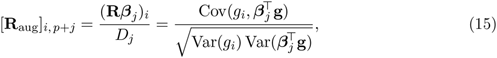

and the SV*↔*SV block is the predicted-dosage correlation

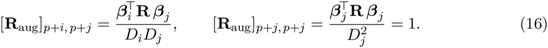

By Cauchy–Schwarz in the **R**-inner product, every off-diagonal entry of **R**_aug_ lies in [*−*1, 1] and the diagonal is exactly 1. So we can view **R**_aug_ as a valid correlation matrix of the augmented vector (Supplementary Fig. S19b). Its SV*↔*SV block encodes cross-SV LD, allowing the *G* SVs to be fine-mapped jointly.

#### S10.7 Rank deficiency and numerical safeguards

Writing **W** = [ **I***_p_ |* **BD***^−^*^1^ ] *∈* ℝ*^p×^*^(^*^p^*^+^*^G^*^)^, the construction in Eq. (14) is exactly a congruence transform

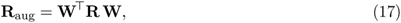

so **R**_aug_ is positive semi-definite whenever **R** is. **W** has *p* + *G* columns but only *p* rows, hence

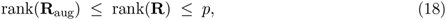

i.e. **R**_aug_ is **rank-deficient by construction**: each SV row is a deterministic linear combination of the SNP rows. This is the cTWAS augmentation identity, and it has two consequences. (i) Diagnostics computed *within* the augmented space (e.g. an `estimate_s_rss` *z*–**R** consistency check on the SV row) are not independent, because the SV row is by definition consistent with the SNP rows from which it was formed; such checks are implementation checks, not independent evidence. (ii) The imputed SV adds *zero independent degrees of freedom*: 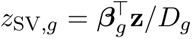 is a *deterministic* function of the SNP *z*-scores, with no independently measured component.

An imputed SV is therefore non-identifiable at the SV level: it can never be distinguished from the SNP linear combination it equals. It differs from a *measured* SNP, whose *z*-score, though correlated with its LD neighbours, has independent sampling variation given them (Var(*z_j_ | z_−j_*) *>* 0). Fine-mapping may still assign such an SV a high PIP when it is the most compact summary of a diffuse signal, but a high PIP then reflects *summarization*, not SV-specific causation. The question is thus one of *identifiability*, quantified in Sections S10.11– S10.13.

SuSiE-RSS operates directly on the unregularized rank-deficient canonical correlation matrix, whose smallest eigenvalue lies at numerical zero because the augmentation is rank-deficient by at least *G* (Eq. 18).

#### S10.8 SuSiE-RSS fine-mapping procedure

Joint fine-mapping used SuSiE-RSS [Wang et al., 2020, Zou et al., 2022], the sum-of-single-effects regression-with-summary-statistics model. The augmented effect vector is the sum of *L* single effects,

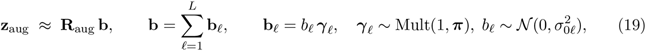

each ***γ****_ℓ_* a one-hot “which variable” indicator with posterior inclusion probabilities (the variational single-effect posterior) *α_ℓj_* = Pr(*γ_ℓj_* = 1 *|* data). The marginal posterior inclusion probability (PIP) of variable *j* (SNP or SV) is

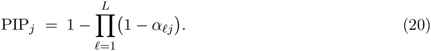

**Table S17:**
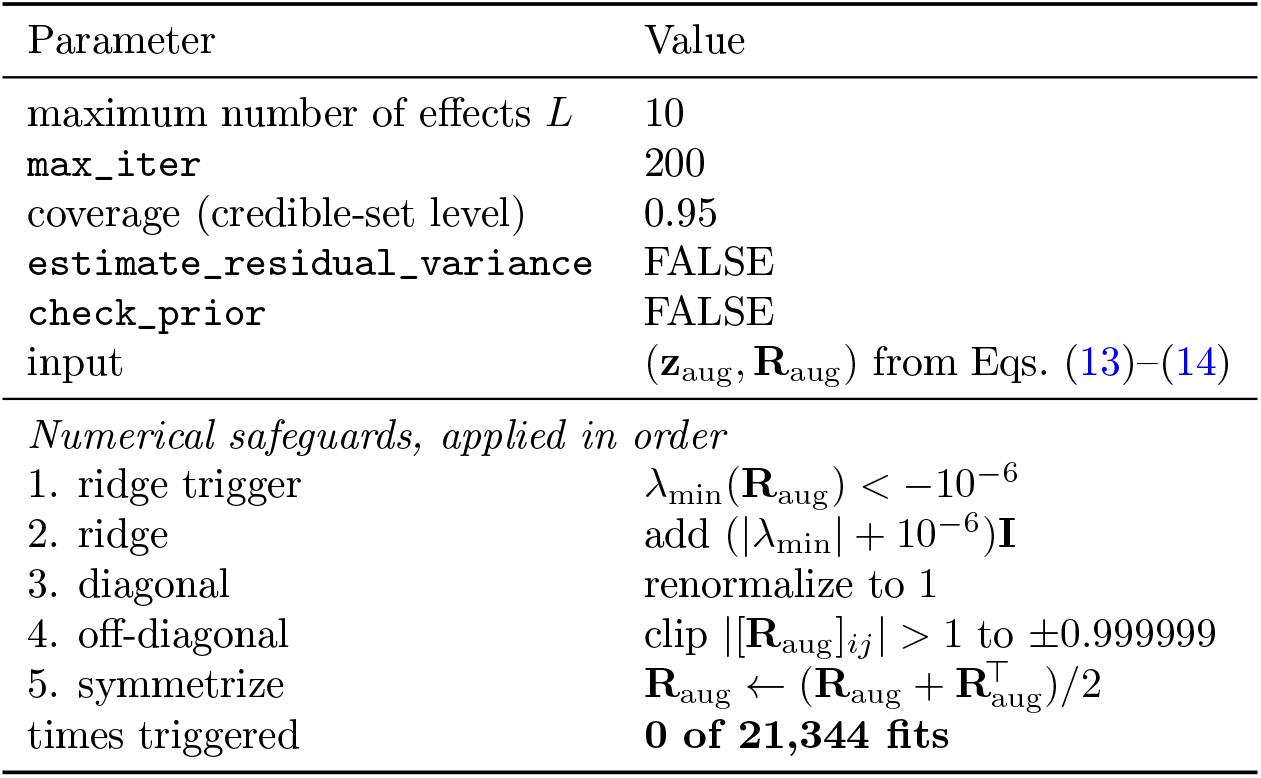
SuSiE-RSS settings. (susieR 0.14.2).

We report the last *G* PIPs as the SV PIPs and the first *p* as SNP PIPs. These settings (Table S17) were fixed before the phenome-wide application.

We keep estimate_residual_variance=FALSE because **R**_aug_ is a reference-panel (not in-sample) correlation matrix, the setting recommended for summary-statistic fine-mapping with an external LD reference [Zou et al., 2022]. Code, the complete analysis procedure and the **R**_aug_ derivation are given under Data and code availability.

#### S10.9 GWAS harmonization and missing-support handling

For each (super-region, GWAS) pair, we retrieved GWAS rows in the region with tabix and harmonized them to the panel: variant identifiers `chr_pos_ref_alt` were parsed to canonical `chrom:pos:ref:alt`, matched to panel SNP identifiers, and *z*-scores sign-flipped on REF/ALT swaps. In these regions many panel SNPs with a predictor weight are *absent* from the GWAS (for some SVs a median of *∼* 84% of the support is missing). We compared three approaches to missing summary statistics (Table S18) and treated their discordance as a diagnostic.

##### Summary-statistic imputation (IMPUTE)

We imputed missing SNP *z*-scores with the ImpG Gaussian-process estimator [Pasaniuc et al., 2014], implemented in its maintained successor ssimp 0.5.5 [Rüeger et al., 2018]: *ẑ_u_* = Σ*_ut_*(Σ*_tt_* + *λ***I**)*^−^*^1^*z_t_* for untyped variant *u* given typed variants *t*, with the ssimp default ridge 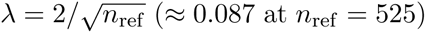 (*≈* 0.087 at *n*_ref_ = 525). Summary-statistic imputation is a *single-variant* estimator, and feeding imputed *z* into the *joint* SuSiE-RSS like-lihood risks miscalibration if the imputation uncertainty is not modeled [Kartau and Pirinen, 2025]; the calibration experiments quantify this (Section S10.10).

**Table S18:** Three approaches to missing GWAS summary statistics. “Support” = the SNPs with a non-zero predictor weight for a given SV.

| Handling | Definition | Status |
|---|---|---|
| PAD | missing-support SNP *z* set to 0 (zero-pad) | non-standard; miscalibrated (Section S10.10) |
| IMPUTE | missing-support SNP *z* filled by ImpG/ssimp summary-statistic imputation | calibrated, higher-power sensitivity analysis |
| MATCHED | predictor restricted to observed-support SNPs only (no pseudo-data) | calibrated; *main* (fewest assumptions) |

**Table S19:** Null calibration. (false-positive rate at PIP 0.8, 230 draws each). Parenthetical values are the maximum attainable PIP for the simulated SNP under that handling.

| Null design | PAD | IMPUTE (ImpG) | MATCHED |
|---|---|---|---|
| Global (pure noise, no signal) | 5.7% | 0.0% | 0.0% |
| Regional signal (one observed causal SNP, all SVs null) | 29% (67/230) | 0/230 (ceil. 0.21) | 0/230 (ceil. 0.56) |
| Missing causal lead (causal lead absent from the GWAS) | 24.3% | 0.43% (1/230) | 0.43% |

#### S10.10 Calibration: null designs and false-positive rates

We characterized the false-positive (FP) behavior of each handling with three nested null designs, each drawing 230 replicate *z*-vectors and recording FP@*τ* = Pr(PIP_SV_ *≥ τ |* no SV is causal) at *τ* = 0.8 (Table S19).

1. **Global null** (pure noise, no signal): **z** *∼ N* (**0**, **R**). Tests whether a handling produces false positives from noise.
2. **Regional-signal null** (one *observed* causal SNP *c*, all SVs null): 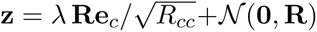. Tests whether real regional signal is *mis-attributed* to an SV.
3. **Missing-causal-lead null.** The causal lead is absent from the GWAS (*≥* 84% of predictor support missing). PAD assigns it a zero *z*-score, IMPUTE estimates it, and MATCHED excludes it.

PAD produces false positives from noise and assigns real regional signal to a collinear SV, reaching PIP *→* 1 on converged fits, whereas IMPUTE and MATCHED are well calibrated (FP *≤* 0.43%; Supplementary Fig. S20a); MATCHED is the main handling (fewest assumptions, no pseudo-data) and IMPUTE a higher-power sensitivity analysis, and because all nulls draw *z* from the LD used to impute, IMPUTE’s calibration covers single-causal, LD-consistent pericentromeric simulations.

**Table S20:** Power benchmark. = Pr(causal SV PIP *≥* 0.8) by SV signal strength *λ*. PAD additionally produces incorrect assignments of high PIP to SNPs (*≈* 0.71 at *λ* = 8).

| Handling | λ = 4 | λ = 6 | λ = 8 (*z*≈8) |
|---|---|---|---|
| PAD | 0.02 | 0.10 | 0.12 |
| IMPUTE (ImpG) | 0.03 | 0.17 | **0.35** |
| MATCHED | 0.02 | 0.17 | **0.27** |

#### S10.11 Power and limits of SV identifiability

To bound sensitivity we simulated each SV’s *EN predictor* as the causal genotype and measured power. The simulated causal unit is exactly the predicted dosage ***β****^⊤^***g**, so a real SV, which deviates from its predictor by imputation error, is harder to distinguish from SNP effects: this is an upper bound on identifiability for the augmented model. Simulating the predictor as causal induces SNP *z*-scores *z_i_* = (**R*β***)*_i_λ/D* with E[*z*_SV_] = *λ*; we swept the signal strength *λ ∈ {*4, 6, 8*}* (230 draws each) and recorded power(*τ*) = Pr(causal-unit PIP *≥ τ*) (Table S20; Supplementary Fig. S20b,c).

Power is low even at *z ≈* 8 (*≤* 35%), and power *decreases* with imputability: SVs with CV *R*^2^ *≥* 0.6 reach the PIP threshold in only 5–10%, versus 31–40% for CV *R*^2^ *<* 0.4. This is an **identifiability ceiling**, not a weak-signal artifact: a well-imputed SV is well-tagged by SNPs, so fine-mapping distributes the posterior probability across the SV and its correlated tag SNPs, leaving no single variant with high PIP. Increasing sample size raises 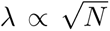 but does *not* remove the CV *R*^2^ ceiling for well-imputed SVs. Two corollaries follow: (i) a high-PIP imputed SV is preferentially a *poorly* imputed one (low CV *R*^2^), and so the SV least trustworthy as a genotype proxy; (ii) testing more traits does not by itself resolve this ambiguity, even in larger cohorts.

#### S10.12 Phenome-wide application

We applied the calibrated pipeline (IMPUTE and MATCHED) across all 23 pericentromeric super-regions and all 464 GWAS (159 FinnGen + 305 Neale-lab UK Biobank), i.e. 23 *×* 464 = 10,672 region*×* trait fits per handling (21,344 total).

**Table S21:** Candidate selection in the phenome-wide analysis. Counts are of (super-region, trait, SV) combinations. The two approaches differ only in how GWAS-unmatched reference SNPs are treated, so the marginals and the exclusive counts are given together: 411 and 706 SV–trait pairs reach PIP 0.5 under IMPUTE and MATCHED respectively, of which 222 do so under both.

|  | IMPUTE | MATCHED | both / total |
|---|---|---|---|
| super-region × GWAS fits | 10,672 | 10,672 | 21,344 |
| stored (region, trait, SV) rows at PIP ≥ 0.01 |  |  | 160,388 |
| with a finite posterior under both approaches |  |  | 159,751 |
| with no finite posterior under one |  |  | 637 |
| SV–trait pairs at PIP ≥ 0.5 | **411** | **706** | **222** |
| under that handling alone | 189 | 484 | – |
| distinct SVs among them | 76 | 167 | 33 |
| *Triage of the 222 SV–trait pairs reaching the PIP threshold under both approaches* |  |  |  |
| candidate (converged, at most three SVs with high PIP) |  | **39** rows | **24** SVs |
| flagged, more than three SVs with high PIP |  | 165 rows | 5 SVs |
| flagged, fit did not converge |  | 18 rows | 6 SVs |

For each fit we stored every variable with PIP *≥* 0.01, a per-(region, trait, handling) summary, and an automated selection across the two approaches (Table S21). We selected a row as a candidate only if it met thresholds under both approaches (IMPUTE PIP *≥* 0.5 *and* MATCHED PIP *≥* 0.5), the fit converged, and at most three SVs in the region*×* trait reached PIP *≥* 0.5. Several correlated SV rows can reach high posterior in one super-region because each decomposition category is included separately (Section S10.4), so one genomic event may appear in a fit more than once. More SV–trait pairs reach the PIP threshold under MATCHED than under IMPUTE (Table S21), because restricting the predictor to SNPs with available summary statistics changes the predicted dosage. A MATCHED predictor therefore represents only the component of SV dosage predicted by the observed SNPs. The two approaches share weights **B** and LD **R**, so their agreement measures robustness to the treatment of missing summary statistics.

#### S10.13 Identifiability audit: own-tag conditioning and ancestry-matched LD

We assessed each candidate SV to determine whether its signal is specific to the SV or reflects tagging SNPs.

##### Own-tag conditioning

For SV *g* we ranked SNPs by their augmented correlation with the SV, *|*[**R**_aug_]*_i, p_*_+_*_g_|* = *|*(**R*β****_g_*)*_i_|/D_g_*, and took the top-*K* (*K ≤* 40) as the SV’s own tags *S*. Using the multivariate-normal conditional under **R**_aug_, the SV *z*-score conditioned on its own tags is

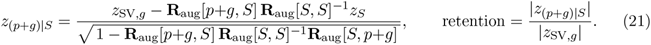

A signal with low retention is a proxy for its own tag SNPs (SNP-non-identifiable). High retention is consistent with an SV-specific signal but does not establish one, so the screen is one-sided.

Two controls calibrate it. As a positive control, a known proxy deletion on chromosome 3 falls to 7% retention at the full tag set (*K* = 40). As a negative control for *specificity*, simulating a single causal tag SNP (a true single-SNP proxy) reduces retention to 0.000 at every *K*, whereas simulating the SV’s own predictor as the causal unit retains 0.3–0.8 (mean 0.62 at *K*=10). The known proxy shows marked attenuation after conditioning, whereas unresolved candidates retain more of their association, so the audit separates the two. The distinction is less clear at low CV *R*^2^ (a simulated causal low-CV-*R*^2^ SV can retain as little as 0.32), and at very low panel size the **R**_aug_[*S, S*]*^−^*^1^ inverse is numerically unstable.

##### Ancestry-matched LD

For the candidates surviving own-tag conditioning, we rebuilt **R** from 1000 Genomes subsets matched to the GWAS ancestry (FIN *n* = 99; EUR *n* = 503), holding ***β*** and **z** fixed so that only the denominator *D_g_* in Eq. 13 changes. An internal control (1000 Genomes EUR *≈* the EUR-525 reference panel) agreed, *z*_SV_ moved by at most 18% across the surviving candidates, and at *n* = 99 the own-tag inverse was numerically unstable (retention *>* 2000%); FinnGen in-sample LD or direct WGS or long-read SV genotyping is the decisive test of SV-specificity.

Of the 24 candidates, 21 were SNP-non-identifiable (12 whose associations attenuated after conditioning on their tag SNPs, 7 explained by a stronger regional SNP, and 2 SNP-equivalent at CV *R*^2^ *≥* 0.6), and three remained *unresolved* (chr12 sv721 with standing height, CV *R*^2^ = 0.15, *z*_SV_ = 7.04 against a strongest regional SNP of 5.77; chr11 sv263 with BMI, 0.43, *−*6.26 against 4.86; chr11 sv271 with serum albumin, 0.41, *−*6.02 against 4.33), each with *|z*_SV_*|* above any single regional SNP, retained under own-tag conditioning and stable under ancestry-matched LD. The result is therefore **identifiability-limited** (21 SNP-non-identifiable and 3 WGS-unresolved candidates), and determining whether the signal is specific to an SV at these pericentromeric loci requires WGS or long-read SV genotyping.

##### Dense-panel identifiability validation

To distinguish identifiable SV signals from proxies for tagging SNPs, every candidate SV–trait association from the single-SV and the joint multi-SV analyses (6,519 imputable-SV predictors *×* 464 UK Biobank/FinnGen GWAS) was re-fine-mapped against a densified panel: the SV’s own predictor SNPs augmented with the strongest same-region GWAS SNPs, an additional set of SNPs with the largest *|z|* values, capped near 12,000 variants. Fits used `susie_rss` (*L* = 10) in both the matched and ImpG-imputed *z* constructions and at CV *R*^2^ *≥* 0.3 and *≥* 0.1 inclusion thresholds, and an SV was retained at PIP *>* 0.5. SV–trait pairs at SV PIP *≤* 0.5 were classified as SNP-supported (a SNP at PIP *>* 0.5), fits that converged without resolving a credible variant, and non-converged fits. Three SVs retained PIP *>* 0.5, on chromosomes 11 and 19. A positive-control power calibration across the CV *R*^2^ range was not performed.

**Figure S19:**
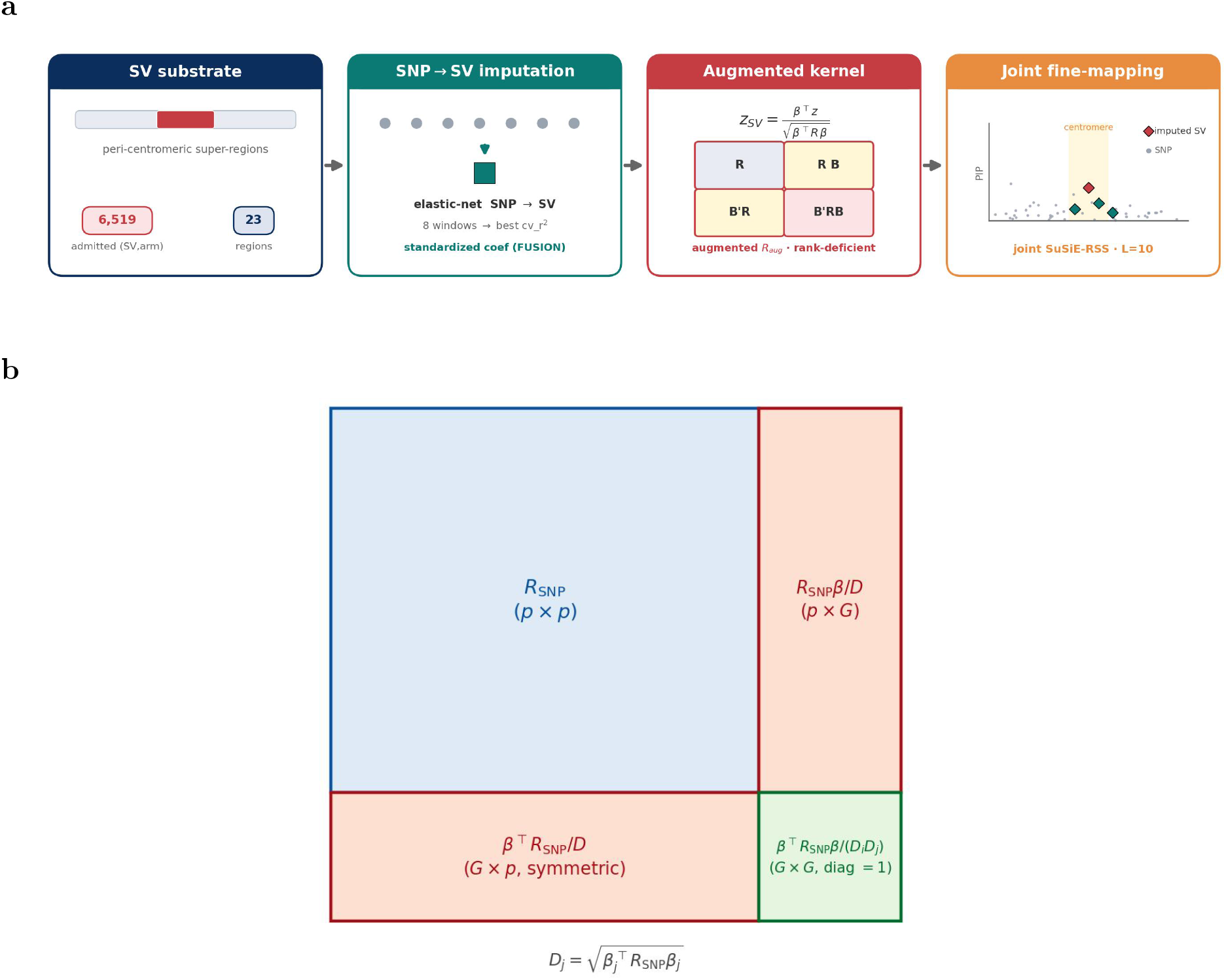
The imputed-SV joint fine-mapping method, and the block structure of R_aug_. **a** Per pericentromeric super-region, included SVs (6,519 predictors over 23 regions) are imputed from local SNPs by elastic net (standardized coefficients, FUSION convention), added to the augmented correlation matrix **R**_aug_ (in which the SV occupies one rank-deficient row, 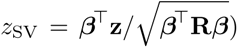 and jointly fine-mapped with the SNPs by SuSiE-RSS (*L*=10). The phenome-wide analysis applies this method to 464 traits 23 regions (10,672 region trait fits per handling); the successive selection steps (222 robust region trait hits, 24 unique candidate SVs, 3 surviving own-tag conditioning and ancestry-matched LD, and no SV-specific signal identifiable) is given in the text and tables. **b** The augmented model of panel a at full size. The *p* SNPs of a region contribute the reference LD block **R** (1000 Genomes EUR, *n* = 525); each of the *G* imputed SVs adds one row and column built analytically from its elasticnet weights ***β***, since reference panels contain no observed SV genotypes. The SNP–SV block is **R*β****/D* and the SV–SV block ***β****^⊤^***R*β****/*(*D_i_D_j_*), with 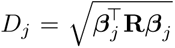 the induced SV standard deviation, so the SV–SV diagonal is 1 by construction. Because every SV row is a linear combination of SNP rows, **R**_aug_ is rank-deficient by construction, the algebraic origin of the non-identifiability quantified in this note.

**Figure S20:**
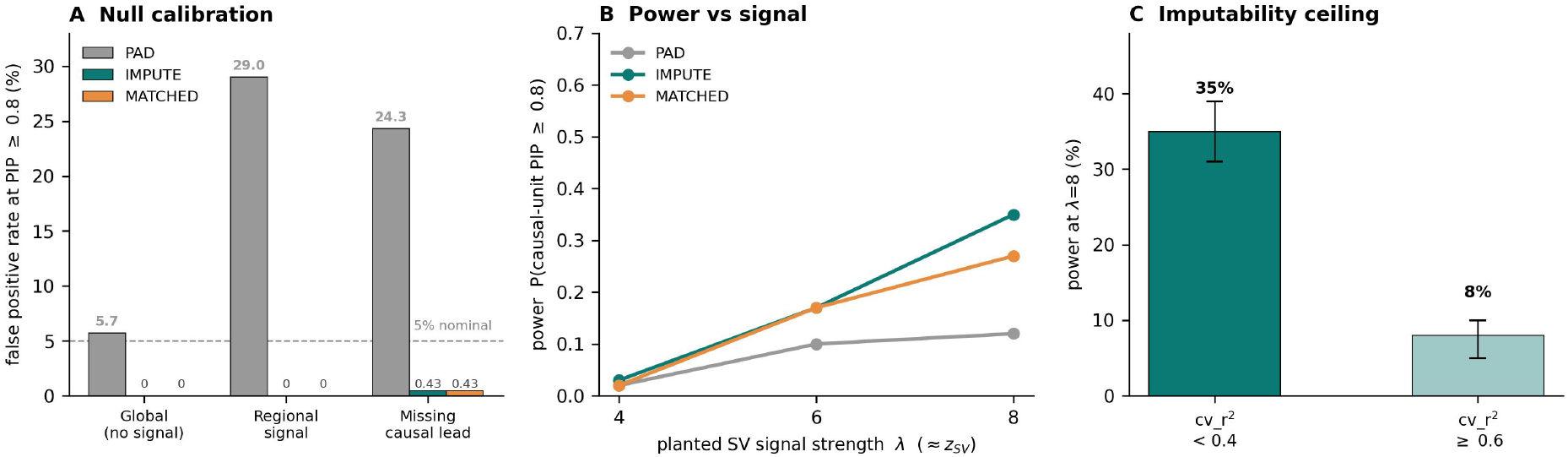
Calibration and the identifiability ceiling. **a** Null false-positive rate at PIP 0.8 across the three null designs by handling: zero-padding (PAD) is miscalibrated; IMPUTE and MATCHED are well-calibrated. **b** Power = Pr(causal-unit PIP 0.8) against simulated SV signal strength *λ*; PAD additionally assigns high PIP to noncausal SNPs. **c** Power at *λ*=8 falls sharply with imputability (CV *R*^2^), because better-imputed SVs are more tightly correlated with SNP predictors and less often receive high individual PIP. Values: Tables S19–S20.

### Supplementary Note 11 Sensitivity analyses

#### Merged CDR calls reproduce the chromosome-8 association

Gao et al. [2026] merge hypomethylated calls within 150 kb into a single CDR; ours are the unmerged dips. Applying their merge reduces 22,537 dips across 5,034 active arrays to 5,400 CDRs, a mean of 1.07 per array, and widens the median call from 30 kb to 215 kb.

We recomputed every phenotype from the merged calls and repeated the association analysis with the same genotypes, covariates, permutation seed and null. The chromosome-8 lead is unchanged (*t* = *−*6.173 against *t* = *−*6.172) and the study-wide significant set for dip position is identical on chromosomes 8 and 5, with no locus gained (Supplementary Fig. S15); of the four chromosome-8 loci in Extended Data Table 1 the 1,868 bp insertion alone loses significance. After merging, the two position phenotypes are identical on 93.3% of arrays, against 17.4% before.

#### Local methylation intervals

Testing methylation in intervals around each SV assesses local associations separately from the array-wide mean (Supplementary Note 7). In 192 individuals with assembly-derived SV genotypes, we tested each SV against mean 5mC in *±*10, *±*50 and *±*100 kb intervals around it. The tests used Minigraph–Cactus chains or the SVIM-asm alignments used for SV calling, with at least 50 individuals per test. We applied Bonferroni correction within each interval width and, in a sensitivity analysis, across all three widths; we assessed significance using 10^7^ permutations. We required each candidate to show no significant association with sequencing depth, and used a second control to test whether SV dosage predicted the availability of a methylation value. In intervals around each SV, three loci reach the threshold at 10^7^ permutations in the SVIM-asm alignments, which make 9,889–10,499 loci testable; the Minigraph–Cactus chain spans *α*-satellite on a subset of chromosomes, reaches *n ≥* 50 at 12.4% of the catalog with chromosomes 1, 4 and 17 untested, and gives a null result at every scale. The three loci are an insertion at chr11:54,314,992 (*P* = 1–2 *×* 10*^−^*^6^), an insertion at chr17:26,767,558 and a deletion at chr19:28,468,779, and all three are associated with lower methylation at higher SV dosage. The chromosome-11 and chromosome-19 associations remain significant after correction across the three interval widths. The coverage negative control showed a *larger* excess of associations than methylation at every interval width.

#### Measurement presence in the interval-based test

The interval-based test of local methylation includes a coverage negative control, but we compute that control only among samples that produced a value, so the control conditions on measurement presence. We repeated the test’s design with measurement presence as the outcome, on the same 11,129 loci and 150 individuals under the same covariates and Freedman–Lane permutation null. SV dosage predicts whether an interval has a methylation value at 14.7%, 12.6% and 12.3% of loci at *±*10, *±*50 and *±*100 kb, against a 5% nominal rate and against 8.2%, 8.4% and 8.2% for the depth control on the same loci (genomic control factors 1.84, 1.60 and 1.54). The direction depends on interval width: among significant loci, 27.4% have a negative slope at *±*10 kb against 57.8% at *±*100 kb. Both insertions and deletions show associations in either direction: at *±*10 kb 32.2% of significant deletions and 20.8% of significant insertions have a negative slope. Measurement presence tracks dosage in excess of the depth control at every interval width, so we report the chromosome-11, -17 and -19 loci as candidates.

#### Filter sensitivity comparison

For each trait, we computed the genomic-control factor *λ*_GC_ = median(*z*^2^)*/*0.4549 over all (SV, trait) pairs for that trait, deflated each *z* to 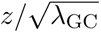, and applied Benjamini–Hochberg to the corrected *P* values at *q <* 0.05. A pair qualified as a candidate if it also passed the `estimate_s_rss` LD-consistency filter and CV *R*^2^ *≥* 0.3 in both analyses. The *≥* 0.1 analysis therefore differs only in which predictors enter the joint fits and in each trait’s test family: *z* is identical in all 2,957 SV–trait pairs common to the two analyses, and candidate status changes only because including additional predictors changes the calibration and multiple-testing correction.

Across the 191 traits tested in both, *λ*_GC_ rose in 60 of the 70 traits that lost candidate SV– trait pairs (median ratio 1.15) and changed little in the 22 that gained (median 0.99; Spearman *ρ* = *−*0.69 between the *λ*_GC_ ratio and the change in the number of candidates); because a per-trait rescaling is monotone, no trait both gained and lost.

### Supplementary Note 12 Population differentiation of centromeric SVs

#### S12.1 Highly differentiated QTL SVs cluster at a few loci, most prominently on chromosome 10

Structural variants can track population structure [Almarri et al., 2020], which makes allele-frequency differentiation a relevant property of each SV associated with expression or splicing. Each SV is described by its maximum pairwise *F_ST_* across the five 1000 Genomes superpopulations (Section S12.2).

Centromeric SVs show low population differentiation. Across the full catalog the mean *F_ST_* was 0.0164, 0.58% had *F_ST_ >* 0.15, and the most differentiated variant reached *F_ST_*= 0.580 (Supplementary Fig. S21). On average, imputable SVs were more differentiated (mean 0.0238; 1.47% above 0.15), and that excess was spread across centromeres and imputation-accuracy levels (Supplementary Fig. S21). At the stringent imputability threshold (CV *R*^2^ *≥* 0.3), SVs associated with expression or splicing were enriched 2.9-fold for high-*F_ST_* variants relative to SVs passing the same threshold (6 of 106 above *F_ST_* = 0.15, 5.66% against a 1.99% background; one-sided Fisher *P* = 1.7 *×* 10*^−^*^2^); at the permissive threshold (CV *R*^2^ *≥* 0.1) the corresponding ratio was 1.4 (4 of 182, 2.20% against 1.62%; *P* = 0.34).

The most differentiated hits cluster at a few loci. A chromosome-10 insertion at *∼*39.6 Mb (*F_ST_*= 0.580, depleted on African haplotypes) tags the *ZNF33A* association, an adjacent 1,357-bp insertion 163 bp away (*F_ST_*= 0.292) tags *TMEM273* /*HSD17B7P2* /*GTF2IP*, and further differentiated SVs lie on chromosomes 1 and 6 (*F_ST_ ≈* 0.20). The SVs associated with *SUV39H1* (*F_ST_* = 0.00) and the chromosome-20 *TTPAL*/*PLTP* /*NEURL2* splicing trio (*F_ST_ ≤* 0.04) fall in the undifferentiated majority. Population differentiation among QTL SVs is concentrated in a small set of loci, most prominently on chromosome 10.

#### S12.2 Population differentiation analysis

To assess whether centromeric-SV associations reflect population structure, we computed the single-locus *F_ST_* for each SV using the Hudson estimator [Bhatia et al., 2013], from directly called HPRC SVIM-asm genotypes rather than imputed dosages. We computed SV allele frequencies in the 188 HPRC training individuals, partitioned into five 1000 Genomes superpopulations (AFR = 55, AMR = 39, EAS = 39, SAS = 34, EUR = 21). For a pair of superpopulations (*i, j*) with alternate-allele frequencies *p_i_, p_j_* over *n_i_, n_j_* diploid individuals, the Hudson estimator is

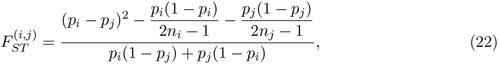

with each frequency estimated as *p* = Σ*_g_ g /* (2*n*) over diploid genotypes *g ∈ {*0, 1, 2*}*, so that the sampling-variance correction applies to the 2*n* sampled gene copies. We summarized each SV by its maximum *F_ST_* across the ten superpopulation pairs and classified SVs with *F_ST_ >* 0.15 as highly differentiated. We retained negative per-pair estimates, which arise as sampling noise when differentiation is near zero, without truncation. We tested enrichment of QTL hits for differentiated SVs with a one-sided Fisher exact test on the 2 *×* 2 table of (hit vs. non-hit) *×* (high-vs. low-*F_ST_*) over the imputable SV analysis set, separately for the permissive (CV *R*^2^ *≥* 0.1) and strict (CV *R*^2^ *≥* 0.3) imputability levels. We matched QTL hits to the frequency table using the identifier the QTL analysis itself assigns each tested unit (chromosome and within-chromosome SV index), which is one-to-one.

**Figure S21:**
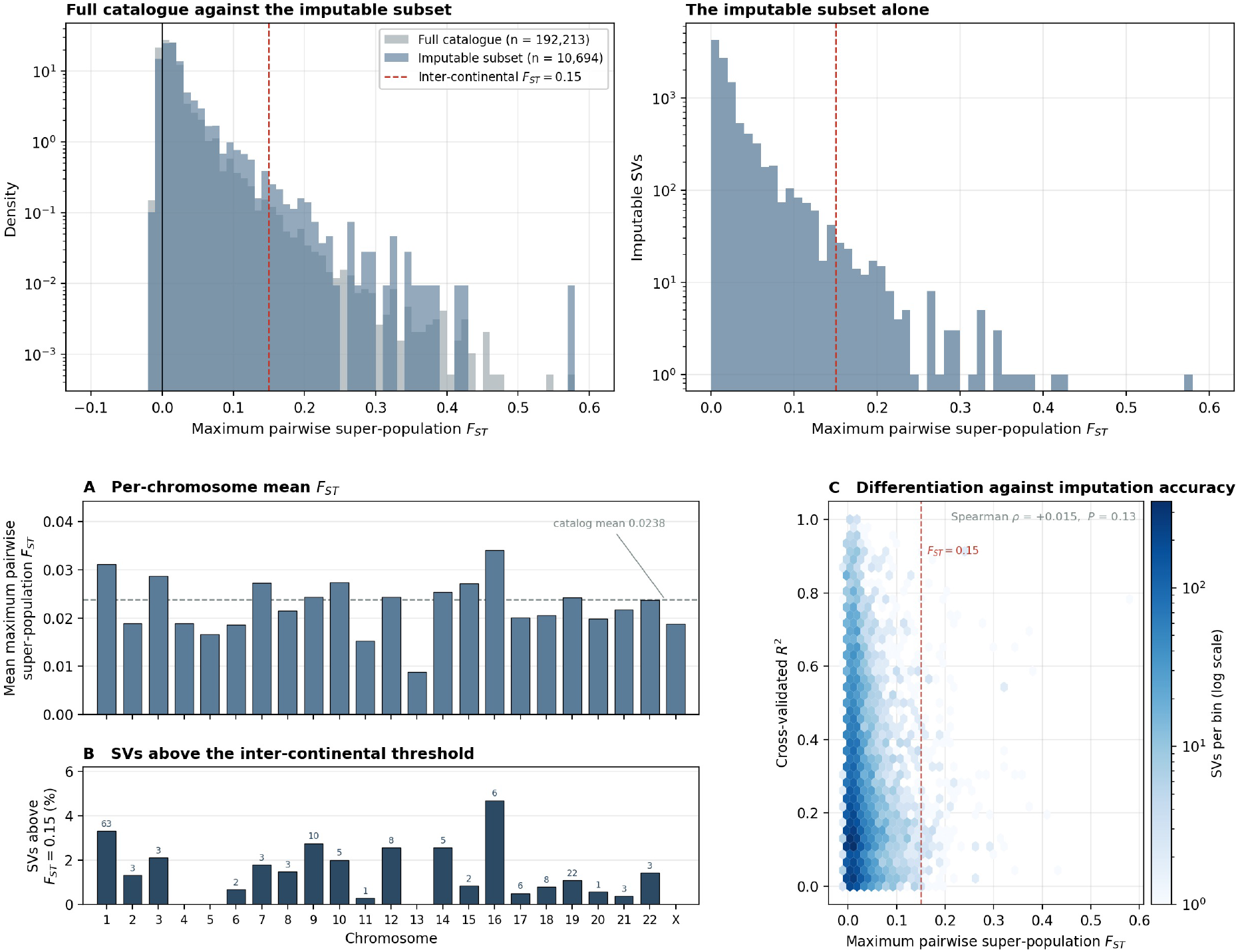
Population differentiation of centromeric SVs is low and uncorrelated with imputation accuracy. Maximum pairwise superpopulation *F_ST_*(Hudson estimator, sample-size corrected) for the full catalog (192,213 SVs with a defined *F_ST_*) and for the imputable subset (10,694), both concentrated near zero at means 0.0164 and 0.0238. The imputable subset includes 157 highly differentiated SVs (1.47%) above *F_ST_*= 0.15, reaching 0.580. A dashed line marks the threshold for high differentiation, *F_ST_*= 0.15. Negative values are retained. **a** Mean maximum pairwise superpopulation *F_ST_* by chromosome for the 10,694 imputable SVs. A dashed line marks the subset mean, 0.0238, and chromosome 16 reaches the highest mean, 0.0340. Per-chromosome counts range from 57 (chromosome X) to 2,006 (chromosome 19). **b** Percentage of SVs above *F_ST_* = 0.15 by chromosome, SV counts above each bar. Chromosomes 4, 5, 13, and X contribute none of the 157. **c** *F_ST_* against cross-validated imputation *R*^2^, uncorrelated at Spearman *ρ* = +0.015, *P* = 0.13.

### Supplementary Note 13 Accuracy of SVIM-asm calls inside active HOR arrays

#### Simulation design

We simulated CHM13 chromosomes 1–22 and X with clustered substitutions at 0, 1 and 2% in the active HOR array and 0.1% elsewhere. We introduced 12–15 SVs of known sequence per simulated chromosome (1,017 in total), comprising insertions, deletions, tandem and dispersed duplications, inversions and complex replacements of 50 bp to 100 kb with *α*-satellite inserted sequence (Supplementary Table S22). A matched negative control for each chromosome contained no simulated SVs. We aligned all chromosomes to CHM13 and processed them with the catalog pipeline (Methods).

#### Scoring

Because the same sequence change can have several equivalent representations within a tandem repeat, we scored the reconstructed sequence rather than requiring identical breakpoint coordinates. We considered a simulated SV detected when the reported calls, applied to the reference, reconstructed its sequence exactly; we accepted shifted breakpoints along the repeat and split representations. With *P* the simulated SVs, *E* the reported calls and *U* the negative-control units (whole arrays and 50-kb windows),

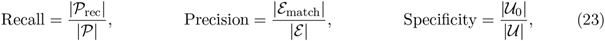

where *P*_rec_ are detected SVs, *E*_match_ calls contributing to a reconstruction and *U*_0_ units without calls. We scored calls with an unresolved match (101 of 2,055) as incorrect. Recall measures the fraction of introduced changes reconstructed exactly, whereas precision measures the fraction of reported calls contributing to those reconstructions. Thus, a large event reported as several incorrect small calls can reduce both recall and precision.

#### Accuracy in simulation

Recall decreased with SV size and background divergence, and the pipeline detected no inversions or complex replacements (Supplementary Fig. S22b; Supplementary Table S22). Precision varied widely among chromosomes (Supplementary Fig. S22a), and the caller frequently represented large simulated SVs as multiple small incorrect calls. Truvari with the settings of the HG002 comparison yielded a similar precision (Table S2). Negative controls were free of calls in 59 of 69 arrays. The low precision despite few calls in negative controls suggests that alignment and representation of true structural changes within repeats contribute to errors. Agreement between assemblies of the same individual (Supplementary Note 1) therefore measures reproducibility, which can exceed accuracy against known sequence changes.

#### Frequency filtering

In a simulated cohort of 256 haplotypes merged and filtered as for the catalog, carrier-level precision increased from 30% to 53% at a minimum minor allele frequency of 5%, whereas recall of rare SVs decreased (Supplementary Fig. S22c). Haplotypes of the same lineage frequently shared incorrect common calls. Frequency filtering therefore enriches for accurate calls but also retains systematic errors shared within lineages. Such calls may predict haplotype membership without correctly specifying the underlying structural change; imputation accuracy alone cannot establish sequence-level accuracy.

#### Sequence composition and length periodicity

We annotated the changed sequence of each call (inserted sequence for insertions, removed CHM13 sequence for deletions) with RepeatMasker 4.1.9; we classified calls with *≥*80% of these bases annotated as *α*-satellite or trans-posable element accordingly. We considered a length *L* periodic at period *P* and tolerance *t* when min*_k≥_*_1_ *|L − kP | ≤ t*; control periods and the expectation under uniformly distributed lengths, (2*t* + 1)*/P*, provide null comparisons (Supplementary Table S23; Supplementary Fig. S22d). Enrichment near multiples of 171 bp is consistent with gains and losses of *α*-satellite monomers.

**Table S22:** Recall of simulated SVs by type, size and background substitution rate. (Supplementary Note 13). Detected/simulated over all 23 chromosomes; a simulated SV is detected when the reported calls reconstruct its sequence exactly.

| Type | Size | 0% | 1% | 2% | All |
|---|---|---|---|---|---|
| Insertion | 50 bp–1 kb | 63/69 | 62/69 | 62/69 | 187/207 (90%) |
| Insertion | 1–10 kb | 15/23 | 15/23 | 13/23 | 43/69 (62%) |
| Insertion | 10–50 kb | 9/21 | 9/21 | 5/21 | 23/63 (37%) |
| Deletion | 50 bp–1 kb | 43/46 | 41/46 | 37/46 | 121/138 (88%) |
| Deletion | 1–10 kb | 13/23 | 14/23 | 9/23 | 36/69 (52%) |
| Deletion | 100 kb | 2/21 | 1/21 | 2/21 | 5/63 (8%) |
| Tandem duplication | 50 bp–1 kb | 42/46 | 40/46 | 37/46 | 119/138 (86%) |
| Tandem duplication | 100 kb | 5/21 | 1/21 | 0/21 | 6/63 (10%) |
| Dispersed duplication | 680 bp | 21/23 | 19/23 | 18/23 | 58/69 (84%) |
| Inversion | 10 kb | 0/23 | 0/23 | 0/23 | 0/69 (0%) |
| Complex replacement | 50 bp–1 kb | 0/23 | 0/23 | 0/23 | 0/69 (0%) |

**Figure S22:**
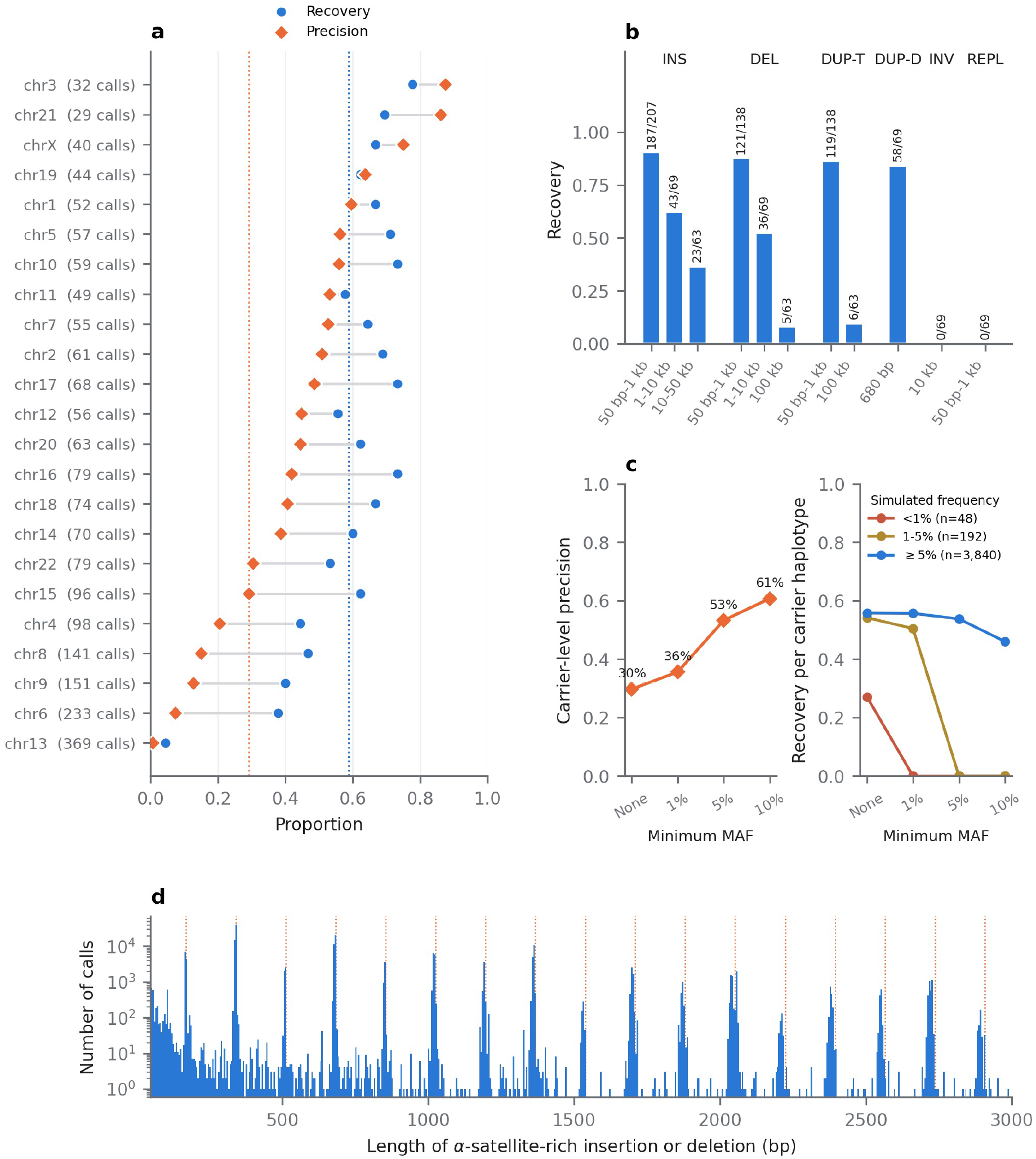
Accuracy of SVIM-asm calls in simulated centromeres and length periodicity of *α*-satellite indels in the catalog. (Supplementary Note 13). **a** Recall of simulated SVs and precision of reported calls per chromosome, pooled across substitution rates (0, 1 and 2%) and negative controls. Chromosomes are ordered by precision; the number of reported calls is given in parentheses; dotted lines indicate pooled values. **b** Recall by SV type and size; numbers above bars give detected/simulated SVs. INS, insertion; DEL, deletion; DUP-T, tandem duplication; DUP-D, dispersed duplication; INV, inversion; REPL, complex replacement. **c** Carrier-level precision (left) and recall per carrier haplotype by simulated carrier frequency (right) in a simulated cohort of 256 haplotypes, after the catalog filters and an additional minimum minor allele frequency. **d** Length distribution of *α*-satellite-rich insertions and deletions of 50 bp to 3 kb in the catalog; dotted lines indicate multiples of 171 bp.

**Table S23:** Length periodicity of *α*-satellite-rich indels. (Supplementary Note 13). Proportion of *α*-satellite-rich insertions and deletions of at least 50 bp whose length lies within *t* of an integer multiple (*k ≥* 1) of period *P*. The expected proportion under a uniform length distribution is (2*t* + 1)*/P*.

| Period *P* (bp) | *t* = 5 bp | *t* = 10 bp | *t* = 17 bp |
|---|---|---|---|
| **171** | 64.6% | 83.7% | 92.0% |
| 150 | 1.6% | 8.7% | 17.6% |
| 157 | 2.0% | 6.1% | 12.2% |
| 177 | 0.6% | 6.2% | 38.5% |
| 185 | 1.7% | 2.8% | 6.9% |
| 190 | 0.5% | 2.0% | 4.2% |
| Uniform length distribution (*P* = 171) | 6.4% | 12.3% | 20.5% |
| *By length (P = 171 bp, t = 10 bp)* |  |  |  |
| 160 bp–1 kb |  | 99.2% |  |
| 1–2 kb |  | 81.4% |  |
| 2–5 kb |  | 30.4% |  |
| >5 kb |  | 5.0% |  |

## References

Kara L McKinley and Iain M Cheeseman. The molecular basis for centromere identity and function. Nature Reviews Molecular Cell Biology, 17(1):16–29, 2016. doi: 10.1038/nrm.2015.5.

Steven Henikoff, Kami Ahmad, and Harmit S Malik. The centromere paradox: stable inheritance with rapidly evolving DNA. Science, 293:1098–1102, 2001.

Nicolas Altemose, Glennis A Logsdon, Andrey V Bzikadze, Pragya Sidhwani, Sasha A Langley, Gina V Caldas, Savannah J Hoyt, Lev Uralsky, et al. Complete genomic and epigenetic maps of human centromeres. Science, 376(6588):eabl4178, 2022.

Karen H Miga and Ivan A Alexandrov. Variation and evolution of human centromeres: a field guide and perspective. Annual Review of Genetics, 55:583–602, 2021.

Sergey Aganezov et al. A complete reference genome improves analysis of human genetic variation. Science, 376(6588):eabl3533, 2022. doi: 10.1126/science.abl3533.

Sergey Nurk, Sergey Koren, Arang Rhie, Mikko Rautiainen, Andrey V Bzikadze, Alla Mikheenko, Mitchell R Vollger, Nicolas Altemose, et al. The complete sequence of a human genome. Science, 376(6588):44–53, 2022. doi: 10.1126/science.abj6987.

Wen-Wei Liao, Mobin Asri, Jana Ebler, Daniel Doerr, Marina Haukness, Glenn Hickey, Shuangjia Lu, Julian K Lucas, et al. A draft human pangenome reference. Nature, 617 (7960):312–324, 2023. doi: 10.1038/s41586-023-05896-x.

Peter Ebert, Peter A Audano, Qihui Zhu, Bernardo Rodriguez-Martin, David Porubsky, Marc Jan Bonder, Arvis Sulovari, Jana Ebler, et al. Haplotype-resolved diverse human genomes and integrated analysis of structural variation. Science, 372(6537):eabf7117, 2021. doi: 10.1126/science.abf7117.

Daniela Cimini, Bonnie Howell, Paul Maddox, Alexey Khodjakov, Francesca Degrassi, and E. D. Salmon. Merotelic kinetochore orientation is a major mechanism of aneuploidy in mitotic mammalian tissue cells. The Journal of Cell Biology, 153(3):517–528, 2001. doi: 10.1083/jcb.153.3.517.

Agata P. Zielinska, Zuzana Holubcova, Martyn Blayney, Kay Elder, and Melina Schuh. Sister kinetochore splitting and precocious disintegration of bivalents could explain the maternal age effect. eLife, 4:e11389, 2015. doi: 10.7554/eLife.11389.

Leonardo Gomes de Lima et al. The formation and propagation of human Robertsonian chromosomes. Nature, 647(8091):952–961, 2025. doi: 10.1038/s41586-025-09540-8.

Viviana Barra and Daniele Fachinetti. The dark side of centromeres: types, causes and consequences of structural abnormalities implicating centromeric DNA. Nature Communications, 9:4340, 2018.

Xuelan Chen, Albert S Agustinus, Jun Li, et al. Chromosomal instability as a driver of cancer progression. Nature Reviews Genetics, 26(1):31–46, 2025. doi: 10.1038/s41576-024-00761-7.

Francesca Bersani, Eunjung Lee, Peter V Kharchenko, et al. Pericentromeric satellite repeat expansions through RNA-derived DNA intermediates in cancer. Proceedings of the National Academy of Sciences, 112(49):15148–15153, 2015. doi: 10.1073/pnas.1518008112.

Wei-Yang Bai et al. Genome-wide associations of structural variants with human traits through imputation from long-read assemblies. Nature Genetics, 58(6):1258–1267, 2026. doi: 10.1038/s41588-026-02612-z.

Colby Chiang, Alexandra J Scott, Joe R Davis, Emily K Tsang, Xin Li, Yungil Kim, et al. The impact of structural variation on human gene expression. Nature Genetics, 49(5):692–699, 2017. doi: 10.1038/ng.3834.

Alexandra J. Scott, Colby Chiang, and Ira M. Hall. Structural variants are a major source of gene expression differences in humans and often affect multiple nearby genes. Genome Research, 31(12):2249–2257, 2021. doi: 10.1101/gr.275488.121.

Julian K. Lucas, Prajna Hebbar, Wen-Wei Liao, Juan F. Macias-Velasco, Adam M. Novak, et al. HPRC2: A human pangenome reference with near-complete coverage of common genetic variation. bioRxiv, 2026. doi: 10.64898/2026.07.21.739710.

Daniel Taliun, Daniel N Harris, Michael D Kessler, Jedidiah Carlson, Zachary A Szpiech, Raul Torres, Sarah A Gagliano Taliun, André Corvelo, et al. Sequencing of 53,831 diverse genomes from the NHLBI TOPMed Program. Nature, 590(7845):290–299, 2021. doi: 10.1038/s41586-021-03205-y.

Shane McCarthy, Sayantan Das, Warren Kretzschmar, Olivier Delaneau, Andrew R Wood, Alexander Teumer, Hyun Min Kang, Christian Fuchsberger, et al. A reference panel of 64,976 haplotypes for genotype imputation. Nature Genetics, 48(10):1279–1283, 2016. doi: 10.1038/ng.3643.

Eric R Gamazon, Heather E Wheeler, Kaanan P Shah, Sahar V Mozaffari, Keston Aquino-Michaels, Robert J Carroll, Anne E Eyler, Joshua C Denny, et al. A gene-based association method for mapping traits using reference transcriptome data. Nature Genetics, 47(9):1091–1098, 2015.

Alexander Gusev, Arthur Ko, Huwenbo Shi, Gaurav Bhatia, Wonil Chung, Brenda WJH Penninx, Rick Jansen, Eco JC de Geus, Dorret I Boomsma, et al. Integrative approaches for large-scale transcriptome-wide association studies. Nature Genetics, 48(3):245–252, 2016. doi: 10.1038/ng.3506.

Sasha A Langley, Karen H Miga, Gary H Karpen, and Charles H Langley. Haplotypes spanning centromeric regions reveal persistence of large blocks of archaic DNA. eLife, 8:e42989, 2019.

Peter H Sudmant, Tobias Rausch, Eugene J Gardner, Robert E Handsaker, Alexej Abyzov, John Huddleston, Yan Zhang, Kai Ye, Goo Jun, Markus Hsi-Yang Fritz, et al. An integrated map of structural variation in 2,504 human genomes. Nature, 526(7571):75–81, 2015. doi: 10.1038/nature15394.

Peter A Audano, Arvis Sulovari, Tina A Graves-Lindsay, et al. Characterizing the major structural variant alleles of the human genome. Cell, 176(3):663–675.e19, 2019. doi: 10.1016/j.cell.2018.12.019.

Justin M Zook, Nancy F Hansen, Nathan D Olson, et al. A robust benchmark for detection of germline large deletions and insertions. Nature Biotechnology, 38(11):1347–1355, 2020. doi: 10.1038/s41587-020-0538-8.

Ryan L Collins, Harrison Brand, Konrad J Karczewski, et al. A structural variation reference for medical and population genetics. Nature, 581(7809):444–451, 2020. doi: 10.1038/s41586-020-2287-8.

Xiao Du et al. Robust benchmark structural variant calls of an Asian using state-of-the-art long-read sequencing technologies. Genomics, Proteomics & Bioinformatics, 20(1):192–204, 2022. doi: 10.1016/j.gpb.2020.10.006.

Jana Ebler, Peter Ebert, Wayne E Clarke, et al. Pangenome-based genome inference allows efficient and accurate genotyping across a wide spectrum of variant classes. Nature Genetics, 54(4):518–525, 2022. doi: 10.1038/s41588-022-01043-w.

Glennis A Logsdon et al. The variation and evolution of complete human centromeres. Nature, 629(8010):136–145, 2024. doi: 10.1038/s41586-024-07278-3.

Glennis A Logsdon, Peter Ebert, Peter A Audano, et al. Complex genetic variation in nearly complete human genomes. Nature, 644:430–441, 2025. doi: 10.1038/s41586-025-09140-6.

Heng Li. Minimap2: pairwise alignment for nucleotide sequences. Bioinformatics, 34(18):3094–3100, 2018.

David Heller and Martin Vingron. SVIM-asm: structural variant detection from haploid and diploid genome assemblies. Bioinformatics, 36(22-23):5519–5521, 2021.

Heng Li, Jonathan M Bloom, Yossi Farjoun, Mark Fleharty, Laura Gauthier, Benjamin Neale, and Daniel MacArthur. A synthetic-diploid benchmark for accurate variant-calling evaluation. Nature Methods, 15(8):595–597, 2018a. doi: 10.1038/s41592-018-0054-7.

Jonas A Gustafson et al. High-coverage nanopore sequencing of samples from the 1000 Genomes Project to build a comprehensive catalog of human genetic variation. Genome Research, 34 (11):2061–2073, 2024. doi: 10.1101/gr.279273.124.

Glennis A Logsdon, Mitchell R Vollger, PingHsun Hsieh, Yafei Mao, Mikhail A Liskovykh, Sergey Koren, et al. The structure, function and evolution of a complete human chromosome 8. Nature, 593(7857):101–107, 2021. doi: 10.1038/s41586-021-03420-7.

Dylan J Taylor et al. Sources of gene expression variation in a globally diverse human cohort. Nature, 632(8023):122–130, 2024. doi: 10.1038/s41586-024-07708-2.

Yuichi Shiraishi et al. Rare k-mers reveal centromere haplogroups underlying human diversity and cancer translocations. Cell, 2026. doi: 10.1016/j.cell.2026.08.053.

Stephen Rea et al. Regulation of chromatin structure by site-specific histone H3 methyltransferases. Nature, 406(6796):593–599, 2000. doi: 10.1038/35020506.

Antoine H F M Peters et al. Loss of the Suv39h histone methyltransferases impairs mammalian heterochromatin and genome stability. Cell, 107(3):323–337, 2001. doi: 10.1016/S0092-8674(01)00542-6.

Runze Ma, Yan Zhang, Jing Zhang, Pinqi Zhang, Zeqi Liu, Yiming Fan, Hao-Tian Wang, Zhuqiang Zhang, and Bing Zhu. Targeting pericentric non-consecutive motifs for heterochromatin initiation. Nature, 631(8021):678–685, 2024. doi: 10.1038/s41586-024-07640-5.

Ben L. Carty, Danilo Dubocanin, Marina Murillo-Pineda, Marie Dumont, Emilia Volpe, Pawel Mikulski, Julia Humes, Oliver Whittingham, Daniele Fachinetti, Simona Giunta, Nicolas Altemose, and Lars E. T. Jansen. Heterochromatin boundaries maintain centromere position, size and number. Nature Structural & Molecular Biology, 33(2):220–234, 2026. doi: 10.1038/s41594-025-01706-2.

Pragya Sidhwani, Jacob P. Schwartz, Kelsey A. Fryer, and Aaron F. Straight. Histone H3 lysine methyltransferase activities control compartmentalization of human centromeres. bioRxiv, 2025. doi: 10.1101/2025.07.01.662447. Preprint.

All of Us Research Program Genomics Investigators. Genomic data in the All of Us research program. Nature, 627:340–346, 2024. doi: 10.1038/s41586-023-06957-x.

Megan M. Shuey, William W. Stead, Ida Aka, et al. Next-generation phenotyping: introducing phecodeX for enhanced discovery research in medical phenomics. Bioinformatics, 39(11): btad655, 2023. doi: 10.1093/bioinformatics/btad655.

S. Saini et al. A reference haplotype panel for genome-wide imputation of short tandem repeats. Nature Communications, 9:4397, 2018. doi: 10.1038/s41467-018-06694-0.

Melissa Gymrek et al. Abundant contribution of short tandem repeats to gene expression variation in humans. Nature Genetics, 48:22–29, 2016.

Stephanie Feupe Fotsing et al. The impact of short tandem repeat variation on gene expression. Nature Genetics, 51:1652–1659, 2019.

Mehrdad Bakhtiari et al. Variable number tandem repeats mediate the expression of proximal genes. Nature Communications, 12:2075, 2021.

Ronen E Mukamel, Robert E Handsaker, Maxwell A Sherman, Alison R Barton, Yiming Zheng, Steven A McCarroll, and Po-Ru Loh. Protein-coding repeat polymorphisms strongly shape diverse human phenotypes. Science, 373(6562):1499–1505, 2021. doi: 10.1126/science.abg8289.

Boris Noyvert et al. Imputation of structural variants using a multi-ancestry long-read sequencing panel enables identification of disease associations. eLife, 14:RP106115, 2025. doi: 10.7554/eLife.106115.1. Reviewed Preprint.

Carsten Hain, Tobias Rausch, et al. HOROSCOPE: Decoding human centromere architecture from short reads using k-mer signatures. bioRxiv, 2026. doi: 10.64898/2026.06.10.731283. Preprint.

Catalina Salinas-Luypaert, Danilo Dubocanin, Rosa Jooyoung Lee, Lorena Andrade Ruiz, Riccardo Gamba, Marine Grison, Leonid Velikovsky, Annapaola Angrisani, et al. DNA methylation influences human centromere positioning and function. Nature Genetics, 57(10):2509–2521, 2025. doi: 10.1038/s41588-025-02324-w.

Yanqing Sun, Shijie Wan, Lei Nie, Dan Yu, Feifei Zhou, Yiqing Yang, Xiangyu Yang, Anguo Liu, Quanyu Chen, Kezhi Fu, Qingyang Ni, Yaoxi He, Bing Su, Yafei Mao, Kai Ye, Xiaofei Yang, Guojie Zhang, and Dongya Wu. Multidimensional variation and population stratification across 8000 complete human centromeres. bioRxiv, 2026. doi: 10.64898/2026.07.22.740206.

Shenghan Gao, Keisuke K. Oshima, Shu-Cheng Chuang, Mark Loftus, Tamara A. Potapova, Annalaura Montanari, David S. Gordon, Zikun Yang, et al. A global view of human centromere variation and evolution. Nature, 2026. doi: 10.1038/s41586-026-10841-9.

Arang Rhie, Sergey Nurk, Monika Cechova, Savannah J. Hoyt, et al. The complete sequence of a human Y chromosome. Nature, 621(7978):344–354, 2023. doi: 10.1038/s41586-023-06457-y.

Ivan Alexandrov, Alexei Kazakov, Irina Tumeneva, Valery Shepelev, and Yuri Yurov. Alpha-satellite DNA of primates: old and new families. Chromosoma, 110(4):253–266, 2001. doi: 10.1007/s004120100146.

Valery A Shepelev, Lev I Uralsky, Alexander A Alexandrov, Yuri B Yurov, Evgeny I Rogaev, and Ivan A Alexandrov. Annotation of suprachromosomal families reveals uncommon types of alpha satellite organization in pericentromeric regions of hg38 human genome assembly. Genomics Data, 5:139–146, 2015. doi: 10.1016/j.gdata.2015.05.035.

Naruya Saitou and Masatoshi Nei. The neighbor-joining method: a new method for reconstructing phylogenetic trees. Molecular Biology and Evolution, 4(4):406–425, 1987.

Marta Byrska-Bishop, Uday S Evani, Xuefang Zhao, Anna O Basile, Haley J Abel, Allison A Regier, André Corvelo, Wayne E Clarke, et al. High-coverage whole-genome sequencing of the expanded 1000 Genomes Project cohort including 602 trios. Cell, 185(18):3426–3440.e19, 2022. doi: 10.1016/j.cell.2022.08.004.

Terrence S. Furey and David Haussler. Integration of the cytogenetic map with the draft human genome sequence. Human Molecular Genetics, 12(9):1037–1044, 2003. doi: 10.1093/hmg/ddg113.

W. James Kent, Charles W. Sugnet, Terrence S. Furey, Krishna M. Roskin, Tom H. Pringle, Alan M. Zahler, and David Haussler. The human genome browser at UCSC. Genome Research, 12(6):996–1006, 2002.

Hui Zou and Trevor Hastie. Regularization and variable selection via the elastic net. Journal of the Royal Statistical Society Series B: Statistical Methodology, 67(2):301–320, 2005. doi: 10.1111/j.1467-9868.2005.00503.x.

Fabian Pedregosa, Gaël Varoquaux, Alexandre Gramfort, Vincent Michel, Bertrand Thirion, Olivier Grisel, Mathieu Blondel, Peter Prettenhofer, et al. Scikit-learn: machine learning in Python. Journal of Machine Learning Research, 12:2825–2830, 2011.

Jonathan Marchini and Bryan Howie. Genotype imputation for genome-wide association studies. Nature Reviews Genetics, 11(7):499–511, 2010. doi: 10.1038/nrg2796.

Sayantan Das, Lukas Forer, Sebastian Schönherr, Carlo Sidore, Adam E Locke, Alan Kwong, Scott I Vrieze, Emily Y Chew, et al. Next-generation genotype imputation service and methods. Nature Genetics, 48(10):1284–1287, 2016.

Yang I Li, David A Knowles, Jack Humphrey, Alvaro N Barbeira, Scott P Dickinson, Hae Kyung Im, and Jonathan K Pritchard. Annotation-free quantification of RNA splicing using Leaf-Cutter. Nature Genetics, 50:151–158, 2018b. doi: 10.1038/s41588-017-0004-9.

Amaro Taylor-Weiner, François Aguet, Nicholas J Haradhvala, Sager Gosai, Shankara Anand, Jaegil Kim, Kristin Ardlie, Eliezer M Van Allen, et al. Scaling computational genomics to millions of individuals with GPUs. Genome Biology, 20(1):228, 2019. doi: 10.1186/s13059-019-1836-7.

Oliver Stegle, Leopold Parts, Matias Piipari, John Winn, and Richard Durbin. Using probabilistic estimation of expression residuals (PEER) to obtain increased power and interpretability of gene expression analyses. Nature Protocols, 7(3):500–507, 2012. doi: 10.1038/nprot.2011.457.

Halit Ongen, Alfonso Buil, Andrew A Brown, Emmanouil T Dermitzakis, and Olivier Delaneau. Fast and efficient QTL mapper for thousands of molecular phenotypes. Bioinformatics, 32 (10):1479–1485, 2016.

John D Storey and Robert Tibshirani. Statistical significance for genomewide studies. Proceedings of the National Academy of Sciences, 100(16):9440–9445, 2003.

Ashis Saha and Alexis Battle. False positives in trans-eQTL and co-expression analyses arising from RNA-sequencing alignment errors. F1000Research, 7:1860, 2018a. doi: 10.12688/f1000research.17145.2.

Moritz Smolka, Luis F. Paulin, Christopher M. Grochowski, Dominic W. Horner, Medhat Mahmoud, Sairam Behera, et al. Detection of mosaic and population-level structural variants with Sniffles2. Nature Biotechnology, 42(10):1571–1580, 2024. doi: 10.1038/s41587-023-02024-y.

Gao Wang, Abhishek Sarkar, Peter Carbonetto, and Matthew Stephens. A simple new approach to variable selection in regression, with application to genetic fine mapping. Journal of the Royal Statistical Society: Series B, 82(5):1273–1300, 2020. doi: 10.1111/rssb.12388.

Aaron R Quinlan and Ira M Hall. BEDTools: a flexible suite of utilities for comparing genomic features. Bioinformatics, 26(6):841–842, 2010. doi: 10.1093/bioinformatics/btq033.

Francesco Kumara Mastrorosa, Keisuke K Oshima, Allison N Rozanski, William T Harvey, Evan E Eichler, and Glennis A Logsdon. Identification and annotation of centromeric hypomethylated regions with CDR-Finder. Bioinformatics, 40(12):btae733, 2024. doi: 10.1093/bioinformatics/btae733.

Petr Danecek, James K. Bonfield, Jennifer Liddle, John Marshall, Valeriu Ohan, Martin O. Pollard, Andrew Whitwham, Thomas Keane, Shane A. McCarthy, Robert M. Davies, and Heng Li. Twelve years of SAMtools and BCFtools. GigaScience, 10(2):giab008, 2021. doi: 10.1093/gigascience/giab008.

David Freedman and David Lane. A nonstochastic interpretation of reported significance levels. Journal of Business and Economic Statistics, 1(4):292–298, 1983. doi: 10.1080/07350015.1983.10509354.

Olga Kunyavskaya, Tatiana Dvorkina, Andrey V Bzikadze, Ivan A Alexandrov, and Pavel A Pevzner. Automated annotation of human centromeres with HORmon. Genome Research, 32 (6):1137–1151, 2022. doi: 10.1101/gr.276362.121.

Tatiana Dvorkina, Andrey V Bzikadze, and Pavel A Pevzner. The string decomposition problem and its applications to centromere analysis and assembly. Bioinformatics, 36:i93–i101, 2020. doi: 10.1093/bioinformatics/btaa454.

Christopher C. Chang, Carson C. Chow, Laurent C. A. M. Tellier, Shashaank Vattikuti, Shaun M. Purcell, and James J. Lee. Second-generation PLINK: rising to the challenge of larger and richer datasets. GigaScience, 4(1):7, 2015. doi: 10.1186/s13742-015-0047-8.

David Firth. Bias reduction of maximum likelihood estimates. Biometrika, 80(1):27–38, 1993. doi: 10.1093/biomet/80.1.27.

Georg Heinze and Michael Schemper. A solution to the problem of separation in logistic regression. Statistics in Medicine, 21(16):2409–2419, 2002. doi: 10.1002/sim.1047.

Yoav Benjamini and Yosef Hochberg. Controlling the false discovery rate: a practical and powerful approach to multiple testing. Journal of the Royal Statistical Society: Series B, 57 (1):289–300, 1995.

Adam C. English, Vipin K. Menon, Richard A. Gibbs, Ginger A. Metcalf, and Fritz J. Sedlazeck. Truvari: refined structural variant comparison preserves allelic diversity. Genome Biology, 23 (1):271, 2022. doi: 10.1186/s13059-022-02840-6.

Kishwar Shafin, Trevor Pesout, Ryan Lorig-Roach, et al. Nanopore sequencing and the Shasta toolkit enable efficient de novo assembly of eleven human genomes. Nature Biotechnology, 38 (9):1044–1053, 2020. doi: 10.1038/s41587-020-0503-6.

Brian L Browning, Ying Zhou, and Sharon R Browning. A one-penny imputed genome from next-generation reference panels. The American Journal of Human Genetics, 103(3):338–348, 2018.

Na Li and Matthew Stephens. Modeling linkage disequilibrium and identifying recombination hotspots using single-nucleotide polymorphism data. Genetics, 165(4):2213–2233, 2003.

Ashis Saha and Alexis Battle. Mappability resources for GRCh38 using Gencode v26 annotation. figshare dataset, 2018b. doi: 10.6084/m9.figshare.7315136.v3.

Alber Aqil, Betty Y. H. Huang, Pavlos Pavlidis, Rajiv C. McCoy, and Omer Gokcumen. Mismapping of sequencing reads from polymorphic duplications generates spurious trans-eQTLs. The American Journal of Human Genetics, 113(8):1604–1617, 2026. doi: 10.1016/j.ajhg.2026.07.004.

Tatiana Dvorkina, Olga Kunyavskaya, Andrey V Bzikadze, Ivan Alexandrov, and Pavel A Pevzner. CentromereArchitect: inference and analysis of the architecture of centromeres. Bioinformatics, 37(Supplement_1):i196–i204, 2021. doi: 10.1093/bioinformatics/btab265.

Sina Rüeger, Aaron McDaid, and Zoltán Kutalik. Evaluation and application of summary statistic imputation to discover new height-associated loci. PLoS Genetics, 14(5):e1007371, 2018.

Yuxin Zou, Peter Carbonetto, Gao Wang, and Matthew Stephens. Fine-mapping from summary data with the “Sum of Single Effects” model. PLoS Genetics, 18(7):e1010299, 2022. doi: 10.1371/journal.pgen.1010299.

Siming Zhao, Wesley Crouse, Sheng Qian, Kaixuan Luo, Matthew Stephens, and Xin He. Adjusting for genetic confounders in transcriptome-wide association studies improves discovery of risk genes of complex traits. Nature Genetics, 56:336–347, 2024. doi: 10.1038/s41588-023-01648-9.

Mitja I. Kurki et al. FinnGen provides genetic insights from a well-phenotyped isolated population. Nature, 613:508–518, 2023.

Tomaz Berisa and Joseph K. Pickrell. Approximately independent linkage disequilibrium blocks in human populations. Bioinformatics, 32(2):283–285, 2016. doi: 10.1093/bioinformatics/btv546.

Alvaro N. Barbeira et al. Exploring the phenotypic consequences of tissue specific gene expression variation inferred from GWAS summary statistics. Nature Communications, 9:1825, 2018. doi: 10.1038/s41467-018-03621-1.

Bogdan Pasaniuc et al. Fast and accurate imputation of summary statistics enhances evidence of functional enrichment. Bioinformatics, 30(20):2906–2914, 2014.

Joonas Kartau and Matti Pirinen. FINEMAP-miss: fine-mapping genome-wide association studies with missing genotype information. Bioinformatics, 41(12):btaf616, 2025. doi: 10.1093/bioinformatics/btaf616.

Mohamed A Almarri, Anders Bergström, Javier Prado-Martinez, Fengtang Yang, Beiyuan Fu, Alistair S Dunham, Yuan Chen, Matthew E Hurles, Chris Tyler-Smith, and Yali Xue. Population structure, stratification, and introgression of human structural variation. Cell, 182(1): 189–199.e15, 2020. doi: 10.1016/j.cell.2020.05.024.

Gaurav Bhatia, Nick Patterson, Sriram Sankararaman, and Alkes L Price. Estimating and interpreting FST: the impact of rare variants. Genome Research, 23(9):1514–1521, 2013.

